# FinnGen: Unique genetic insights from combining isolated population and national health register data

**DOI:** 10.1101/2022.03.03.22271360

**Authors:** Mitja I. Kurki, Juha Karjalainen, Priit Palta, Timo P. Sipilä, Kati Kristiansson, Kati Donner, Mary P. Reeve, Hannele Laivuori, Mervi Aavikko, Mari A. Kaunisto, Anu Loukola, Elisa Lahtela, Hannele Mattsson, Päivi Laiho, Pietro Della Briotta Parolo, Arto Lehisto, Masahiro Kanai, Nina Mars, Joel Rämö, Tuomo Kiiskinen, Henrike O. Heyne, Kumar Veerapen, Sina Rüeger, Susanna Lemmelä, Wei Zhou, Sanni Ruotsalainen, Kalle Pärn, Tero Hiekkalinna, Sami Koskelainen, Teemu Paajanen, Vincent Llorens, Javier Gracia-Tabuenca, Harri Siirtola, Kadri Reis, Abdelrahman G. Elnahas, Katriina Aalto-Setälä, Kaur Alasoo, Mikko Arvas, Kirsi Auro, Shameek Biswas, Argyro Bizaki-Vallaskangas, Olli Carpen, Chia-Yen Chen, Oluwaseun A. Dada, Zhihao Ding, Margaret G. Ehm, Kari Eklund, Martti Färkkilä, Hilary Finucane, Andrea Ganna, Awaisa Ghazal, Robert R. Graham, Eric Green, Antti Hakanen, Marco Hautalahti, Åsa Hedman, Mikko Hiltunen, Reetta Hinttala, Iiris Hovatta, Xinli Hu, Adriana Huertas-Vazquez, Laura Huilaja, Julie Hunkapiller, Howard Jacob, Jan-Nygaard Jensen, Heikki Joensuu, Sally John, Valtteri Julkunen, Marc Jung, Juhani Junttila, Kai Kaarniranta, Mika Kähönen, Risto M. Kajanne, Lila Kallio, Reetta Kälviäinen, Jaakko Kaprio, Nurlan Kerimov, Johannes Kettunen, Elina Kilpeläinen, Terhi Kilpi, Katherine Klinger, Veli-Matti Kosma, Teijo Kuopio, Venla Kurra, Triin Laisk, Jari Laukkanen, Nathan Lawless, Aoxing Liu, Simonne Longerich, Reedik Mägi, Johanna Mäkelä, Antti Mäkitie, Anders Malarstig, Arto Mannermaa, Joseph Maranville, Athena Matakidou, Tuomo Meretoja, Sahar V. Mozaffari, Mari EK. Niemi, Marianna Niemi, Teemu Niiranen, Christopher J. O’Donnell, Ma’en Obeidat, George Okafo, Hanna M. Ollila, Antti Palomäki, Tuula Palotie, Jukka Partanen, Dirk S. Paul, Margit Pelkonen, Rion K. Pendergrass, Slavé Petrovski, Anne Pitkäranta, Adam Platt, David Pulford, Eero Punkka, Pirkko Pussinen, Neha Raghavan, Fedik Rahimov, Deepak Rajpal, Nicole A. Renaud, Bridget Riley-Gillis, Rodosthenis Rodosthenous, Elmo Saarentaus, Aino Salminen, Eveliina Salminen, Veikko Salomaa, Johanna Schleutker, Raisa Serpi, Huei-yi Shen, Richard Siegel, Kaisa Silander, Sanna Siltanen, Sirpa Soini, Hilkka Soininen, Jae H. Sul, Ioanna Tachmazidou, Kaisa Tasanen, Pentti Tienari, Sanna Toppila-Salmi, Taru Tukiainen, Tiinamaija Tuomi, Joni A. Turunen, Jacob C. Ulirsch, Felix Vaura, Petri Virolainen, Jeffrey Waring, Dawn Waterworth, Robert Yang, Mari Nelis, Anu Reigo, Andres Metspalu, Lili Milani, Tõnu Esko, Caroline Fox, Aki S. Havulinna, Markus Perola, Samuli Ripatti, Anu Jalanko, Tarja Laitinen, Tomi Mäkelä, Robert Plenge, Mark McCarthy, Heiko Runz, Mark J. Daly, Aarno Palotie

## Abstract

Population isolates such as Finland provide benefits in genetic studies because the allelic spectrum of damaging alleles in any gene is often concentrated on a small number of low-frequency variants (0.1% ≤ minor allele frequency < 5%), which survived the founding bottleneck, as opposed to being distributed over a much larger number of ultra--rare variants. While this advantage is well-- established in Mendelian genetics, its value in common disease genetics has been less explored. FinnGen aims to study the genome and national health register data of 500,000 Finns, already reaching 224,737 genotyped and phenotyped participants. Given the relatively high median age of participants (63 years) and dominance of hospital-based recruitment, FinnGen is enriched for many disease endpoints often underrepresented in population-based studies (e.g., rarer immune-mediated diseases and late onset degenerative and ophthalmologic endpoints). We report here a genome-wide association study (GWAS) of 1,932 clinical endpoints defined from nationwide health registries. We identify genome--wide significant associations at 2,491 independent loci. Among these, finemapping implicates 148 putatively causal coding variants associated with 202 endpoints, 104 with low allele frequency (AF<10%) of which 62 were over two-fold enriched in Finland.

We studied a benchmark set of 15 diseases that had previously been investigated in large genome-wide association studies. FinnGen discovery analyses were meta-analysed in Estonian and UK biobanks. We identify 30 novel associations, primarily low-frequency variants strongly enriched, in or specific to, the Finnish population and Uralic language family neighbors in Estonia and Russia.

These findings demonstrate the power of bottlenecked populations to find unique entry points into the biology of common diseases through low-frequency, high impact variants. Such high impact variants have a potential to contribute to medical translation including drug discovery.

## Main

Large biobank studies have become an important source for genetic discoveries. The FinnGen study aims to construct a resource combining the power of nationwide biobanks, structured national healthcare data and a unique, isolated population. Due to increased genetic drift, isolated populations with recent bottlenecks can have deleterious, disease predisposing alleles at considerably higher frequencies than permitted by selection in larger and older outbred populations. Counterbalancing this enrichment of specific low-frequency alleles, the other consequence of a recent bottleneck is that isolated populations have considerably fewer very rare variants overall^1–3^. As a result, isolated populations provide an opportunity to identify high-impact disease-causing variants that are extremely rare in other populations^1, 4–7^. In Finland, a strong founding bottleneck occurred ∼120 generations ago, followed by rapid population expansion. This bottleneck effect has resulted in numerous strongly deleterious alleles that are more frequent in Finland compared to other Europeans. This is manifested in the Finnish Disease Heritage, a set of 36 mostly recessive diseases that are more prevalent in Finland than elsewhere in the world^8^. This population history (facilitating identification of low frequency deleterious alleles) combined with longitudinal information from registers recording hospital in-patient and outpatient diagnoses, purchases of prescription medications and many other national health registries centrally collected for decades provides unique opportunities for understanding the genetic basis of health and disease.

FinnGen is a public-private partnership research project combining imputed genotype data generated from newly collected and legacy samples of Finnish biobanks and digital health record data from Finnish health registries (https://www.finngen.fi/en) aiming to provide new insight in disease genetics. It includes nine Finnish biobanks, universities and university hospitals, 13 international pharmaceutical industry partners and Finnish biobank cooperative (FINBB) in a pre-competitive partnership. As of August 2020 (Release 5 described in this paper) samples from 412,000 individuals had been collected and 224,737 analysed with an aim of a cohort of 500 000 participants (See Supplementary Methods Section 2). The project utilizes the nationwide longitudinal health register data collected since 1969 from every resident in Finland.

Here, we describe the FinnGen project and its current genotype and phenotype content and highlight a series of genetic discoveries from the first data collection phase. In accompanying manuscripts, we describe more detailed studies showcasing different aspects of the rich data available from population registries. We first show that FinnGen register-based phenotypes are comparable to those used in disease specific genome-wide association studies (GWAS) in 15 previously well-studied common diseases. We demonstrate the power of the combination of isolated population and register data to discover novel low frequency variant associations, even in previously well-studied diseases where FinnGen has a much smaller number of cases than in published disease specific GWASs. Finally, via genome-wide association study of 1932 endpoints followed by statistical fine-mapping, we demonstrate the ability to identify likely causal coding variants even with very low allele frequencies.

### Phenotyping and genotyping

In Finland, similar to the other Nordic countries, there are nationwide electronic health registers originally established primarily for administrative purposes to monitor the usage of healthcare nationwide and over the lifespan of each Finnish resident. These registers have almost complete coverage of major health-related events, such as hospitalizations, prescription drug purchases (not including hospital administered medications), medical procedures or deaths with a history of data collection spanning more than 50 years. Health register based phenotypes (“endpoints”) were created by combining data (mainly using International Classification of Diseases (ICD) and Anatomical Chemical Therapeutic (ACT) classification codes) from one or more nationwide heath registers (Supplementary Table 1). For a phenome-wide GWAS, we have initially constructed more than 2800 endpoints by combining data from different health registers including hospital discharge register, prescription medication purchase register and cancer register (Figure 1, See Supplementary Methods section 1 and https://r5.risteys.finngen.fi/).

**Figure 1.**
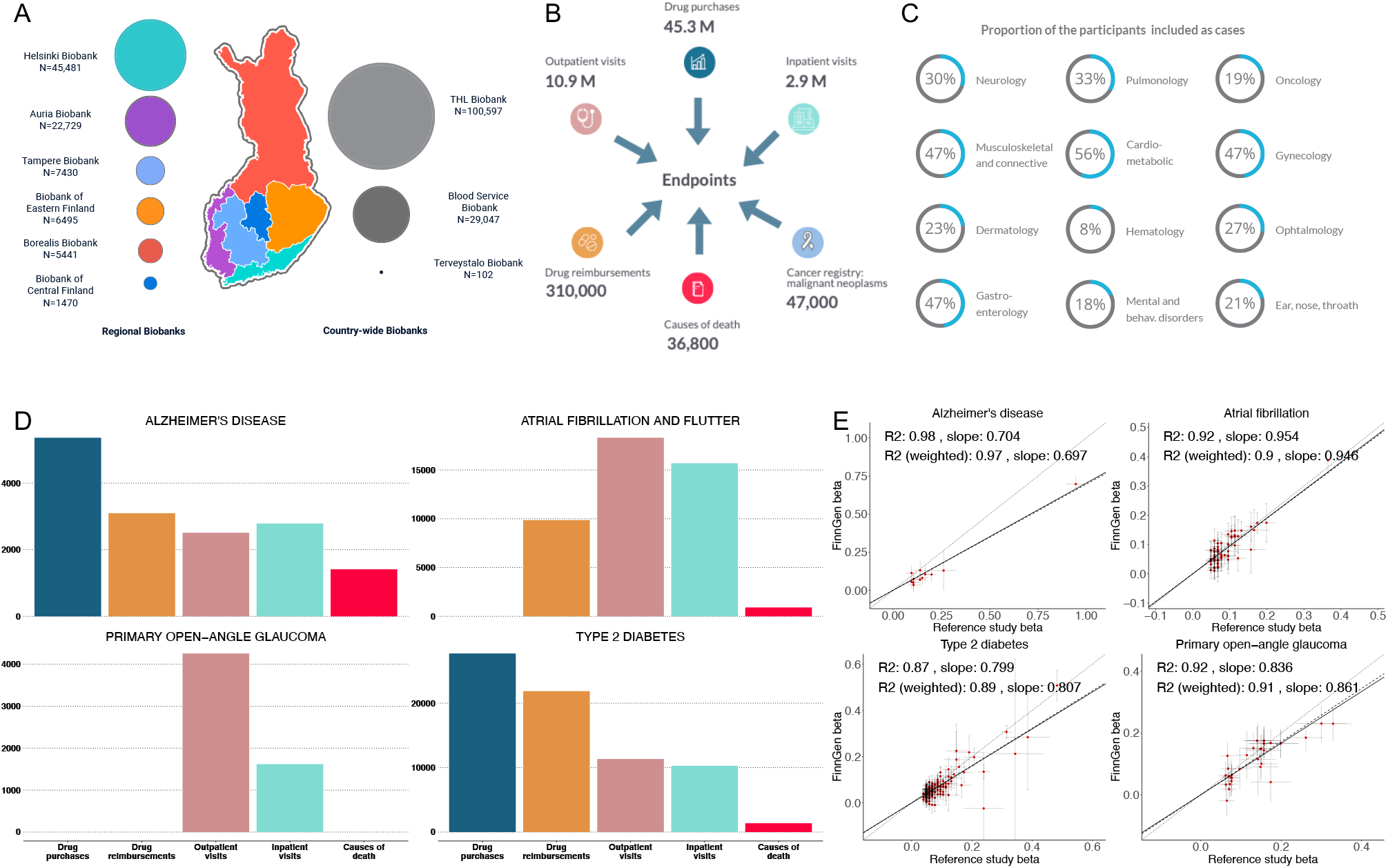
**A) Sample numbers collected from different geographical areas:** The map of Finland is divided into major administrative areas. Colors in the geographical regions represent the catchment areas of each of the nine biobanks providing samples to FinnGen. The size of the circle next to each biobank name represents relative sample sizes. The number of samples given are those that were used in the analyses after QC. The THL, Blood service and Terveystalo biobanks are not regional like the other six and are separately presented on the right side of the map. B) **National registries utilized to construct FinnGen phenotype endpoints.** The numbers indicate the number of events in each register at the time of FinnGen release. The same individual can have multiple diagnosis and can have health information in multiple registers that contribute to individuals endpoint data 5. **C) Sample prevalence of major disease categories in FinnGen.** Major representative diseases for each category were chosen for demonstration purposes (See Supplementary Table 3 and 4 for the selected diseases). Same study subject might be represented in more than one disease category. **5. D) Examples of register codes used for constructing 4 selected endpoints from the 15 highlighted diseases.** The number represents matching register codes according to each phenotype definition in FinnGen. Each individual can contribute only once to each register but the same individual can be counted in multiple registers. **E) Comparison of effects sizes** (betas) **in known genome wide significant loci between four example FinnGen endpoints (D) and large published reference GWAS.** Y and X axes represent FinnGen and reference GWAS betass respectively. Betas are aligned to be positive in reference studies. Lines extending from points indicate standard errors of betas in respective studies. Regression lines omit intercept and two types of regressions are provided: unweighted and weighted by pooled standard errors from the two studies. Solid line indicates identity line and dotted line and dashed lines indicate unweighted and weighted regression respectively. Only variants with p < 10^-10^ in reference study were included to mitigate the effect of winner’s curse of inflated betas in reference studies. See beta comparison of all 15 diseases in Supplementary File 1.

FinnGen Release 5 presented here contains genotype data for 224,737 individuals after quality control (QC). A total of 154,714 individuals were genotyped with a custom Axiom FinnGen1 array. Data on 70,023 additional individuals were derived from legacy collections (Supplementary Table 2) genotyped with non-custom genotyping arrays (See Supplementary Methods Section 3 for QC details). We developed and utilized a population specific imputation reference panel of 3,775 high-coverage (25-30x) whole-genome sequenced (WGS) Finns, containing 16,962,023 SNPs and INDELS (minor allele count>=3) (See Supplementary Methods section 3). The vast majority (16 387 711) of the variants were confidently imputed (INFO>0.6, See Supplementary Figure 5).

### Population structure and cryptic relatedness

To study the genetic ancestry of 224,737 FinnGen participants passing genotyping QC (see Supplementary Methods Section 3), we combined the FinnGen data with 2504 phase 3 1000 Genomes Project reference samples^9^ and used principal component analysis (PCA) to identify FinnGen participants that have non-Finnish genetic ancestry. The vast majority were of broadly Finnish ancestry with only 3,676/224,737 (1.63%) outliers removed (see Supplementary Methods Section 4, Supplementary Figure 6). We estimated that 165,448 individuals (73.6%) have 3rd degree or closer relatives among FinnGen participants, which is higher than the estimated 30.3% in the UKBB^10^, but is partially explained by family based legacy cohorts. We removed 5780 duplicates/monozygotic twins (one from each pair removed) and genetic population outliers (See Supplementary Methods section 4) and built a set of approximately unrelated individuals where the relation between any couple is of degree 3 or higher. In total there were 156,977 independent individuals, which were used to compute PCA and the 61,980 related individuals were projected onto those PCs (see Supplementary Methods Section 4). The first two PCs captured the well-known east-west and north-south genetic differences in Finland (Figure 1B)^11^. Out of the total of 218,957 genotyped samples remaining, we had phenotype data for 218,792 individuals (56.5% females [123,579]), which were then used in all analyses.

### Genome-wide association studies utilizing nationwide health registries

To benchmark our register-based phenotyping and explore the value of the isolated setting of Finland, we selected 15 diseases with over 1000 cases in FinnGen and for which well-powered GWASs have been published. We evaluated the accuracy of our phenotyping by comparing the genetic correlations and effect sizes with the earlier GWAS results (Supplementary Table 6). None of the genetic correlations (GC) were significantly lower than 1 (lowest GC 0.89 [SE: 0.07] in AMD, Supplementary Table 6). For diseases with a large number of cases in FinnGen the effect sizes of lead variants in known loci were largely consistent between FinnGen and the literature meta-analyses, demonstrating that our register-based phenotyping is comparable to existing disease-specific GWAS studies (Figure 1E, Supplementary File 1). The effect sizes expectedly varied more in diseases with a smaller number of cases in FinnGen.

GWAS of these 15 diseases identified 235 loci and 275 independent genome-wide significant associations outside of the HLA region (GRCh38 Chr6:25Mb-34Mb). FinnGen PheWAS of Imputed classical HLA gene alleles is reported by Ritari et al^12^. 44 of the non-HLA associations were driven by low frequency (we define “low frequency” as AF < 0.1 in FinnGen) lead variants that were over two-fold enriched in Finns as compared to non-Finnish-Swedish-Estonian European (NFSEE) in gnomAD v2.0.1^13^. We use NFSEE as a general continental European reference point, excluding individuals from Finland, Sweden and Estonia. As there were large-scale migrations from Finland to Sweden in the 20th century, many of the chromosomes from Swedish sequencing studies are of recent Finnish origin, and the geographically close and linguistically and genetically similar^13^ population of Estonia is likely to share elements of the same ancestral founder effect.

Replication of many such Finnish-enriched variant associations is hindered by low allele frequencies or missingness in other European populations. As Finns are genetically more similar to Estonians than other Europeans^13^, we therefore first conducted replication in 136,724 individuals from the Estonian biobank and then extended to UKBB (see Supplementary Table 7 for endpoint definitions and case/control numbers and Methods). The effect sizes in genome-wide significant hits in FinnGen were mostly concordant between Estonia (average inverse variance weighted slope 1.5 [FinnGen higher] and r2 0.69) and UKBB (slope 1.1, r2 0.84) (Supplementary Figure 8). Most likely due to slightly different ascertainment schemes, FinnGen had higher case prevalence in the 15 disease diagnoses than UKBB, however the Estonian biobank had the highest case prevalence in ophthalmic diseases (age-related macular degeneration, glaucoma) and inflammatory skin conditions (atopic dermatitis and psoriasis) (Figure 2A).

**Figure 2.**
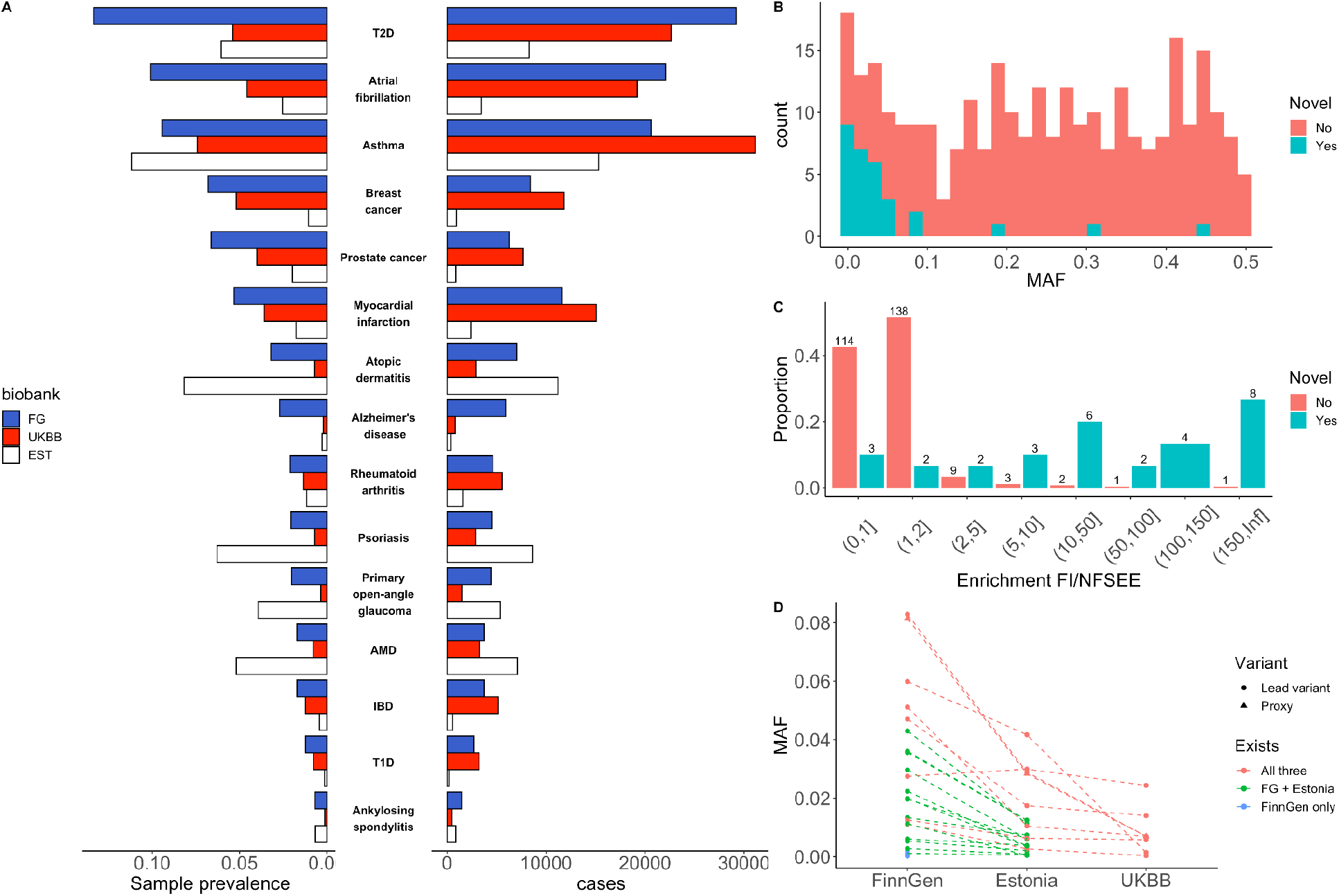
Comparison of novel and known unique lead variants in loci identified in the 15 studied diseases. A) Case prevalences and counts in FinnGen, EstBB and UKBB. The phenotypes are sorted by FinnGen prevalence B) Distribution of minor allele frequencies in known (red) and novel (blue) loci. C) Distribution of allele frequency enrichment between Finland and other North-Western Europeans in GnomAD (excluding Estonia and Sweden). D) Allele frequencies of 25 replicated genome-wide significant (in FinnGen discovery) novel low frequency (< 10%) unique variants in FinnGen, Estonia and UKBB. Dotted line indicates the same variants and no line means absence of the variant in other biobanks. AMD, Age-related Macular Degeneration; IBD, Inflammatory Bowel Disease; T1D, Type 1 Diabetes; T2D, Type 2 diabetes

After meta-analysis with Estonia and UKBB, 241 of the 275 associations remained genome-wide significant (Supplementary Table 8). We further meta-analysed 232 loci that did not meet the genome-wide significance threshold in FinnGen (5 * 10^-8^< p < 1* 10^-6^) and 57 of those were genome-wide significant after meta-analysis, resulting in 298 genome-wide significant meta-analysed independent association signals (See Supplementary Table 8 also for results after multiple testing correction for 15 endpoints).

In order to determine whether the observed associations had been previously reported, we queried the GWAS catalog (and largest recent relevant GWASs) for genome-wide significant (p < 5*10^-8^) variants that are in linkage disequilibrium (LD) (r2>0.1 in FinnGen imputation panel) with observed lead variants in FinnGen. As the lowest allele frequency of novel findings was low (0.15%), in addition to published GWASs, we checked if credible set variants in these loci have also been previously reported in ClinVar^14^. We observed multiple, known pathogenic variants such as a frameshift variant in *PALB2* (p.Leu531fs, AF 0.1%, not observed outside of Finland in gnomAD, Supplementary Table 8) associated with breast cancer. Thirty of the 298 associations were not previously reported in the largest published meta-analyses or GWAS catalog (Supplementary Table 8). As expected, we observed that lead variants in novel loci were mostly of low frequency (27 lead variants had MAF<10% in FinnGen) and enriched in Finland as compared to known loci from previous GWAS. In most cases, the allele frequencies of lower frequency variants (MAF < 10% in FinnGen) were the highest in FinnGen followed by Estonia and lowest in NFSEE in gnomAD (Figure 2C).

Next, we performed statistical fine-mapping of FinnGen associations (see Methods) on all 298 genome-wide significant associations and observed a coding variant (missense, frameshift, canonical splice site, stop gained, stop lost, inframe deletion) with posterior inclusion probability (PIP) >= 0.05 in 44 (18.7%) of the 95% credible sets (17 coding variants had PIP>0.5). Here onwards, we report coding variants with PIP>0.05 as “putatively causal”. We recognize that there may be occasions where the causal variant assignment to a coding variant is incorrect (see our accompanying papers^15, 16^ for discussions on fine-mapping calibration and replicability). In addition to identifying putative causal coding variants, we sought to identify potential gene expression regulatory mechanisms by colocalizing credible sets with fine-mapped expression quantitative trait locus (eQTL) datasets from GTEx^17^ and eQTL catalogue^18^ (see Methods).

To describe the allele frequency spectrum and putative mechanisms of action of risk variants we LD pruned the 298 genome wide significant associations and prioritised the most significant phenotype among the same hits to represent a single putative causal variant (LD r2 between lead variants < 0.2), resulting in 281 unique associations.

The majority of the 281 unique associations were common variant associations, however 73 of those had a lead variant frequency less than 10% in FinnGen and 38 of them were more than two times enriched in Finland vs. NFSEE. We observed a coding variant more often in the credible sets of over two-fold enriched associations (19/38, 50%) than non-enriched associations (9/35, 25.7%) at lower frequencies (MAF < 10%).

Following the discovery of 27 novel unique associations for 30 endpoints, we sought to determine potential mechanisms of action through identification of coding variants in their credible sets and potential regulatory effects by colocalization with eQTL associations from GTEx and eQTL catalogue^18^. We identified putative causal coding variants in 9/27 loci and eQTL colocalization in 5/27 loci. In three out of the five eqtl loci, we observed a coding variant in credible sets (ILR4,MYH14,CFI). The two remaining eQTLs colocalizations were breast cancer locus colocalizing with H2BP2 eQTL in lung tissue and T2D colocalizing with PRRG4 in lipopolysaccharide stimulated monocytes. The disease relevance of these eQTLs is not evident. However, no credible coding variants or eQTL were identified in 16/27 loci (Table 1, Supplementary Table 8). The fraction of associations where we observed eQTL was small (14.8%). The vast majority of the novel associations are driven by variants with very low allele frequency in NFSEE populations (Table 1, Figure 1 B,D). The low fraction of eQTL colocalizatons observed is likely explained by low allele frequency of 25/27 of the variants in available eQTL studies (such as GTEx), where the majority of the samples don’t have Finnish/Estonian ancestry.

**Table 1.**
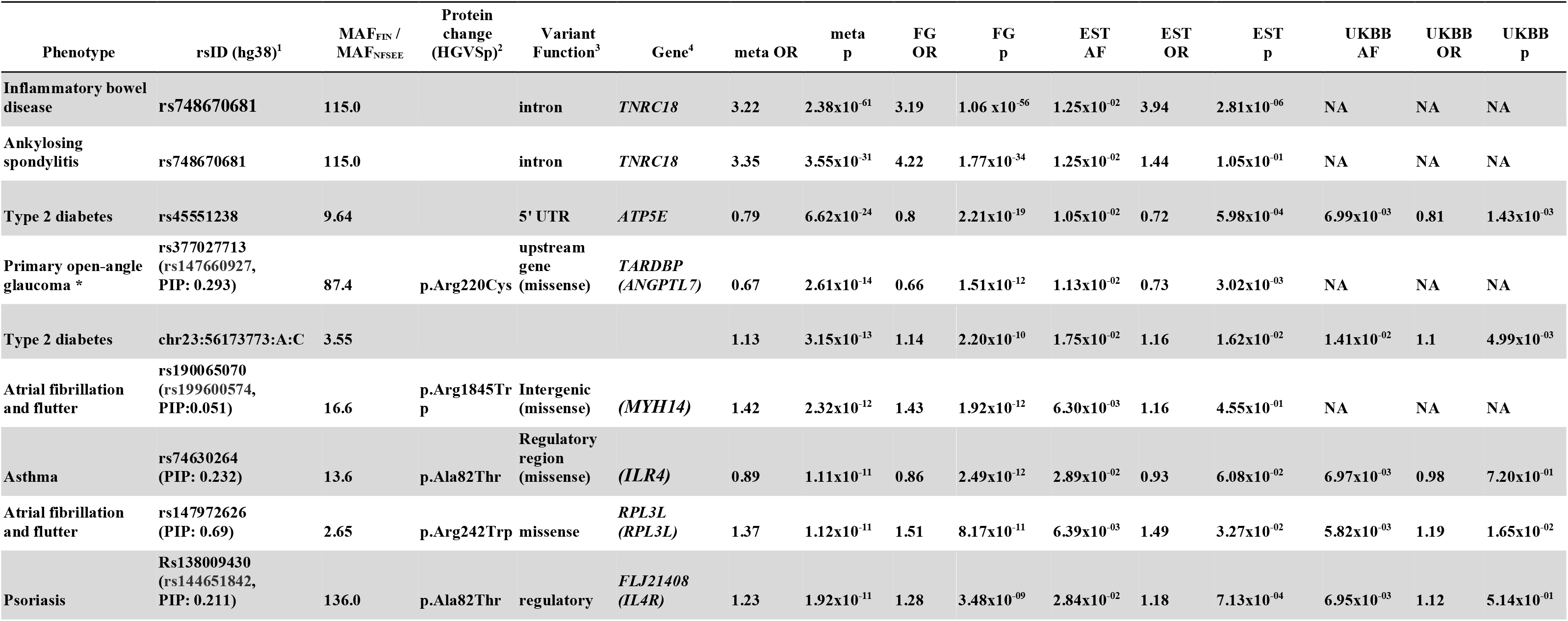
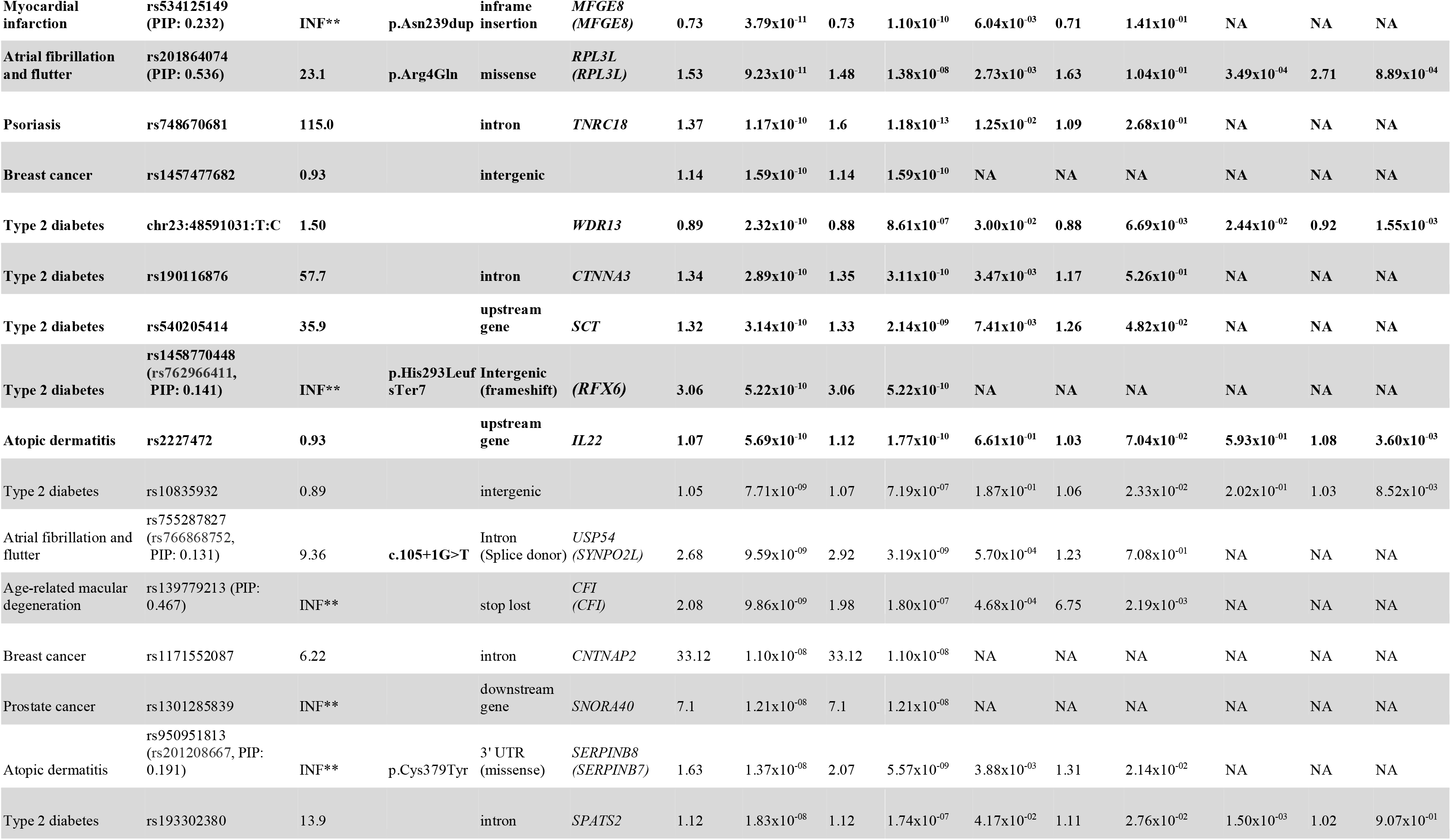
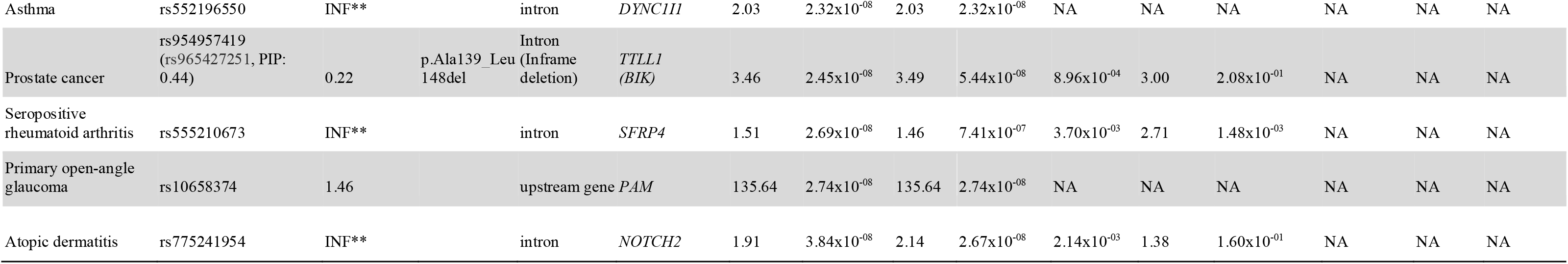
A total of 30 associations were identified in GWAS of 15 selected phenotypes that have not been reported in previous GWA studies. Table is ordered by meta-analysis p-values, in descending order of significance. All reported variants were mapped to GRCh 38. Rows which are in bold are variants surpassing Bonferroni multiple testing correction for 15 endpoints (p < 3.3 x 10-9) * We have previously published the ANGPTL7 variant association with glaucoma^19^, ** denotes values of infinity, resulting from **MAF_NFSEE_** being 0.00. ^1^ Coding variant RSID in PIP given in parenthesis if a coding variant was observed in the credible set. ^2^ HGVS notation protein coding change if either lead variant was coding or coding credible was observed in the credible set (if either one exists) ^3^ Coding variant consequence given in parenthesis in case lead variant was not a coding variant and a coding variant was observed in the credible set. ^4^ Gene corresponding to the variant function. In case a lead variant was not a coding variant, but there was a coding variant in the credible set, the credible set coding variant gene is given in parenthesis.

The identification of a novel signal for IBD mapping to a single variant in an intron of *TNRC18*, highlights the value of Finngen for discovery, even when the case sample size is far below that of existing meta-analyses. This variant has a strong risk increasing effect (AF 3.6%, OR 3.2, p-value 2.4*10^-61^) eclipsing the significance of signals at *IL23R*, *NOD2* and MHC. The variant is 114-fold enriched in Finns compared to NFSEE Europeans where the allele frequency is too low (0.04%) to have been identified in previous GWAS. We were however able to unequivocally replicate this association in the Estonian biobank (AF 1.3%, OR 3.9, p-value 2.8*10^-06^) owing to the relatively higher frequency in the genetically related Estonian population. This variant was also associated with risk to multiple other inflammatory conditions evaluated in FinnGen, including interstitial lung disease (OR 1.43, p 6.3*10^-26^), ankylosing spondylitis (OR 4.2, p 1.8*10^-34^), iridocyclitis (OR 2.3, p 1.2*10^-27^) and psoriasis (OR 1.6, p 1.1*10^-13^). However, the same allele appears protective for an endpoint combining multiple autoimmune diseases (https://r5.risteys.finngen.fi/phenocode/AUTOIMMUNE) (OR 0.84, p 6.2*10^-12^; [e.g. T1D OR 0.64,p 2.7*10^-7^, hypothyroidism OR 0.85, p 7.8*10^-7^]).

The highest number of (eight loci) novel and enriched low frequency associations were identified in type 2 diabetes, most likely due to the large number of T2D patients in FinnGen release 5 (29,193). Other noteworthy findings from this set of 30 novel findings for 15 well-studied diseases are described further in BOX 1.

### Coding variant associations

Motivated by the identification of high effect coding variant associations within the selected 15 diseases, we performed a phenome wide GWAS followed by fine-mapping in order to identify Finnish-enriched, putative causal coding variants.

In a GWAS of 1932 distinct endpoints and 116 387 711 variants (Supplementary Table 4, case overlap < 50% and n cases>80), we identified 2733 independent associations in 2492 loci across 807 endpoints (Supplementary table 9) at genome-wide significance threshold (p<5×10^-8^), and 893 signals in 771 loci across 247 endpoints after stringent Bonferroni correction for 1932 endpoints (p<2.6×10^-11^). HLA region was excluded here and pheWAS of imputed classical HLA gene alleles in FinnGen is reported by Ritari et al^12^.

After statistical fine-mapping, we observed a coding variant (missense, frameshift, canonical splice site, stop gained, stop lost, inframe deletion; PIP>0.05) in 369 associations (13.5% of all associations) spanning 202 endpoints. Full results with all 2803 endpoints (including endpoints with case overlap > 50% that are excluded here) are publicly available via a customized browser based on PheWeb^20^ code base (r5.finngen.fi) and as summary statistic files (https://www.finngen.fi/en/access_results).

To put the frequency spectrum and putative acting mechanisms in interpretable context, we chose a single most significant association per signal by LD based merging (r2>0.3 lead variants merged) resulting in 1838 unique associations in 681 endpoints (Supplementary Table 10). Applying stringent Bonferroni corrected p-value threshold (p<2.6×10^-11^) resulted in 493 associations in 112 endpoints. Although the majority of the 493 stringent unique associations were driven by common variants, 169 and 68 had a lead variant frequency of < 10% and < 1% respectively (Figure 3 A). We observed 87 (51.5%) of the 169 low frequency (MAF < 10%) lead variants to be over two-fold enriched in Finland compared to NFSEE.

**Figure 3.**
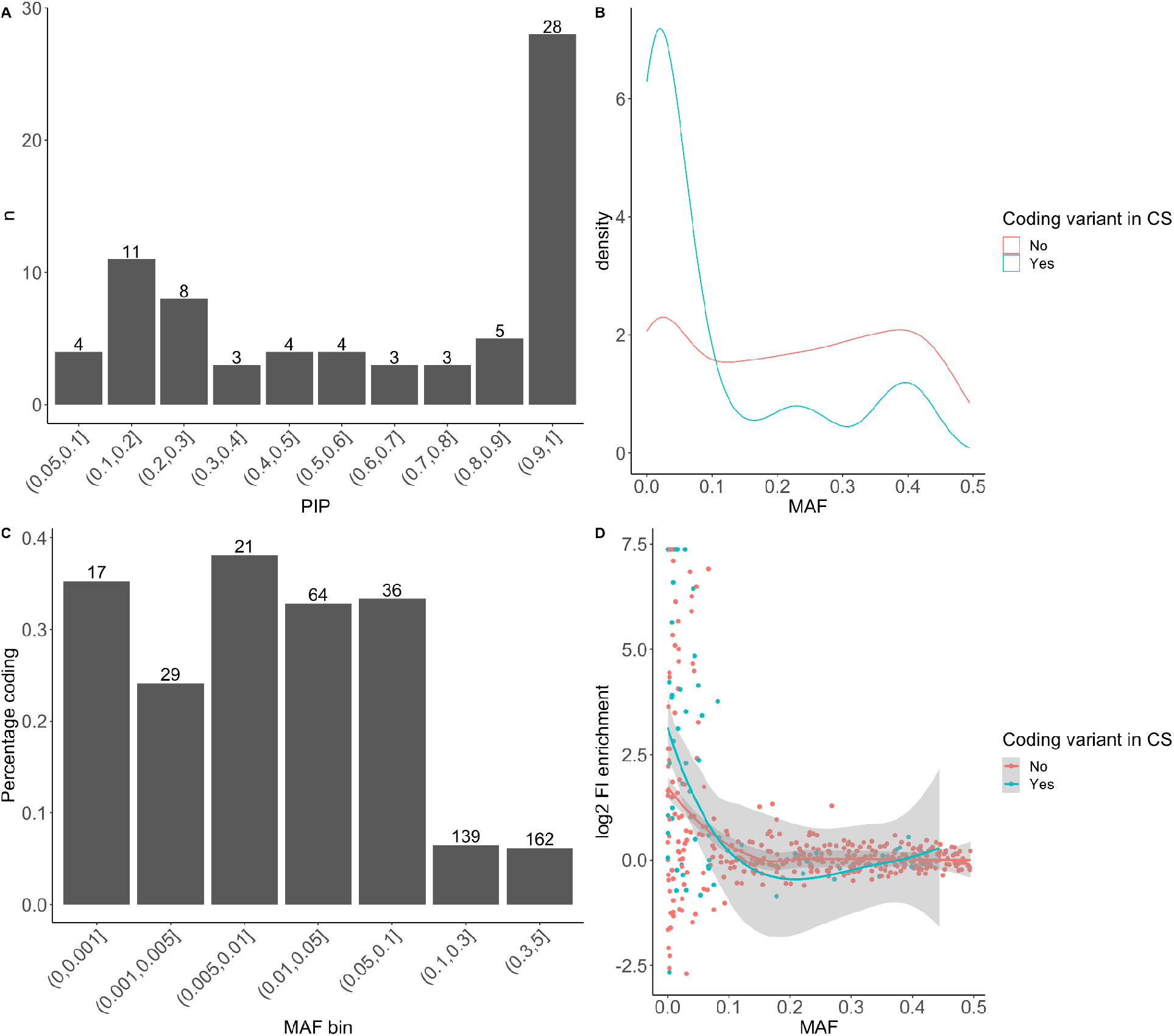
Characteristics of 493 (73 with coding variants in credible set) unique associations in 112 (42 endpoints with coding variants in credible set) endpoints identified in FinnGen release 5. Note that 25 of the associations with a coding variant with PIP<0.05 in credible sets removed from plots as “uncertain to contain coding variant”. A) Distribution of fine-mapping posterior inclusion probabilities (PIP) of the 73 coding variants B) Allele frequency spectrum in associations with and without coding variants in credible sets. C) Proportion of coding variants identified in different allele frequencies. The numbers above bars indicate the number of associations within a bin, the y-axis indicates the proportion of associations with coding variants in their credible sets. D) Enrichment in Finland as a function of allele frequency (enrichment value for variants with allele frequency of 0 in NFEE in GnomAD was set to maximum observed enrichment value of log2(166)=7.38).

After statistical fine-mapping of the 493 unique associations, we identified a coding variant (PIP>0.05) in 73 (14.8%) of the credible sets associated with 42 endpoints (Supplementary Table 10). The majority (43) of the fine-mapped coding variants had PIP>0.5 and 28 had PIP>0.9 (Figure 3 A). The highest proportion and the majority (54/73) of associated coding variants had MAF < 10% (Figure 3 B,C). The coding variant associations were more enriched in Finland than non-coding associations in associations driven by variants with AF < 10% (Figure 3 D, Wilcoxon rank sum test p 2.1 * 10^-4^). For example, we observed a coding variant more often in the credible sets of > 2 times enriched associations (34/89, 38.2%) than non-enriched associations (19/80, 23.8%) among the lower frequency (MAF<10%) associations. The higher proportion of coding variants in>2 times enriched variants persisted when increasing PIP threshold to 0.2 (Enriched 30/82(36.6%) and non-enriched 14/74(18.9%)).

Fine-mapping properties and replicability across diverse biobanks (FinnGen, Biobank Japan and UKBB) are explored in detail in accompanying manuscripts ^15, 16^ and in Sun et al^21^.

We next wanted to quantify the benefits of Finnish population isolate in GWAS discovery. To this end, we assessed if Finnish enriched lower frequency (MAF<10%) variants were more likely to be associated with a phenotype than would be expected by chance. We randomly sampled 1,000,000 times the number of genome-wide significant variants observed (169) from a set of frequency matched variants (MAF<10%) that were not associated with any endpoint (p>0.001). Only six out of 1 million random draws had a higher proportion of two-fold Finnish enriched variants than was observed in the significant associations (51.5% observed vs. 37% expected; p-value 2.8*10^-5^).

### Known rare and low-frequency pathogenic variant associations

Among the genome-wide significant coding variant associations, we identified 13 variant associations (AF range 0.04%-2%) classified as Pathogenic/Likely pathogenic in ClinVar (Supplementary Table 8). Several of these variants are highly enriched in Finland, such as rare frameshift variant at *NPHS1* associated with nephrotic syndrome, including the congenital form (ICD-10: N04,p.Leu41fs, AF FinnGen 0.9%; gnomAD NFSEE 0.009%, OR 185, p 4.3×10^-27^). Congenital nephrotic syndrome of Finnish type is a recessively inherited rare disease belonging to the Finnish Disease heritage^8^. Other known pathogenic variants in ClinVar include a missense variant in *CERKL* associated with hereditary retinal dystrophy (p.Cys125Trp, AF FinnGen 0.6%; gnomAD NFSEE 0%, OR 98716, p 5.15 * 10^-25^), a missense variant in *XPA* associated with non-melanoma neoplasm of skin (*“other malignant neoplasm of skin”*) (p.Arg228Ter, AF FinnGen 0.02%; GnomAD NFSEE 0%. OR 4.4 p 8.3*10^-18^) and above-mentioned frameshift variant in *PALB2* associated with breast cancer (p.Leu531fs *“malignant neoplasm of breast”*) (p.Ala82Pro, AF FinnGen 0.2%, gnomad NFSEE 0%, OR 28.8, p 3.7*10^-33^). These associations demonstrate that imputation using a population specific genotyping array and imputation panel combined with national registry-based phenotyping in the isolated Finnish population can successfully identify associations and fine-map causal variants even in rare variants and phenotypes. An extended study of ClinVar variants as well as variants with novel bi-allelic Mendelian effects in FinnGen is described in an accompanying paper by Heyne et al^22^.

### Variant associations in known disease genes

In the remaining genome-wide significant coding variant associations not reported as pathogenic in ClinVar, 91 had allele frequency < 10%. Of the 91 variants, 44 were over five times more common in Finland than NFSEE and 17 had not been observed at all in NFSEE. (Table 2). Several of the 17 variants are in a gene where other variants are pathogenic for related traits. These FinnGen associations include *RFX6* frameshift variant associated with T2D (p.His293LeufsTer7, AF 0.15%, OR 3.7, p 1.2 * 10^-10^, Clinvar: Monogenic diabetes and others), *SCN4A* associated with “Myotonic diseases” (AF 0.07% p 3.3 * 10^-14^, ClinVar: “Paramyotonia congenita” and others), missense in *MYH14* associated with “Sensorineural hearing loss” (p.Ala1156Ser AF 0.04%, OR 19.9, p 1 * 10^-15^, Clinvar: “Non syndromic hearing loss” and others) and a stop gained variant in *TG* associated with autoimmune hypothyroidism (p.Gln655Ter, AF 0.1%, OR 3.2, p 3.9*10^-11^). These variants in *RFX6, SCN4A and TG* have been observed previously in Finnish cohorts^23–25^ but had uncertain significance (single carrier in *TG*) or conflicting interpretation (*SCN4A)* in ClinVar. Pathogenic variants in *RFX6* cause Mitchell-Riley syndrome with recessive inheritance (characterized by neonatal diabetes), however, heterozygote enrichment of *RFX6* truncating variants have been observed in maturity-onset diabetes of the young^23^ where the same variant observed here was identified in a Finnish replication.

**Table 2.**
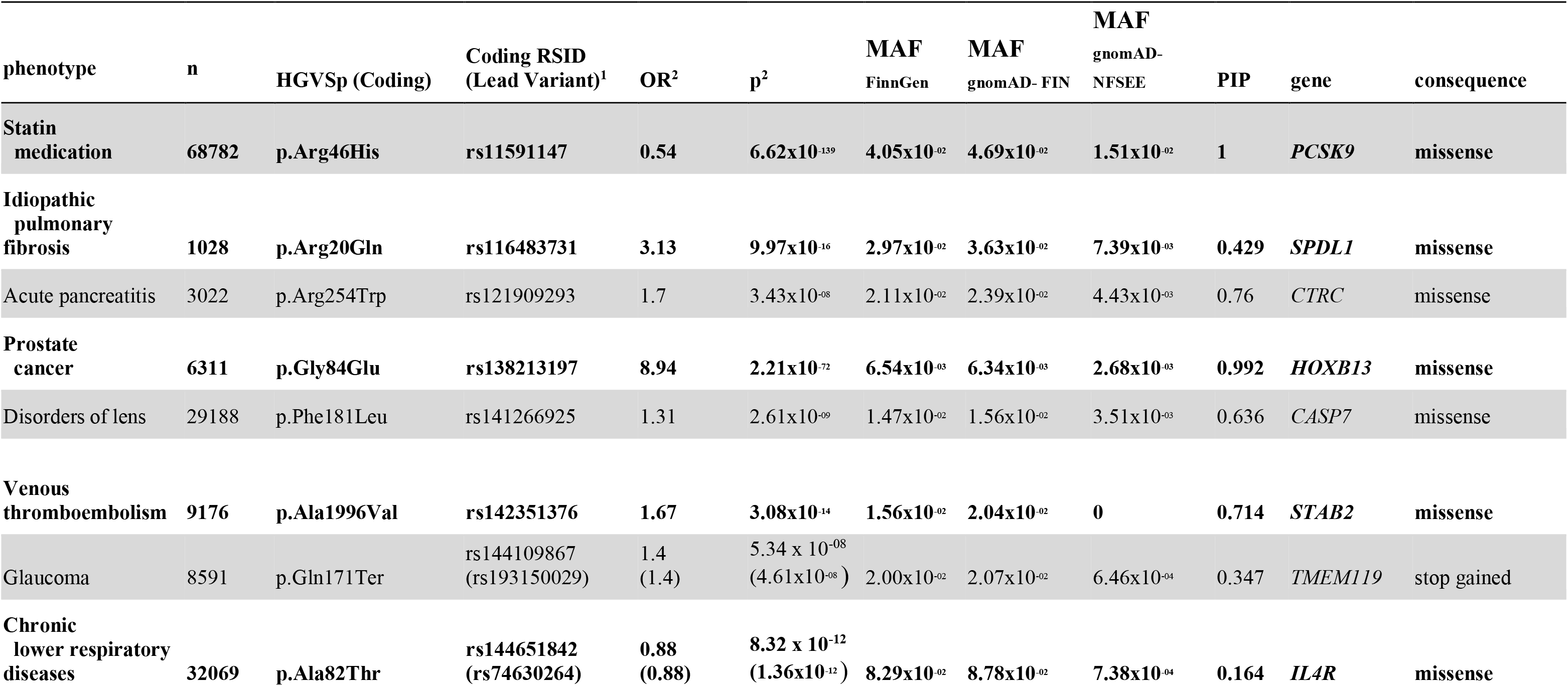
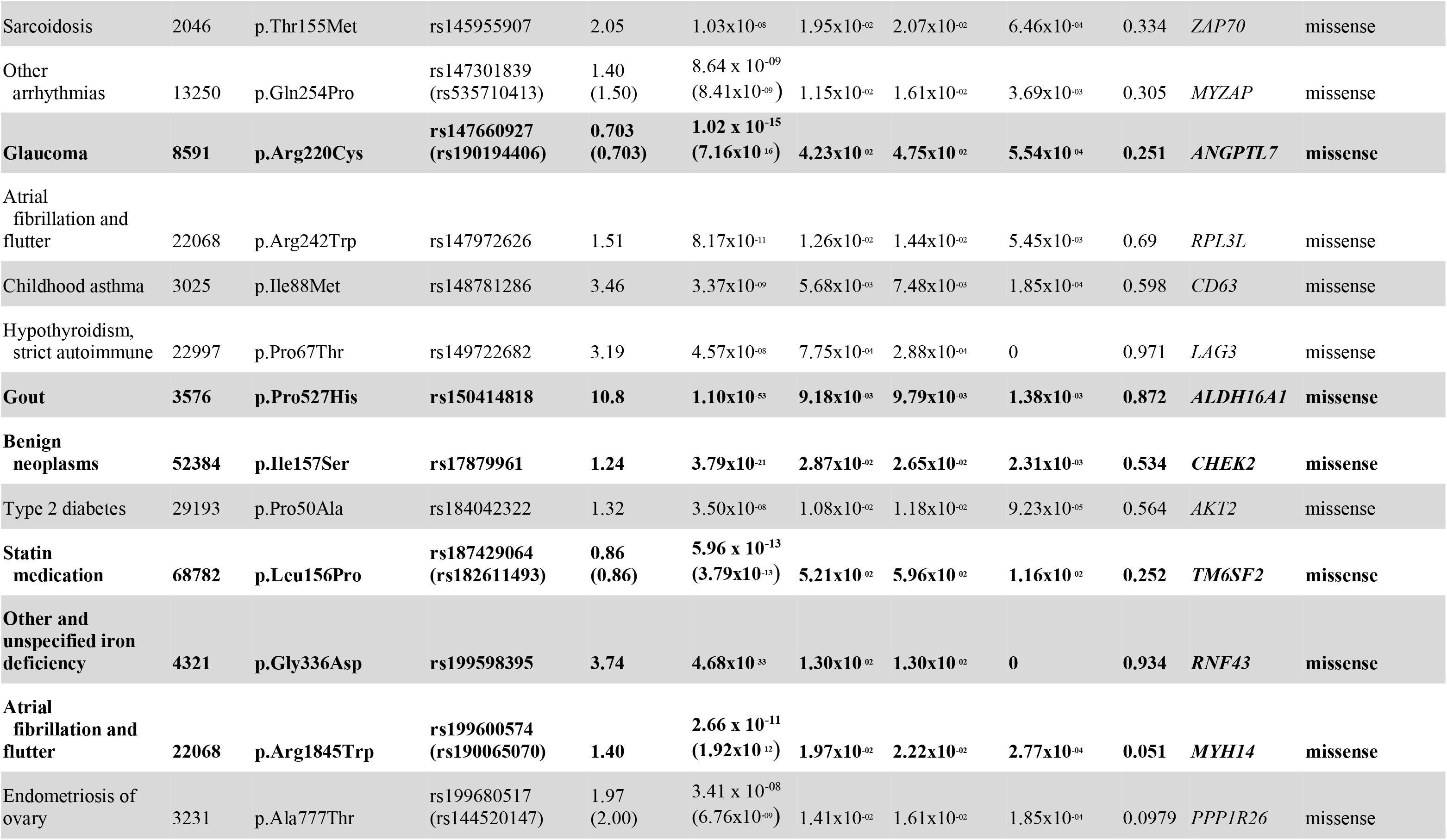
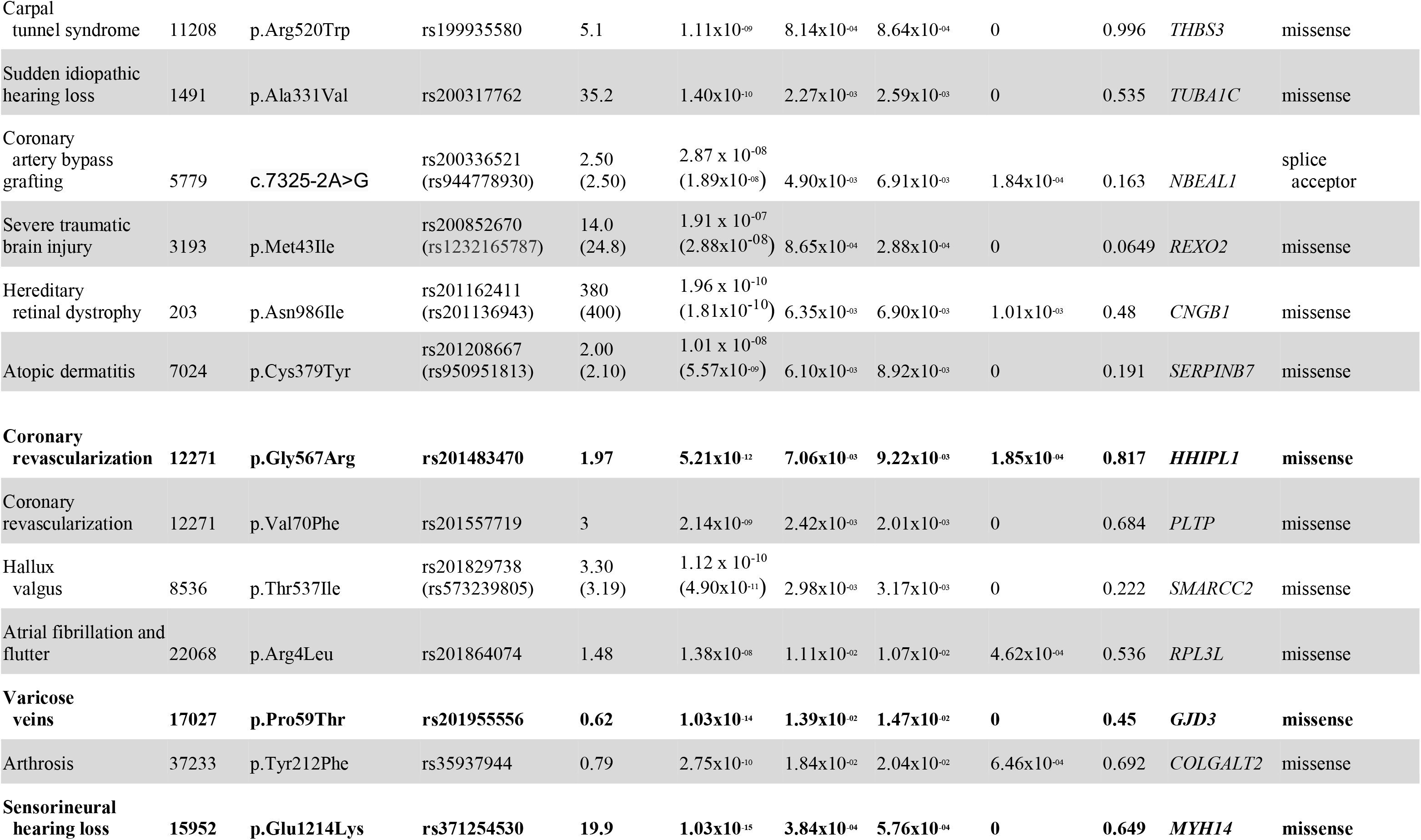
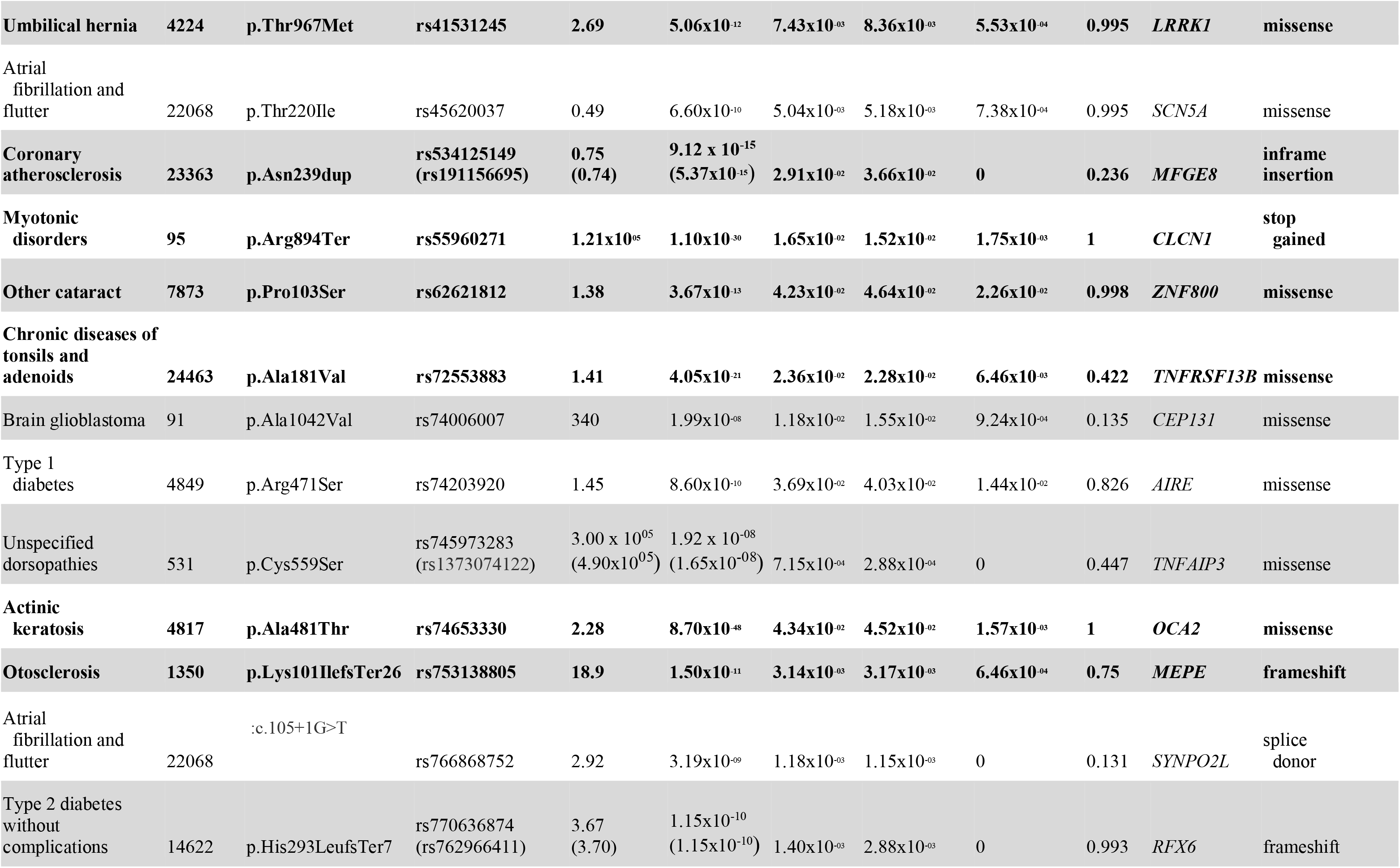
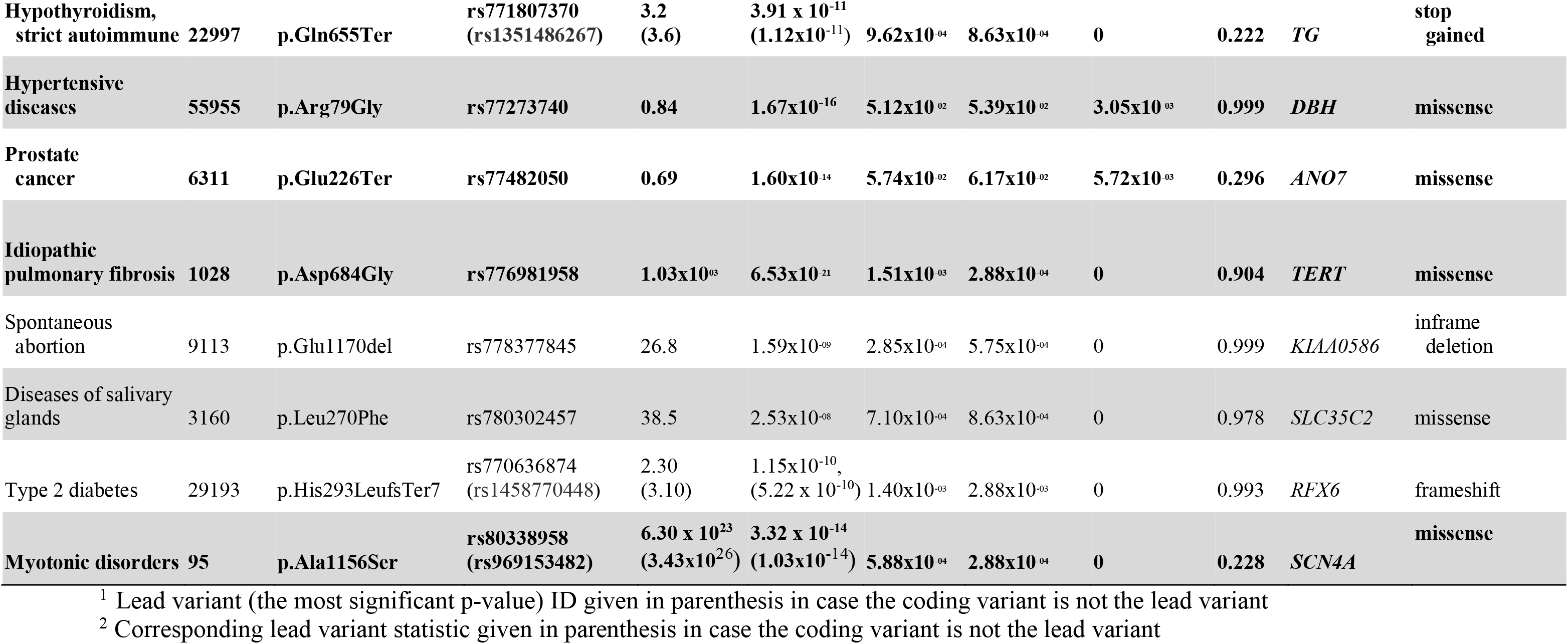
Coding variants enriched over two-fold in Finns over Non-Finnish-Swedish-Estonian European populations in gnomAD that are significantly associated in GWAS in 1,932 FinnGen phenotypes and not classified as pathogenic/likely pathogenic in ClinVar. The strongest association (lowest p-value) reported as single putative causal coding variant and associations in LD are clustered together as putatively sharing a causal variant. Rows which are in **bold** are variants surpassing Bonferroni multiple testing correction for 1,932 endpoints (p < 2.6 x 10^-11^) and arranged in descending order. PIP = posterior inclusion probability.

### New disease associations

Among the other genome-wide significant coding variant associations without prior associations in GWAS catalog (in LD) or ClinVar (same gene) we observed five coding variants where AF in gnomAD NFSEE was zero (Table 2). A missense variant in *STAB*2 (p.Ala1996Val) was associated with venous thromboembolism (VTE, n=9176) (AF 1.6%, OR 1.6, p 3.1 * 10^-14^). *STAB2* has been recently suggested as a new candidate risk gene for VTE, based on enrichment of rare functional variants in exome sequencing of 393 VTE cases and 6114 controls^26^ and our results strongly support this. Another association with no copies in NFSEE was an association of a missense variant (p.Val70Phe, AF 0.2%, OR 3.0, p 2.1*10-9) in *PLTP* and coronary revascularization (n=12271, coronary angioplasty or bypass grafting). *PLTP* is one of lipid transfer proteins in human plasma transferring phospholipids from triglyceride-rich lipoproteins to high-density lipoprotein (HDL) and its activity is associated with atherogenesis in humans and mice^27^. Common and low frequency intergenic variants near *PLTP* independent of p.Val70Phe are associated with lipid levels (HDL, triglycerides)^28, 29^ and coronary artery disease^30^. Our results support *PLTP* being the causal gene for symptomatic atherosclerosis in this locus.

A missense variant in *TUBA1*C (p.Ala331Val, AF 0.2%, OR 35.2, p 1.4*10^-10^) was associated with sudden idiopathic hearing loss (n=1,491). No relevant phenotype has been reported for variants in *TUBA1C* and it is the first reported GWAS locus for sudden idiopathic hearing loss. *TUBA1C* codes for an α-tubulin isotype. The precise roles of α-tubulin isotypes are not known, but mutations in other tubulins can cause various neurodevelopmental disorders^31^. p.Ala331Val was also associated with vestibular neuritis (inflammation of the vestibular nerve, n=1,224, OR = 40.9, p-value= 3.2 * 10^-10^). Pure vestibular neuritis presents acutely with vertigo but not hearing loss and accurate diagnosis of vertigo in acute settings is challenging and misdiagnosis is possible^32^.

We report several of these coding associations in separate manuscripts. One such novel observation is a missense variant (p.Arg20Gln, AF 3%, gnomAD NFEE 0.7%) in *SPDL1* with pleiotropic association with strongly increased risk of idiopathic pulmonary fibrosis (OR 3.1, p 1.0 * 10^-15^) but protective with endpoint combining all cancers (OR 0.82, p 2.1 * 10^-15^)^33^. Other associations described in separate manuscripts are associations between in frame deletion in *MFGE8* and coronary atherosclerosis (p.Asn239dup, AF 2.9%, gnomAD NFSEE 0%, OR 0.74, p 5.4*10^-15^)^34^, a frameshift variant in *MEPE* (p.Lys101IlefsTer26, AF 0.3%, gnomAD NFSEE 0.07%, OR 18.9, p 1.5 *10^-11^) and otosclerosis^35^, and missense variant in *ANGPTL7* and glaucoma (p.Arg220Cys, AF 4.2%, gnomAD NFSEE 0.06%, OR 0.7, p 7.2*10^-16^)^19^

### Coding variants associated with medication use

An exceptional registry available in FinnGen is a prescription medication purchase registry (KELA, Table 1) linking all prescription medication purchases for all FinnGen participants since 1995. Using prescription records from this registry, we identified two enriched low-frequency coding variants that were associated with drug purchase of statin medications (3 or more purchases per individual) (Table 2). A missense variant in *TM6SF2* (p.Leu156Pro*, rs187429064)* was associated with decreased likelihood of being prescribed statins (AF 5.2%, gnomAD NFSEE 1.2%, OR 0.86, p-value 3.8 * 10^-13^) but with an increased likelihood for insulin medication for diabetes (OR 1.17, 8.2 * 10^-11^) and T2D (OR 1.15, p-value 2.6 * 10^-8^). In addition, the same variant showed strong association with strongly increased risk of hepatocellular carcinoma (ICD10 C22 “hepatic and bile duct cancer”) (OR 3.7, p-value 5.9 * 10^-10^). Consistent with decreasing the likelihood of being prescribed statins, *TM6SF2* p.Leu156Pro and another independent (r2 0.003) missense variant (p.Gly167Lys, rs58542926) have previously been associated with decreased LDL and total cholesterol (TC) levels^36^. In a mouse model, both p.Gly167Lys and Leu156Pro lead to increased protein turnover and reduced cellular protein levels of TM6SF2^37^. TM6SF2 p.Gly167Lys decreases hepatic large VLDL particle secretion and increases intracellular lipid accumulation^38, 39^, which likely explain its associations with non-alcoholic fatty liver disease^40^, alcohol related cirrhosis^41^, hepatocellular carcinoma^42^ and incident T2D^43^. Our results provide, in a single pheWAS analysis, strong evidence of novel p.Leu156Pro having similar consequences of decreases in circulating lipid levels and increasing risk of diabetes, cirrhosis and liver cancer, as observed for p.Gly167Lys. Such a variant pleiotropy can be explored in phewas view in custom PheWas view in custom PheWeb^20^ browser (http://r5.finngen.fi/variant/19-19269704-A-G)

## Conclusions

In this paper and accompanying publications, we present FinnGen, one of the largest nationwide genetic studies with access to comprehensive electronic health register data of all participants. The final aim of the study is to collect 500,000 biobank participants by the end of 2023. The interim releases of FinnGen have already contributed to many new discoveries and insights into human genetic variation and how it affects disease and health^19, 44–48^, including contributions to COVID-19 host genetics initiative^49^ and global biobank meta-analysis initiative^50^. Summary statistics from each data release will be made publicly available after a one year embargo period and all summary statistics described here are freely available. (www.finngen.fi/en/access_results).

An important feature of FinnGen compared to other similar projects, such as the UKBB^10^, is the unique genetic makeup of the Finnish population. In the GWAS of selected, well-studied diseases, we were able to identify several novel associations with a fraction of the cases compared to the largest published GWA studies. These associations were, as expected, largely observed with variants that were increased in frequency in the Finnish population bottleneck and would have required prohibitively large sample sizes in older, non-bottlenecked populations (Figure 2D). Moreover, in the GWAS of 1,932 endpoints, we observed that over two-fold Finnish enriched variants were 1.6 times more likely to be associated with a phenotype than would be expected by chance.

Further, we observed that putative coding variant associations were not only of lower allele frequency but also more often enriched in Finland than non-coding variant associations (Figure 3). This observation is expected as coding variant associations are more deleterious on average and selection drives the allele frequencies down. However, some of these deleterious alleles survived the bottleneck and increased in frequency, facilitating the identification of their associations with diseases.

Imputation with a population specific imputation panel provides high imputation accuracy down to very low allele frequencies (Supplementary Figure 5) enabling identification of association with low frequency variants using GWAS approach, instead of direct sequencing. This high imputation accuracy combined with broad population registry-based phenotyping facilitates identification of very low frequency variants associated with rare phenotypes, which have largely been missed in the majority of GWA studies published to date^51^. We demonstrated this by identifying known ClinVar variant associations with diseases such as the Finnish congenital nephrotic syndrome or polycystic liver disease. Furthermore, we uncovered novel low frequency variant associations with common and rare phenotypes, including clinically challenging but not well genetically studied sudden idiopathic hearing loss or Carpal tunnel syndrome. The recently reported Finnish enriched Gln175His variant in the ANGPT7 gene, protective for glaucoma, is also an example of the bottleneck effect in disease associated variant discovery^19^.

The university hospital-based recruitment, coupled with legacy case cohorts of several diseases is another feature of FinnGen. This strategy captures cases in many disease areas and distinguishes it from many working age population cohorts. For example, in UKBB, where the recruitment was based on postal invitation to individuals aged 40-69 and living within 40 km (25 miles) of one of the assessment centers^52^, the participants are likely to be healthier than in hospital based collections. The approach in FinnGen has advantages and disadvantages. For many disease-focused studies, it provides a higher number of cases and a relatively economical way of recruiting a large sample within a feasible time frame. For example, in the 15 common diseases studied in this paper, the sample prevalence in FinnGen was higher than in UKBB. The difference was the most extreme in Alzheimer’s disease (2.7% in FinnGen vs. 0.2% in UKBB), a disease of old age, and the most similar in asthma (9.4% vs. 7.4%) (Figure 2A). FinnGen also has a relatively high sample prevalence of some severe mental disorder cases such as schizophrenia (2.5%, n=5,562) and bipolar disease (2.1%, n=4,501) that are often underrepresented in biobank studies. A key aspect of the recruitment strategy is the Finnish biobank legislation that enables participants to donate samples with a broad consent to medical research in general. This makes recruitment cost-effective as the same samples and data can be used, after appropriate application steps, for many medical research studies. However, due to the recruitment strategy, FinnGen is not epidemiologically representative.

In conclusion, FinnGen as a large-scale biobank study with unique features of the Nordic health care system and population structure provides opportunities for a wide range of genetic discoveries. These include identification of disease associated coding variants, identification of variant pleiotropy and longitudinal analysis of disease trajectories. Combining results with other large scale biobank projects can further improve our understanding of the role of genetic variation in health and disease, especially in genetically understudied diseases.

## Methods

### Biobank samples

The FinnGen study (https://www.finngen.fi/en) is an on-going research project that utilizes samples from a nationwide network of Finnish biobanks and digital health care data from national health registers. FinnGen aims to produce genomic data with linkage to health register data of 500,000 biobank participants. Samples in the FinnGen study include legacy samples (prospected number 200,000) from previous research cohorts (often disease-specific) that have been transferred to the Finnish biobanks, and prospective samples (prospected number 300,000) collected by biobanks across Finland. Prospective samples from six regional hospital biobanks represent a vast variety of patients enrolled in specialized health care, samples from a private health care biobank allow for enrichment of the FinnGen cohort with patients underrepresented in specialized health care, while participants recruited through Blood Service Biobank enrich the cohort with healthier individuals. Samples have not specifically been collected for FinnGen but the study has incorporated all that have been available in the biobanks (see Supplementary Methods for details). In the present study, we included samples from 224,737 biobank participants.

### Phenotyping

Registry data on all FinnGen participants was collected and processed from different national health registers, including hospital and outpatient visits in HILMO - Care Register for Health Care (inpatient and outpatient primary and secondary diagnoses: ICD 8,9,10; operations: NOMESCO Classification of Surgical Procedures, Hospital League surgical procedure codes), AvoHILMO - Register of Primary Health Care (main and side diagnosis using ICD-10 and ICPC2 codes, operations/procedures using NOMESCO and national SPAT codes), Causes of Death (immediate, underlying and contributing causes of death on the death certificate with ICD-8,9,10 codes), reimbursed medication entitlements and prescribed medicine purchases (specific Social Insurance Institution of Finland reimbursement codes and Anatomical Therapeutic Chemical (ATC) codes, respectively), and the Finnish Cancer Registry (FCR; using ICD-O-3 codes). Pseudonymized register data was combined with the minimum phenotype dataset from the Finnish biobanks (age, sex, year of sampling, height, weight and smoking). Clinical endpoints were constructed from the register codes using Finnish version of International Classification of Diseases, 10th revision (ICD-10) diagnosis codes and harmonizing those with definitions from ICD-8 and ICD-9. Finnish ICD version is mostly identical to international ICD classification but has minor modifications, e.g.. there are additions to certain disease classifications in 4^th^ and 5^th^ character level to add specificity. When relevant, the information on reimbursed medication and/or prescription medicine purchases and operations augmented the endpoint data. Cancer endpoints were constructed based on the Finnish Cancer Registry and Causes of Death data. The definitions of FinnGen disease endpoints and their respective controls for each release are available at: https://www.finngen.fi/en/researchers/clinical-endpoints and FinnGen endpoints can also be browsed at https://r5.risteys.finngen.fi/. See Supplementary Methods Section 1 for further details.

Some of the endpoints have a high number of overlapping cases and to avoid reporting highly repetitive endpoints, we clustered all endpoints if there was an overlap of >50% of cases between them and chose the one with the most genome-wide significant hits. On a few occasions a manual choice was made to select the most representative endpoint among the correlating endpoints. After clustering, we had 1932 endpoints for the main GWAS analysis.

### Genotyping and QC

Samples were genotyped with Illumina (Illumina Inc., San Diego, CA, USA) and Affymetrix arrays (Thermo Fisher Scientific, Santa Clara, CA, USA). Genotype calls were made with GenCall and zCall algorithms for Illumina and AxiomGT1 algorithm for Affymetrix data. Chip genotyping data produced with previous chip platforms and reference genome builds were lifted over to build version 38 (GRCh38/hg38) following the protocol described here: dx.doi.org/10.17504/protocols.io.nqtddwn. In sample-wise QC, individuals with ambiguous sex, high genotype missingness (>5%), excess heterozygosity (+-4 standard deviations). In variant-wise QCvariants with high missingness (> 2%), low Hardy–Weinberg Equilibrium (HWE) P-value (< 1e-6) and minor allele count, minor allele count (MAC) < 3 were removed. Chip-genotyped samples were pre-phased with Eagle 2.3.5 (https://data.broadinstitute.org/alkesgroup/Eagle/) with the default parameters, except the number of conditioning haplotypes was set to 20,000.

### Genotype imputation with a population-specific reference panel

The population-specific Sequencing Initiative Suomi (SISu) v3 imputation reference panel was developed by using high-coverage (25-30x) whole-genome sequencing (WGS) data for 3,775 Finnish individuals. Briefly, the variant callset was produced with GATK HaplotypeCaller algorithm by following GATK best-practices for variant calling. Genotype-, sample- and variant-wise QC was performed using the Hail framework (https://github.com/hail-is/hail) v0.1 and the resulting high-quality WGS data were phased (see Supplementary Methods). Genotype imputation was carried out by using the SISu v3 reference panel with Beagle 4.1 (version 08Jun17.d8b, https://faculty.washington.edu/browning/beagle/b4_1.html) as described in the following protocol: dx.doi.org/10.17504/protocols.io.nmndc5e. Post-imputation QC involved non-reference concordance analyses, checking expected conformity of the imputation INFO-values distribution, MAF differences between the target dataset and the imputation reference panel and checking chromosomal continuity of the imputed genotype calls. After these steps, variants with imputation INFO score < 0.6 or MAF < 0.0001 were excluded.

### Association analysis and fine-mapping

Mixed model logistic regression method SAIGE (version 0.35.8.8)^53^, was used for association analysis. We used sex, age, genotyping batch and 10 PCs as covariates (see extended methods for details). We used SuSiE^54^ for fine-mapping. We finemapped all regions with variants with p-value< 10^-6^ and extended regions 1.5MB upstream and downstream from each lead variant. Finally overlapping regions were merged and subjected to fine-mapping. Major histocompatibility region (chr 6: 25–36 Mb) was excluded due to complex LD structure. We allowed up to 10 causal variants per region and SuSiE reports a 95% credible set for each independent signal. As LD, we used in-sample dosages (i.e cases and controls used for each phenotype) computed with LDStore2. FinnGen Fine-mapping pipeline is available in GitHub (https://github.com/FINNGEN/finemapping-pipeline).

To define independent signals within a locus, we utilized fine-mapping results. For each locus we report credible set as an independent hit if it represents primary strongest signal with lead p-value < 5*10^-8^ and for secondary hits we require genome-wide significance and log bayes factor (BF)>2. The BF filtering was necessary due to SuSiE sometimes reporting multiple credible sets for a single very strong signal but this is indicated in SuSiE as low BF (the model does not improve by adding another signal in the region i.e. not independent signal).

### Estimation of expected number of enriched variant associations

We aimed to estimate if we observed more over two-fold Finnish enriched variant associations in lower frequency range (MAF< 10%) than would be expected by chance. To this end, we sampled a subset of variants (MAF<10%) that were not associated with any endpoint in FinnGen (p>0.001). We drew 1 million random samples of the number of independent hits (169) observed in GWAS from the set of non-associated variants. In order to closely follow the observed frequency distribution, we further matched the random samples to contain the same number of variants in each frequency bin ((0,0.001],(0.001,0.005],(0.005,0.01] and then in 0.0 bins up to 0.1). We computed mean and standard deviation of percent two-fold enriched variants from the random samples and calculated p-value from normal distribution using the randomized mean and standard deviation.

### Estonian Biobank and UKBB replication

Estonian Biobank (EstBB) is a population-based biobank at the Institute of Genomics, University of Tartu. The current cohort size is 200,000 individuals (aged ≥ 18), reflecting the age, sex and geographical distribution of the adult Estonian population. Estonian samples represent 83%, Russians 14%, and other ethnicities 3% of all participants. All subjects have been recruited by general practitioners (GP), physicians in hospitals and during promotional events. Upon recruitment, all participants completed a questionnaire about their health status, lifestyle and diet. More specifically, the questionnaire included personal data (place of birth, place(s) of living, nationality etc.), genealogical data (family history of medical conditions spanning four generations), educational and occupational history, lifestyle data (physical activity, dietary habits - FFQ, smoking, alcohol consumption, women’s health, quality of life). The EstBB database is linked with national registries (such as Cancer Registry and Causes of Death Registry), hospital databases, and the database of the national health insurance fund, which holds treatment and procedure service bills. Diseases and health problems are recorded as ICD-10 codes and prescribed medicine according to the ATC classification. These health data are continuously updated through periodical linking to national electronic databases and registries. All participants were genotyped with genome-wide chip arrays and further imputed with a population-specific imputation panel consisting of 2,244 high-coverage (30x) whole genome sequenced individuals and 16,271,975 high-quality variants^55^. Estonian biobank ran association analysis of the 15 phenotypes (See Supplementary Table 8) used in this study in 136,724 individuals. Association analysis was conducted with SAIGE^53^ mixed models with age, sex and 10 PCs used as covariates.

We used the Pan UKBB (https://pan.ukbb.broadinstitute.org/) project European subset association analysis summary statistics in UKBB replication^56^ (See Supplementary Table 6).

As both Estonian biobank and UKBB are on human genome build 37, we lifted over the coordinates to build 38 to match FinnGen. Variants were then matched based on chromosome, position, reference and alternative alleles.

Inverse variance weighted meta-analysis was used to meta-analyse the three cohorts (code available https://github.com/FINNGEN/META_ANALYSIS).

### Variant annotation

We utilized Variant Effect Predictor (VEP; https://www.ensembl.org/info/docs/tools/vep/index.html) for annotating imputation panel variants. For coding variants, we chose a single most severe consequence and corresponding gene among canonical transcripts. We considered stop_gained, frameshift_variant, splice_donor, splice_acceptor, missense_variant, start_lost, stop_lost, inframe_insertion, and inframe_deletion as coding variants. We executed the variant annotation with Hail^57^.

### Colocalization

We applied colocalization to all fine-mapped regions. As a colocalization approach we used the probabilistic model for integrating GWAS and eQTL data presented in eCAVIAR (Hormozdiari et al. 2016). Given the posterior inclusion probabilities (PIP) of each phenotype in a region of interest, we calculated the colocalization posterior probability (CLPP). In contrast to eCAVIAR, we used SuSiE (Wang et al. 2019) to estimate the posterior inclusion probabilities.

For a pair of phenotypes we search for intersection of variants between their credible sets *CS_k_*, *k* = 1,…*K*.

*CLPP_k_* = ∑*_i in CSk_ p*1*_i_*・*p*1*_i_*, where p1 and p2 are the PIPs from phenotype 1 and 2.

We performed colocalization between FinnGen endpoints, eQTL catalogue^18^ as well as selected 36 continuous endpoints and 57 biomarkers from UKBB^16^. eQTL catalogue and UKBB traits were processed with a functionally equivalent fine-mapping pipeline to FinnGen.

### Automatic annotation of known GWAS hits

In order to identify novel hits from the GWAS results, we compared the fine-mapped results against genome-wide significant hits (p<5*10^-8^) in the GWAS Catalog association database^58^ and manually curated genome-wide significant hits from large GWA studies (Table 1). We checked and reported separately 1) matches in credible set variants and 2) matches with any variants in LD with a lead variant (highest PIP) after fine-mapping. LD lookup variants were chosen with the following criteria: 1) They were less than 1500 kb away from the lead variant, 2) They had a p-value < 0.01, 3) Their LD squared Pearson’s correlation with the lead variant was higher than a dynamic LD threshold based on lead variants p-value so that the expected p-value of linked variant would be nominally significant (r^2=5/inverse chi-squared survival function (p-value)).

A variant was considered to be already associated if its chromosome and position were identical to the GWAS Catalog association, and if its reference and alternate allele matched the strand- and effect-aligned association alleles. Because the GWAS Catalog associations do not have complete allele information, the associations’ allele information was retrieved from dbSNP data, human genome build 153, assembly 38. The GWAS Catalog version used was released on Apr 21st 2021.

### Ethics statement

Patients and control subjects in FinnGen provided informed consent for biobank research, based on the Finnish Biobank Act. Alternatively, separate research cohorts, collected prior the Finnish Biobank Act came into effect (in September 2013) and start of FinnGen (August 2017), were collected based on study-specific consents and later transferred to the Finnish biobanks after approval by Fimea, the National Supervisory Authority for Welfare and Health. Recruitment protocols followed the biobank protocols approved by Fimea. The Coordinating Ethics Committee of the Hospital District of Helsinki and Uusimaa (HUS) approved the FinnGen study protocol Nr HUS/990/2017.

The FinnGen study is approved by Finnish Institute for Health and Welfare (THL), approval number THL/2031/6.02.00/2017, amendments THL/1101/5.05.00/2017, THL/341/6.02.00/2018, THL/2222/6.02.00/2018, THL/283/6.02.00/2019, THL/1721/5.05.00/2019, Digital and population data service agency VRK43431/2017-3, VRK/6909/2018-3, VRK/4415/2019-3 the Social Insurance Institution (KELA) KELA 58/522/2017, KELA 131/522/2018, KELA 70/522/2019, KELA 98/522/2019, and Statistics Finland TK-53-1041-17.

The Biobank Access Decisions for FinnGen samples and data utilized in FinnGen Data Freeze 5 include: THL Biobank BB2017_55, BB2017_111, BB2018_19, BB_2018_34, BB_2018_67, BB2018_71, BB2019_7, BB2019_8, BB2019_26, Finnish Red Cross Blood Service Biobank 7.12.2017, Helsinki Biobank HUS/359/2017, Auria Biobank AB17-5154, Biobank Borealis of Northern Finland_2017_1013, Biobank of Eastern Finland 1186/2018, Finnish Clinical Biobank Tampere MH0004, Central Finland Biobank 1-2017, and Terveystalo Biobank STB 2018001.

### Data availability statement

Summary statistics from each data release will be made publicly available after a one-year embargo period and all summary statistics described here are freely available. (www.finngen.fi/en/access_results).

### Code availability statement

Central data analysis/processing pipelines used are freely available: FineMapping pipeline (https://github.com/FINNGEN/finemapping-pipeline), Meta-analysis (https://github.com/FINNGEN/META_ANALYSIS), genetic ancestry and PCA pipeline (https://github.com/FINNGEN/pca_kinship).

## Supporting information

FinnGen banner author list.

## Data Availability

all summary statistics described here are freely available.(www.finngen.fi/en/access_results).

## Acknowledgments

We want to acknowledge the participants and investigators of FinnGen study. The FinnGen project is funded by two grants from Business Finland (HUS 4685/31/2016 and UH 4386/31/2016) and the following industry partners: AbbVie Inc., AstraZeneca UK Ltd, Biogen MA Inc., Bristol Myers Squibb (and Celgene Corporation & Celgene International II Sàrl), Genentech Inc., Merck Sharp & Dohme Corp., Whitehouse Station, NJ, USA, Pfizer Inc., GlaxoSmithKline Intellectual Property Development Ltd., Sanofi US Services Inc., Maze Therapeutics Inc., Janssen Biotech Inc, Novartis AG, and Boehringer Ingelheim. Following biobanks are acknowledged for delivering biobank samples to FinnGen: Auria Biobank (www.auria.fi/biopankki), THL Biobank (www.thl.fi/biobank), Helsinki Biobank (www.helsinginbiopankki.fi), Biobank Borealis of Northern Finland (https://www.ppshp.fi/Tutkimus-ja-opetus/Biopankki/Pages/Biobank-Borealis-briefly-in-English.aspx), Finnish Clinical Biobank Tampere (www.tays.fi/en-US/Research_and_development/Finnish_Clinical_Biobank_Tampere), Biobank of Eastern Finland (www.ita-suomenbiopankki.fi/en), Central Finland Biobank (www.ksshp.fi/fi-FI/Potilaalle/Biopankki), Finnish Red Cross Blood Service Biobank (www.veripalvelu.fi/verenluovutus/biopankkitoiminta) and Terveystalo Biobank (www.terveystalo.com/fi/Yritystietoa/Terveystalo-Biopankki/Biopankki/). All Finnish Biobanks are members of BBMRI.fi infrastructure (www.bbmri.fi). Finnish Biobank Cooperative -FINBB (https://finbb.fi/) is the coordinator of BBMRI-ERIC operations in Finland. The Finnish biobank data can be accessed through the Fingenious® services (https://site.fingenious.fi/en/) managed by FINBB.

We thank Peter Vandehaar for technical consultation on PheWeb

## Contributions

Mitja I. Kurki: FinnGen Analysis Team Leader, data analysis, Browser development, wrote the first draft of the manuscript; Juha Karjalainen: Analysis, Browser development, paper editing; Priit Palta: Paper writing, FinnGen genotype data development; Timo P. Sipilä: team leader; Kati Kristiansson: Paper editing, FinnGen phenotype/register data development; Kati Donner: Paper editing, Data quality control; Mary P. Reeve: Paper editing, Visualizations; Hannele Laivuori: Paper editing, Study design, FinnGen phenotype/register data development; Mervi Aavikko: Paper editing, Study design, Team coordination; Mari A. Kaunisto: Paper editing, Visualizations, Variant novelty annotation; Anu Loukola: Paper editing, Methods / Biobank samples & Supplementary Methods Section 2; Elisa Lahtela: Paper editing, FinnGen phenotype/register data development, clinical team coordination; Hannele Mattsson: FinnGen phenotype/register data development; Päivi Laiho: Overseeing and organizing sample preparation and logistics; Pietro Della Briotta Parolo: Computation tool development, population structure analysis; Arto Lehisto: Computation tool development, fine mapping; Masahiro Kanai: Provided fine-mapped GTEx v8 data, UKBB quant pheno finemapping, contributed original finemapping pipeline; Nina Mars: Paper editing; Joel Rämö: Paper editing, Ear, Nose, and Throat endpoint interpretation; Tuomo Kiiskinen: Paper editing, FinnGen phenotype/register data development, data analysis; Henrike O. Heyne: Paper editing, annotating/interpreting ClinVar variants; Kumar Veerapen: Paper editing and finalization, demo phenotype summ stats curation; Sina Rüeger: Tool development, colocalization analysis; Susanna Lemmelä: Paper editing, Creating all phenotype plots and tables to the paper.; Wei Zhou: GWAS analysis contribution; Sanni Ruotsalainen: Association analysis; Kalle Pärn: Genotype data development; Tero Hiekkalinna: Endpoint development, Register data processing; Sami Koskelainen: Register data processing; Teemu Paajanen: Data processing; Vincent Llorens: Endpoint development, Tool development, construction of RISTEYS; Javier Gracia-Tabuenca: Paper editing, Longitudinal data analysis; Harri Siirtola: Paper editing, Visualization of registry data in FinnGen endpoints; Kadri Reis: Replication analysis, Paper editing; Abdelrahman G. Elnahas: Replication analysis; Katriina Aalto-Setälä: Paper editing; Kaur Alasoo: Paper editing, Peformed fine mapping on the eQTL Catalogue; Mikko Arvas: Phenotype and HLA analysis, Paper editing; Kirsi Auro: Study design, overseeing the project, Paper editing; Shameek Biswas: Paper editing; Argyro Bizaki-Vallaskangas: Paper editing, Endpoint development; Olli Carpen: Paper editing, Study design, endpoint development; Chia-Yen Chen: Data analysis, Paper editing; Oluwaseun A. Dada: Data processing, cloud administration; Zhihao Ding: Study design, data analysis, Paper editing; Margaret G. Ehm: Study design, overseeing the project, Paper editing; Kari Eklund: Endpoint development, Paper editing; Martti Färkkilä: Endpoint development, study design, Paper editing; Hilary Finucane: Provided fine-mapped GTEx v8 data and UKBB quant phenos, Paper editing; Andrea Ganna: Study design, overseeing tool development, Paper editing; Awaisa Ghazal: Participated in R4 imputation, full responsibility of running R5 production runs, Paper editing; Robert R. Graham: Study design, overseeing the project, Paper editing; Eric Green: Study design, overseeing the project, Paper editing; Antti Hakanen: Study design, overseeing the project, Paper editing; Marco Hautalahti: Biobank coordination; Åsa Hedman: Data analysis, Paper editing; Mikko Hiltunen: Cohort collection, clinical expertise, Paper editing; Reetta Hinttala: Data analysis, Paper editing; Iiris Hovatta: Endpoint development, Paper editing; Xinli Hu: Data analysis, Paper editing; Adriana Huertas-Vazquez: Data analysis, Paper editing; Laura Huilaja: Endpoint development, Paper editing; Julie Hunkapiller: Study design, overseeing the project, Paper editing; Howard Jacob: Study design, overseeing the project, Paper editing; Jan-Nygaard Jensen: Study design, Paper editing; Heikki Joensuu: Endpoint development, Paper editing; Sally John: Study design, overseeing the project, Paper editing; Valtteri Julkunen: Enpoint development, study design, Paper editing; Marc Jung: Data analysis, Paper editing; Juhani Junttila: Study design, overseeing the project, Paper editing; Kai Kaarniranta: Endpoint develpoment, Paper editing; Mika Kähönen: Endpoint development, overseeing project, Paper editing; Risto M. Kajanne: Project coordination, Paper editing; Lila Kallio: Sample collection, coordination; Reetta Kälviäinen: Endpoint develpoment, Paper editing; Jaakko Kaprio: Study design, register expertise, Paper editing; Nurlan Kerimov: Peformed fine-mapping on the eQTL Catalogue, Paper editing; Johannes Kettunen: Data analysis, Paper editing; Elina Kilpeläinen: Imputation and QC of R1-5 datasets, Paper editing; Terhi Kilpi: Study design, overseeing project, Paper editing; Katherine Klinger: Study design, overseeing project, Paper editing; Veli-Matti Kosma: Study design, overseeing project, Paper editing; Teijo Kuopio: Sample collection, coordination; Venla Kurra: Endpoint development, Paper editing; Triin Laisk: Data analysis, Paper editing; Jari Laukkanen: Study design, overseeing project, Paper editing; Nathan Lawless: Study design, overseeing project, Paper editing; Aoxing Liu: Data analysis, Paper editing; Simonne Longerich: Study design, overseeing project, Paper editing; Reedik Mägi: Genotype data preparation, Paper editing; Johanna Mäkelä: Sample collection, coordination; Antti Mäkitie: Endpoint development, Paper editing; Anders Malarstig: Study design, overseeing project, Paper editing; Arto Mannermaa: Study design, overseeing project, Paper editing; Joseph Maranville: Study design, overseeing project, Paper editing; Athena Matakidou: Study design, overseeing project, Paper editing; Tuomo Meretoja: Endpoint development, Paper editing; Sahar V. Mozaffari: Data analysis, Paper editing; Mari EK. Niemi: Study design, Study coordination, Paper editing; Marianna Niemi: Tool and endpoint development, Paper editing; Teemu Niiranen: Endpoint development, Paper editing; Christopher J. O’Donnell: Study design, overseeing project, Paper editing; Ma’en Obeidat: Data analysis, Paper editing; George Okafo: Study design, overseeing project, Paper editing; Hanna M. Ollila: Data analysis, Paper editing; Antti Palomäki: Endpoint development and validation, data-analysis, study design, Paper editing; Tuula Palotie: Clinical definitions and endpoint development, Paper editing; Jukka Partanen: Study design, overseeing project, HLA imputation, Paper editing; Dirk S. Paul: Study design, overseeing project, Paper editing; Margit Pelkonen: Enpoint development, Paper editing; Rion K. Pendergrass: Data analysis, Paper editing; Slavé Petrovski: Study design, Paper editing; Anne Pitkäranta: Study design, overseeing project, Paper editing; Adam Platt: Study design, overseeing project, Paper editing; David Pulford: Data analysis, Paper editing; Eero Punkka: Sample collection, coordination; Pirkko Pussinen: Endpoint definition, Paper editing; Neha Raghavan: Data analysis, Paper editing; Fedik Rahimov: data analysis, Paper editing; Deepak Rajpal: data analysis, Paper editing; Nicole A. Renaud: data analysis, Paper editing; Bridget Riley-Gillis: Study design, overseeing project, Paper editing; Rodosthenis Rodosthenous: Study design, study coordination, Paper editing; Elmo Saarentaus: Paper editing, Ear, Nose, and Throat endpoint interpretation; Aino Salminen: Endpoint development, Paper editing; Eveliina Salminen: Endpoint development, study design, Paper editing; Veikko Salomaa: Endpoint development, study design, Paper editing; Johanna Schleutker: Endpoint development, Paper editing; Raisa Serpi: Sample collection, coordination; Huei-yi Shen: Project coordination, Paper editing; Richard Siegel: Study design, Paper editing; Kaisa Silander: Biobank expertise, Paper editing; Sanna Siltanen: Study design, overseeing project, Paper editing; Sirpa Soini: Biobank and legal expertise, Paper editing; Hilkka Soininen: Endpoint development, clinical expertise, Paper editing; Jae H. Sul: data analysis, Paper editing; Ioanna Tachmazidou: data analysis, Paper editing; Kaisa Tasanen: Endpoint development, clinical expertise, Paper editing; Pentti Tienari: Neurology end-point definitions, review the paper, Paper editing; Sanna Toppila-Salmi: Paper editing; Taru Tukiainen: Study Design, Data analysis, Paper editing; Tiinamaija Tuomi: Endpoint development, study design, Paper editing; Joni A. Turunen: Ophthalmology end-point definitions, review the paper, Paper editing; Jacob C. Ulirsch: Fine mapping, Paper editing; Felix Vaura: Endpoint development, Paper editing; Petri Virolainen: Study design, overseeing the project, Paper editing; Jeffrey Waring: Study design, data analysis, Paper editing; Dawn Waterworth: Study design, Paper editing; Robert Yang: Study design, Paper editing; Mari Nelis: Study design, Paper editing; Anu Reigo: Study design, Paper editing; Andres Metspalu: Study design, Head of the Estonian Biobank, paper editing; Lili Milani: Study design, Paper editing; Tõnu Esko: Study design, Paper editing; Caroline Fox: Study design, overseeing the project, Paper editing; Aki S. Havulinna: Paper editing, FinnGen endpoint concept and implementation, register data development; Markus Perola: Paper editing, Study design, Ethical expert, THL science co-ordinator; Samuli Ripatti: Paper editing, Study design, Data-analysis expertise; Anu Jalanko: Paper editing, participation to strategic planning of the project; Tarja Laitinen: Paper editing, Study design, overseeing the project; Tomi Mäkelä: Paper editing, Study design, overseeing the project; Robert Plenge: Paper editing, Study design, overseeing the project; Mark McCarthy: Paper writing and editing, Study design, overseeing the project; Heiko Runz: Paper writing and editing, Study design, overseeing the project; Mark J. Daly*: Paper writing and editing, Study design, overseeing the project; Aarno Palotie*: Paper writing and editing, Study design, overseeing the project

## Supplementary Materials

**Supplementary File 1. Comparison of effects size in known genome wide significant loci between FinnGen and large published reference GWAS (Table 1**). Y and X axes represent FinnGen and reference GWAS effect sizes respectively. Betas are aligned to be positive in reference studies. Lines extending from points indicate standard errors in respective studies. Regression lines omit intercept and two type of regressions are provided: unweighted and weighted by pooled standard errors from the two studies. Solid line indicates identity line and dotted line and dashed lines indicate unweighted and weighted regression respectively. Only variants with p < 10^-10^ in reference study were included to mitigate the effect of winner’s curse of inflated betas in the reference studies.

**Supplementary Data 1. List of variants, summary stats and references for Supplementary File 1 figures.**

**Supplementary Tables**. This file contains Supplementary Tables 1-10 and table legends.

**Supplementary Methods**. This Supplementary Methods file contains the following section: Phenotyping from nationwide population-based health registers; FinnGen participant recruitment and legacy cohorts, Genotyping and genotype data QC; Population structure and cryptic relatedness; GWAS/pheWAS Analysis; Data Access and Dissemination. All Supplementary Figures are inlined in the appropriate sections.

**FinnGen banner author list.** List of all FinnGen working group members and their affiliations

### BOX 1 - Noteworthy novel findings from 15 previously well-studied benchmark diseases

#### Age-related macular degeneration

A Finnish enriched stop loss variant in *CFI* was associated with age-related macular degeneration (FinnGen OR 1.92. P: 1.8*10^-7^). The variant has been observed almost exclusively in Finnish population (gnomAD AF 1.3%) but exists at a very low frequency in Estonian (gnomAD AF 0.02%) and ‘other’ populations in gnomAD (AF 0.008%). Remarkably, the association was Consistent with the variant consequence, we observed an eQTL colocalization with strong gene expression lowering effect of *CFI* gene transcript in adipose tissue (CLPP 0.46, beta -1.6) in a FUSION transcriptome^59^ study using Finnish samples. *CFI* codes for complement factor I, a regulator of classical and alternative complement pathways. Rare recessive missense and stop gained variants in *CFI* have been observed in complement factor I deficiency and, consistent with the eQTL directionality here, heterozygous carriers of such reduced function variants have been strongly associated with AMD in exome sequencing of AMD GWAS loci^60^.

#### Atrial fibrillation

Four novel low frequency (MAF 0.1%-2%) associations were observed for atrial fibrillation and flutter. In all four we identified a putative causal coding variant in credible sets. Two independent coding variants were observed in *RRPL3.* Both variant associations were supported by Estonian and UKBB replication. Coding variants in *RPL3L* that are not in LD (r2<0.005 to lead variants in FinnGen imputation panel) with our associations have been previously associated with atrial fibrillation. In a GWAS meta-analysis with 14,710 cases and 373,897 controls from Iceland and 14,792 cases and 393,863 controls from the UK Biobank, a missense and splice donor variants in *RPL3L* were associated with atrial fibrillation^61^

The third atrial fibrillation association lead variant (chr19_50497261_C_T, AF 2%, OR 1.43, p 1.92 * 10^-12^) was 16.6 times enriched in Finland. The variant exists in the Estonian population and the effect size was consistent although markedly lower and not statistically significant ( AF 0.6%, OR 1.16, p 0.46). The variant did not exist in UKBB data (North-Western europeans in GnomAD AF 0.1%). We observed a missense variant (p.Arg1845Trp) in *MYH14* in the credible set, as well as eQTL colocation with upregulation of putative long non-coding RNA (ENSG00000268518) in skeletal muscle in the Finnish FUSION study^59^. Variants in *MYH14* can cause autosomal dominant peripheral neuropathy and deafness (https://www.omim.org/entry/608568) but no cardiac phenotype has been previously reported.

In the fourth association (AF 1.3%, OR 1.5, p 8.17*10^-11^) we observed a splice donor variant (c.105+1G>T) in the *SYNPO2L* gene (PIP 0.13). The variant was extremely rare in Estonia (0.05%) but showed consistent effect direction (OR 1.23) although not significant due to lack of power in such rare variant association. The variant was absent in UKBB and the AF in NFSEE in gnomad is extremely low (0.02%). An intronic common variant in *SYNPO2L* has been previously associated with atrial fibrillation^62^ and our results provide direct coding variant evidence of *SYNPO2L* being the causal gene in this locus. Further insights on atrial fibrillation from FinnGen coding variants have been described by Sun et al^21^.

#### Prostate cancer

Two novel associations were identified for prostate cancer. The first was a 16-fold Finnish-enriched variant ( chr22_43082544_C_T, AF 0.3%, OR 3.49, p 5.44*10^-8^) and showed consistent effect size in Estonia (AF 0.09%, OR 3, p 0.2) but was absent in UKBB (meta-analysis p 2.45*10^-8^). An inframe deletion variant (p.Ala139_Leu148del) in *BIK* gene was observed in the credible set (PIP 0.44). Common variants in the locus have been previously associated with prostate cancer ^63^. *BIK* (BCL-2 interacting killer) gene product is a pro-apoptotic protein that has been suggested to act as a tumor suppressor gene^64^ and to be a marker for a more aggressive breast cancer^65^, however no direct genetic evidence has previously been published on variants in *BIK* causing prostate cancer.

The other prostate cancer association (chr17_42937117_C_G, OR 7.1 p 1.21 * 10^-8^, MAF 0.1%) was a very rare variant with only 3 copies observed in gnomAD (2 in Finns and 1 in ‘Other’ gnomAd population). Replication of the association was not possible due to the variant’s rarity in other populations and replication in an independent cohort is warranted. The lead variant (PIP 0.62) is a non-coding variant downstream from *SNORA40* and the other equally rare (3 copies in Finns and 1 in ‘Other’ populations in gnomAD) variant (17:42372576:G:C, rs1202093388, MAF 0.09%, OR 9.4, 5.1*10^-8^, PIP 0.37) in 95% credible set resides 600kb away in an intron of *STAT3. SNORA40* codes for small nucleolar RNA (SNORA), the role of which in cancer are not very well characterized but differential expression associations of several SNORAs with different cancers have been observed^66^. Hyperactivation of *STAT3* however has been observed in the majority of cancers and their prognosis including prostate cancer^67^. *STAT3* is a transcription factor activated by IL6/JAK/STAT3-pathway and mediates many inflammatory processes. Chronic inflammation can induce development and progression of tumors^67^. Protein-truncating and missense variants in *STAT3* are depleted in human populations ( PTV observed/expected upper bound 0.1, missense observed/expected upped bound 0.36; gnomAD v2.1.1^13^). No effect on survival was observed in cases only survival analysis from diagnosis to death (hazard ratio 1.04, p 0.94).

#### Asthma

Another interesting locus where we observed coding variants (missense in *IL4R*, p.Ala82Thr, PIP 0.21) within credible sets was associated with both asthma (FinnGen AF 8.2%, OR 0.86, p 2.5*10^-12^, meta p 5.31*10^-9^) and psoriasis (FinnGen OR 1.28, p 3.48*10^-9^, meta p 1.92*10-11) but with an opposing direction of effect. *IL4R* codes for IL4α subunit that is part of receptor complexes for both cytokines *IL4* and *IL13*, which are key cytokines in the type II inflammatory response triggered by allergens or parasites (see^68^ for a detailed review). The key role of type II inflammatory response in asthma has long been recognized^69^ and variants in 5q31 locus containing the genes coding for *IL4* and *IL13* have been associated with asthma in GWA studies^70^. Asthma, atopic dermatitis and hay fever often co-occur and are referred collectively to as atopic diseases^71^. The effect direction of this association was consistent with that of asthma in atopic dermatitis (OR 0.9, p 2.4 * 10^-3^). The reversed effect direction in psoriasis was surprising as there is no evidence that in psoriasis there would be a contribution of type II inflammation but type | and Th17 mediated inflammation^72^.

