## Supplementary File 1 for "FinnGen: Unique genetic insights from combining isolated population and national health register data"

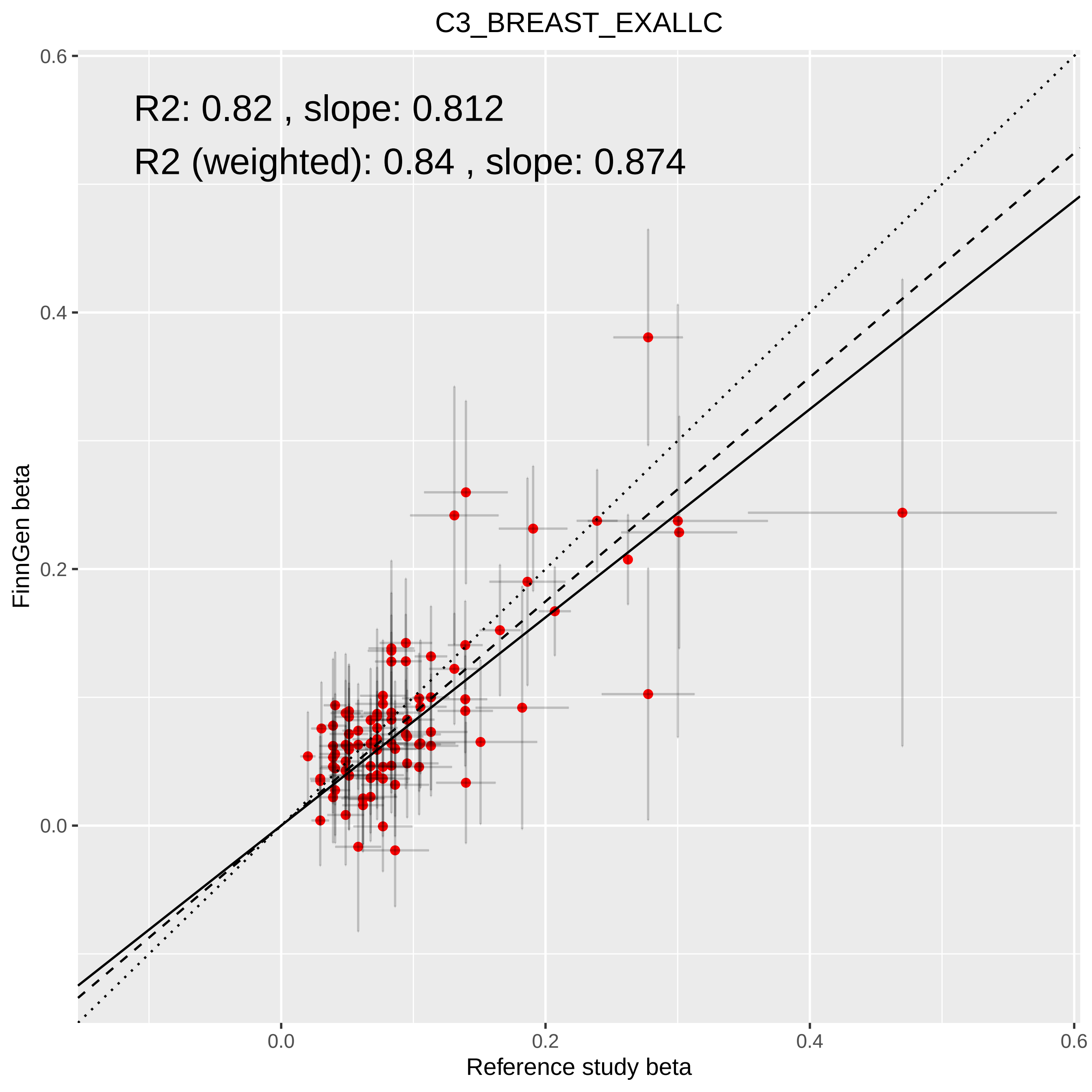

C3_BREAST_EXALLC
R2: 0.82 , slope: 0.812
R2 (weighted): 0.84 , slope: 0.874
FinnGen beta
Reference study beta
0.0
0.2
0.4
0.6

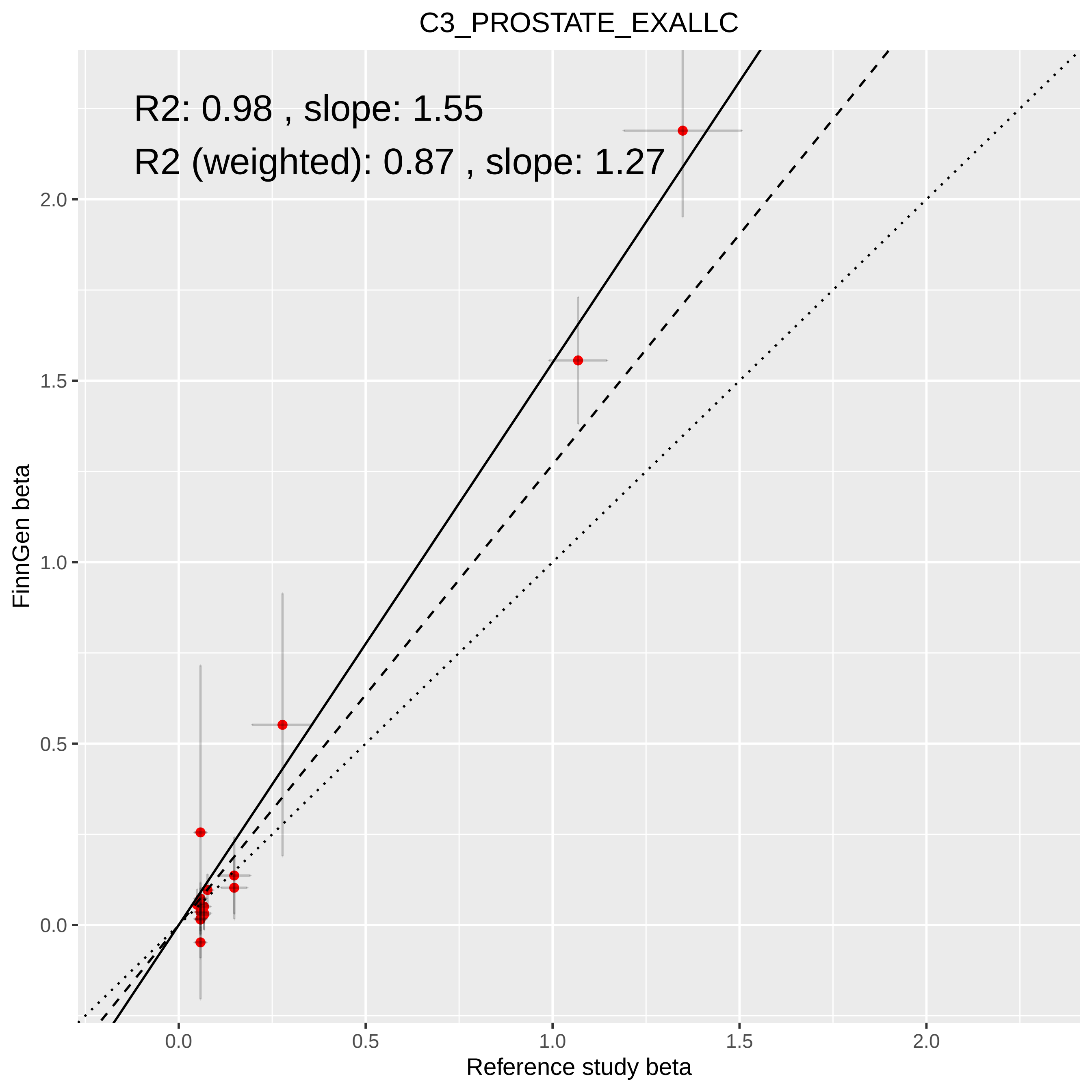
C3_PROSTATE_EXALLC
R2: 0.98 , slope: 1.55
R2 (weighted): 0.87 , slope: 1.27
FinnGen beta
Reference study beta
0.0
0.5
1.0
1.5
2.0

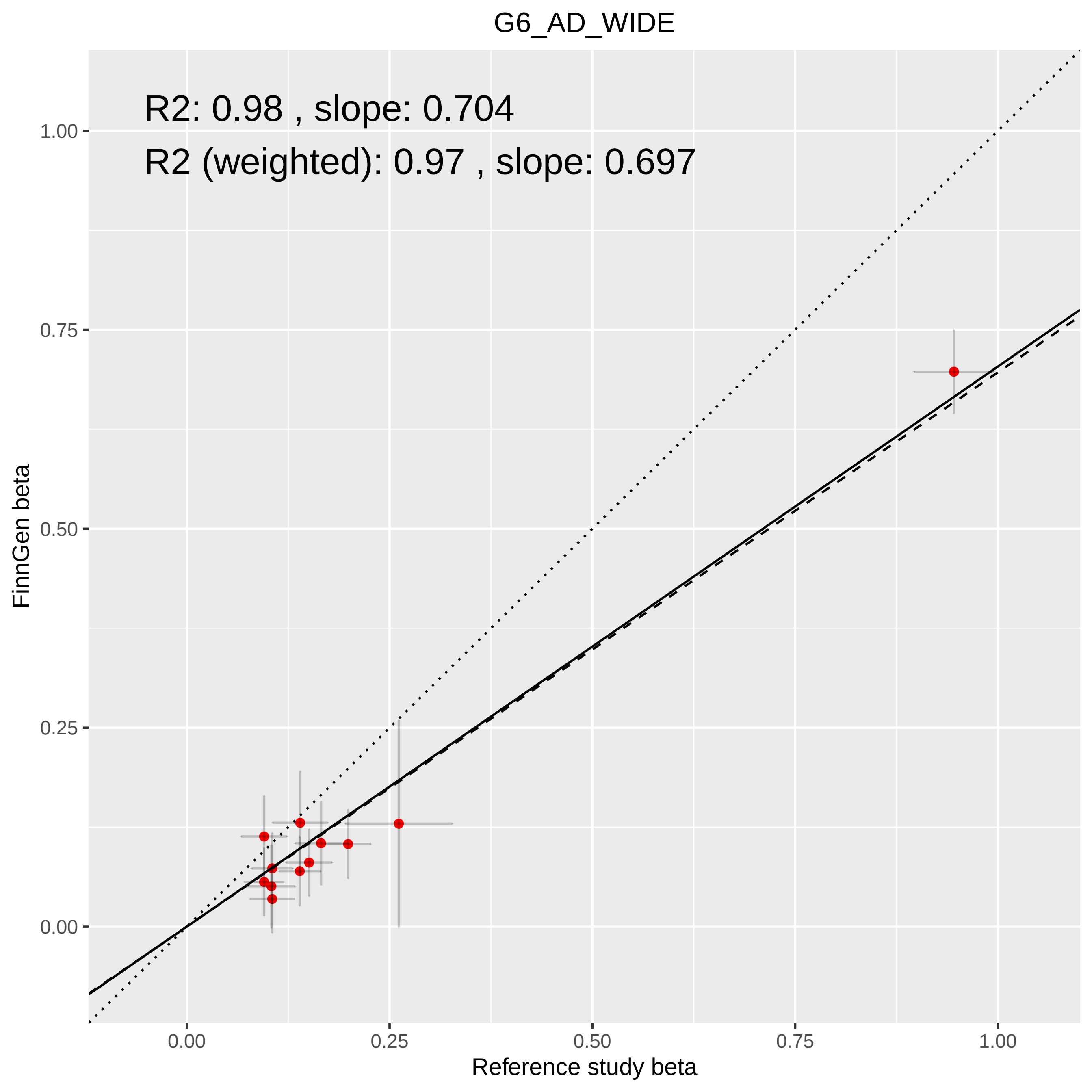
G6_AD_WIDE
R2: 0.98 , slope: 0.704
R2 (weighted): 0.97 , slope: 0.697
FinnGen beta
Reference study beta
0.00
0.25
0.50
0.75
1.00

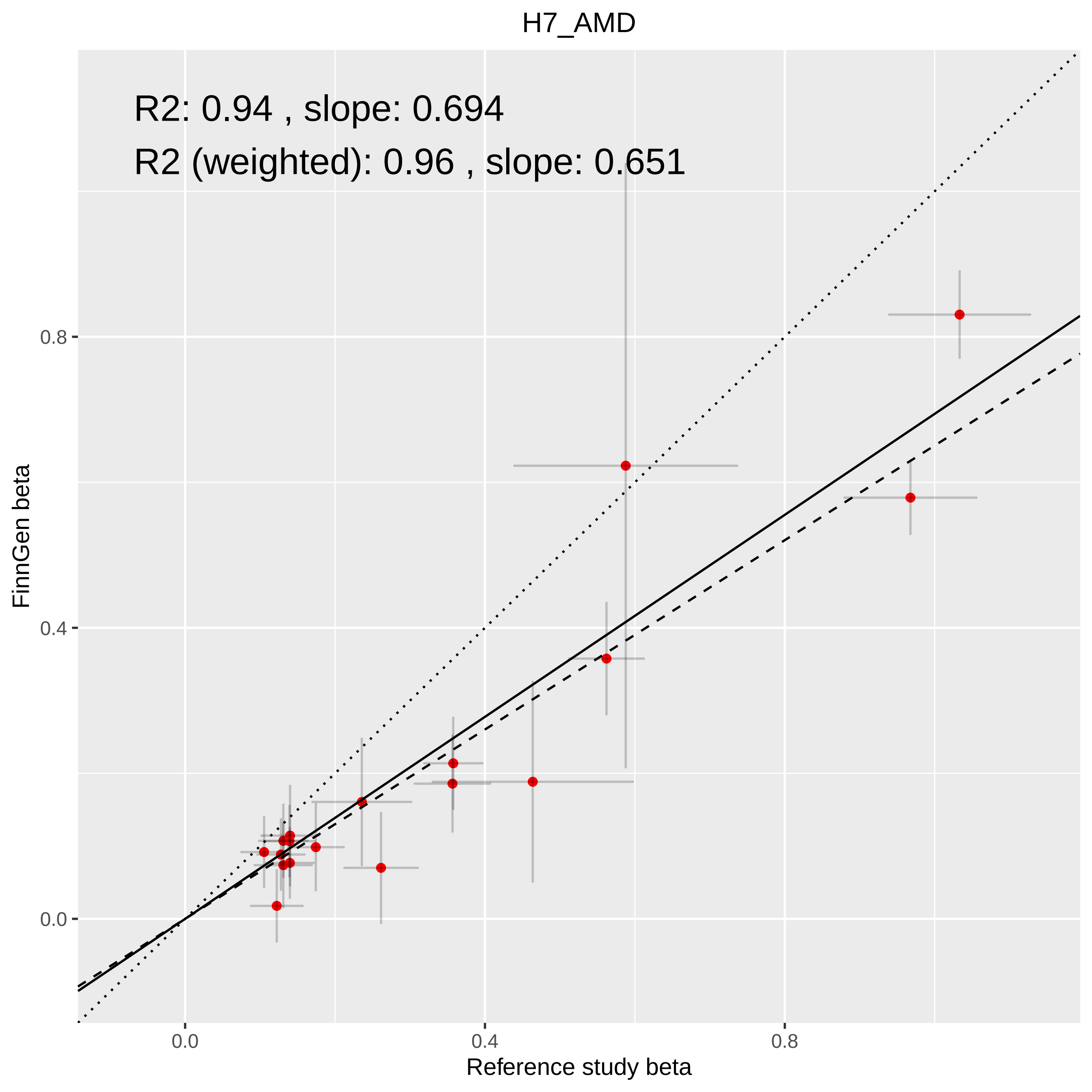
H7_AMD
R2: 0.94 , slope: 0.694
R2 (weighted): 0.96 , slope: 0.651
FinnGen beta
0.0
0.4
0.8
0.0
0.4
0.8
Reference study beta

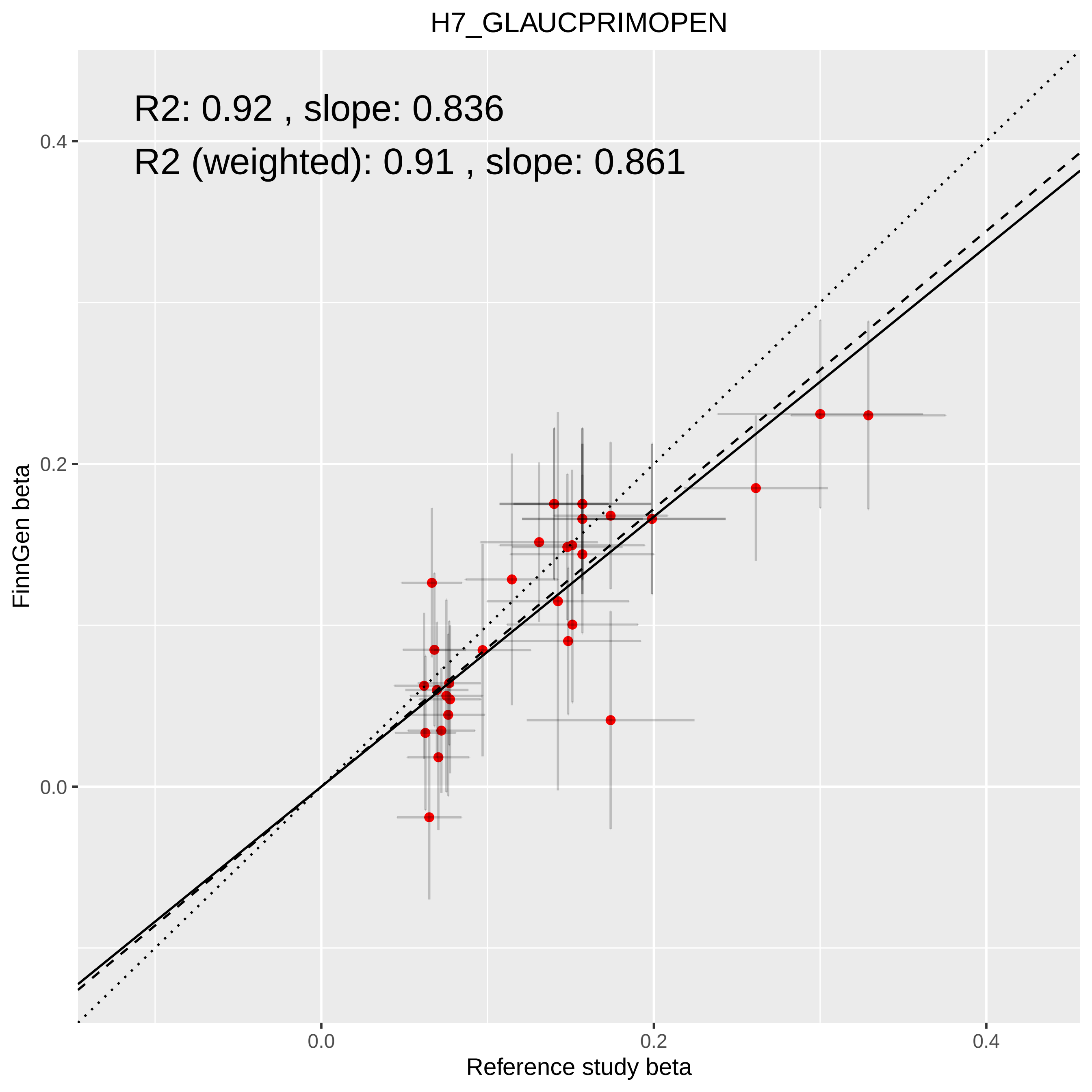
H7_GLAUCPRIMOPEN
R2: 0.92 , slope: 0.836
R2 (weighted): 0.91 , slope: 0.861
FinnGen beta
0.4
0.2
0.0
Reference study beta
0.0
0.2
0.4

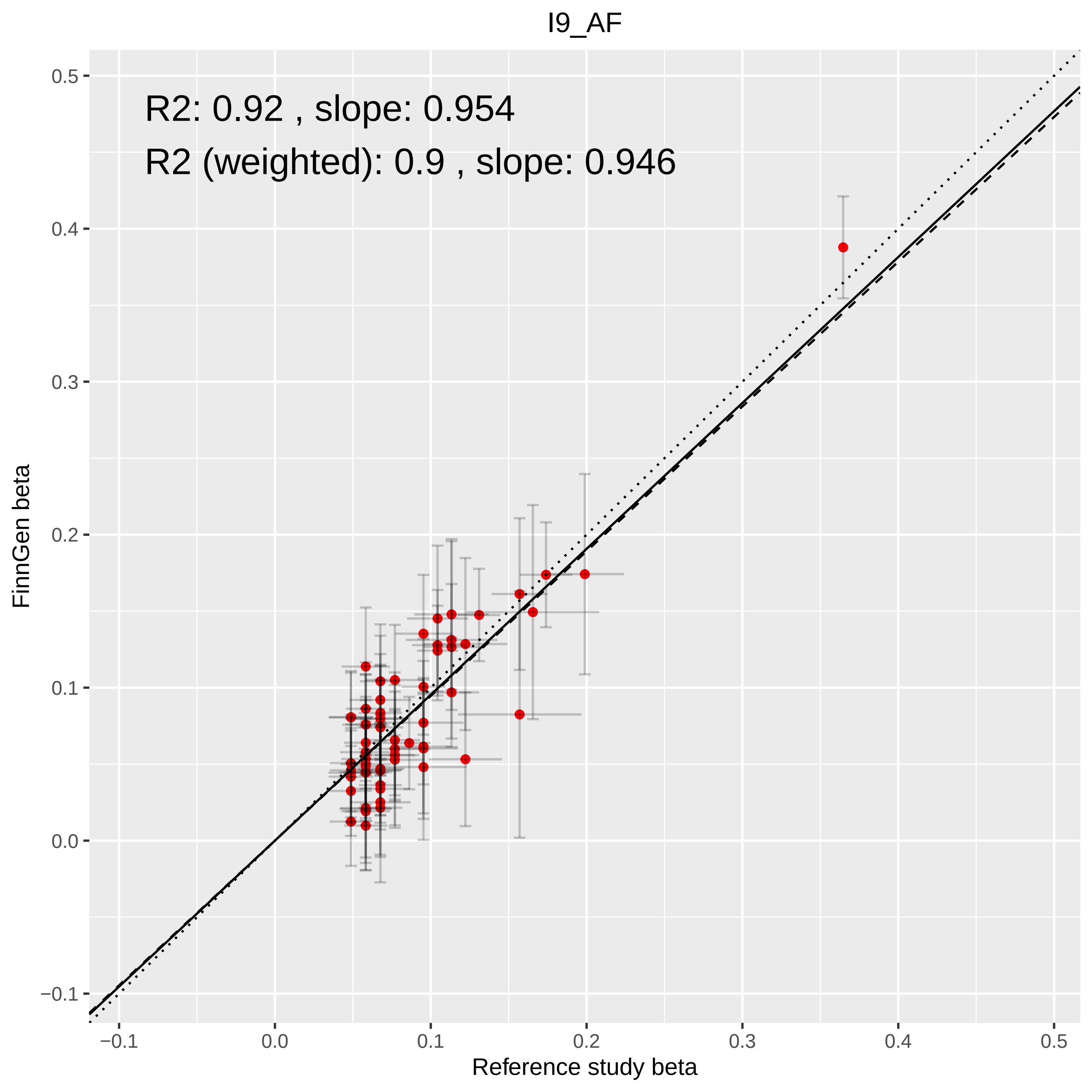

I9_AF
R2: 0.92 , slope: 0.954
R2 (weighted): 0.9 , slope: 0.946
FinnGen beta
Reference study beta
0.5
0.4
0.3
0.2
0.1
0.0
−0.1
−0.1
0.0
0.1
0.2
0.3
0.4
0.5

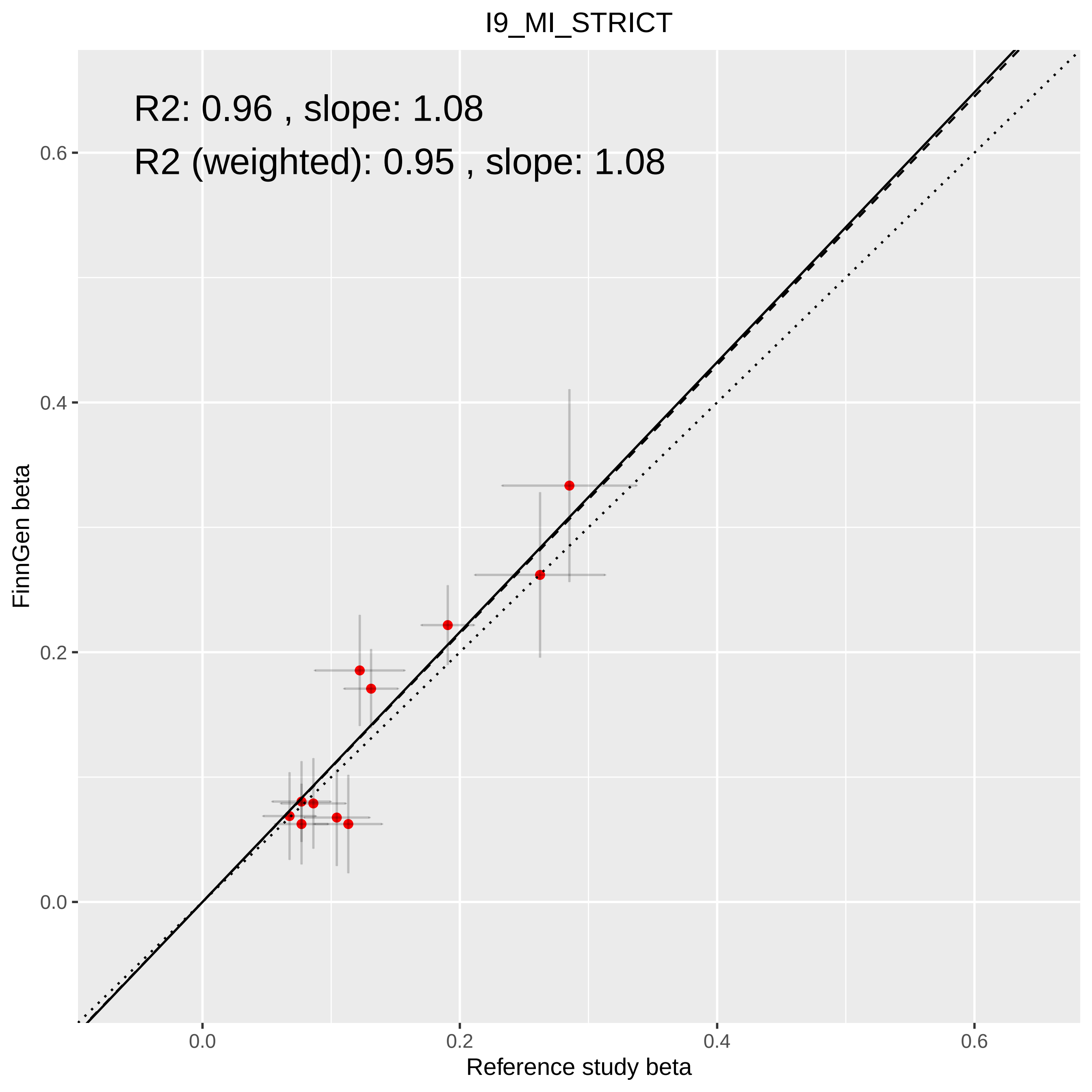
I9_MI_STRICT
R2: 0.96 , slope: 1.08
R2 (weighted): 0.95 , slope: 1.08
0.6
0.4
0.2
0.0
FinnGen beta
0.0
0.2
0.4
0.6
Reference study beta

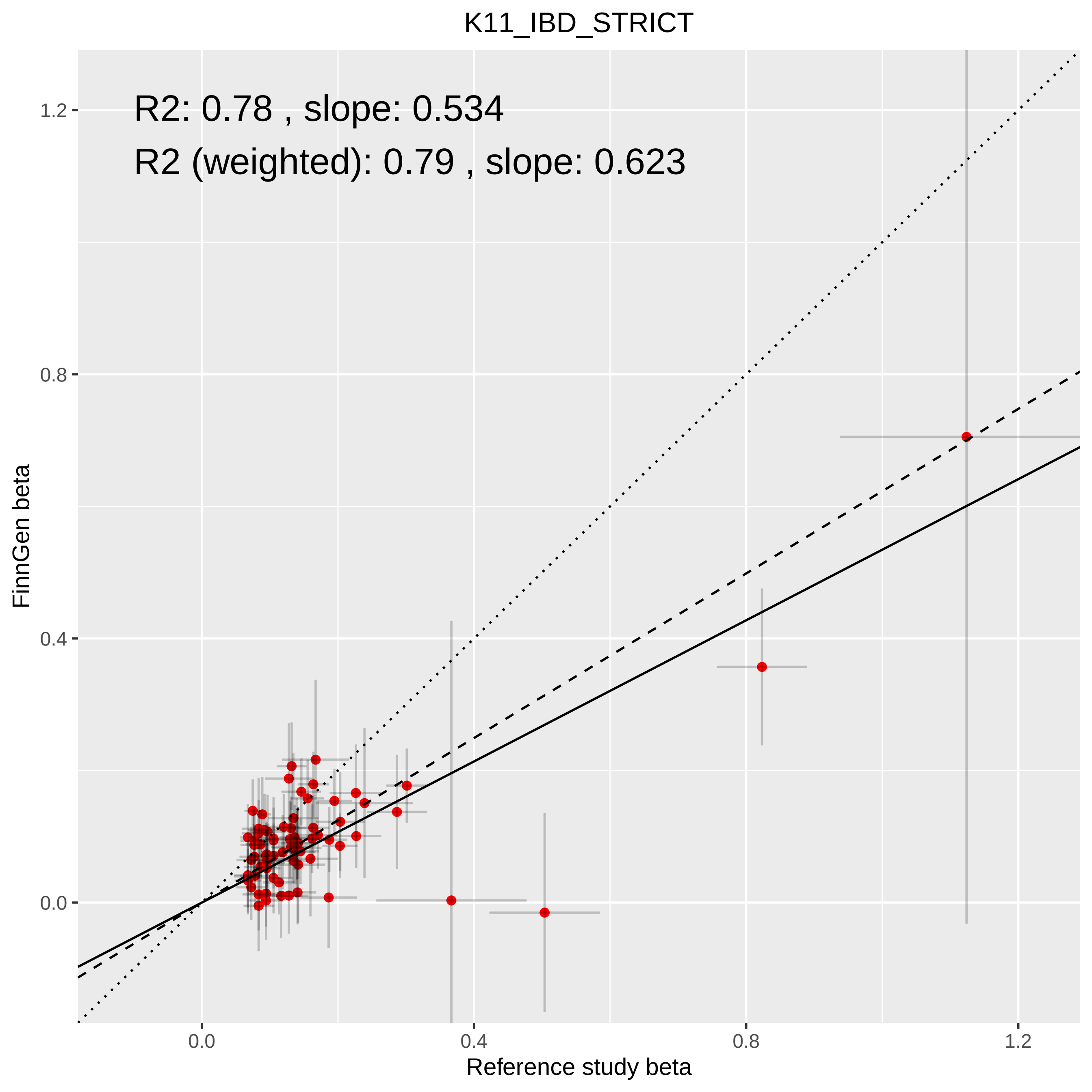
K11_IBD_STRICT
R2: 0.78 , slope: 0.534
R2 (weighted): 0.79 , slope: 0.623
FinnGen beta
Reference study beta
1.2
0.8
0.4
0.0
0.0
0.4
0.8
1.2

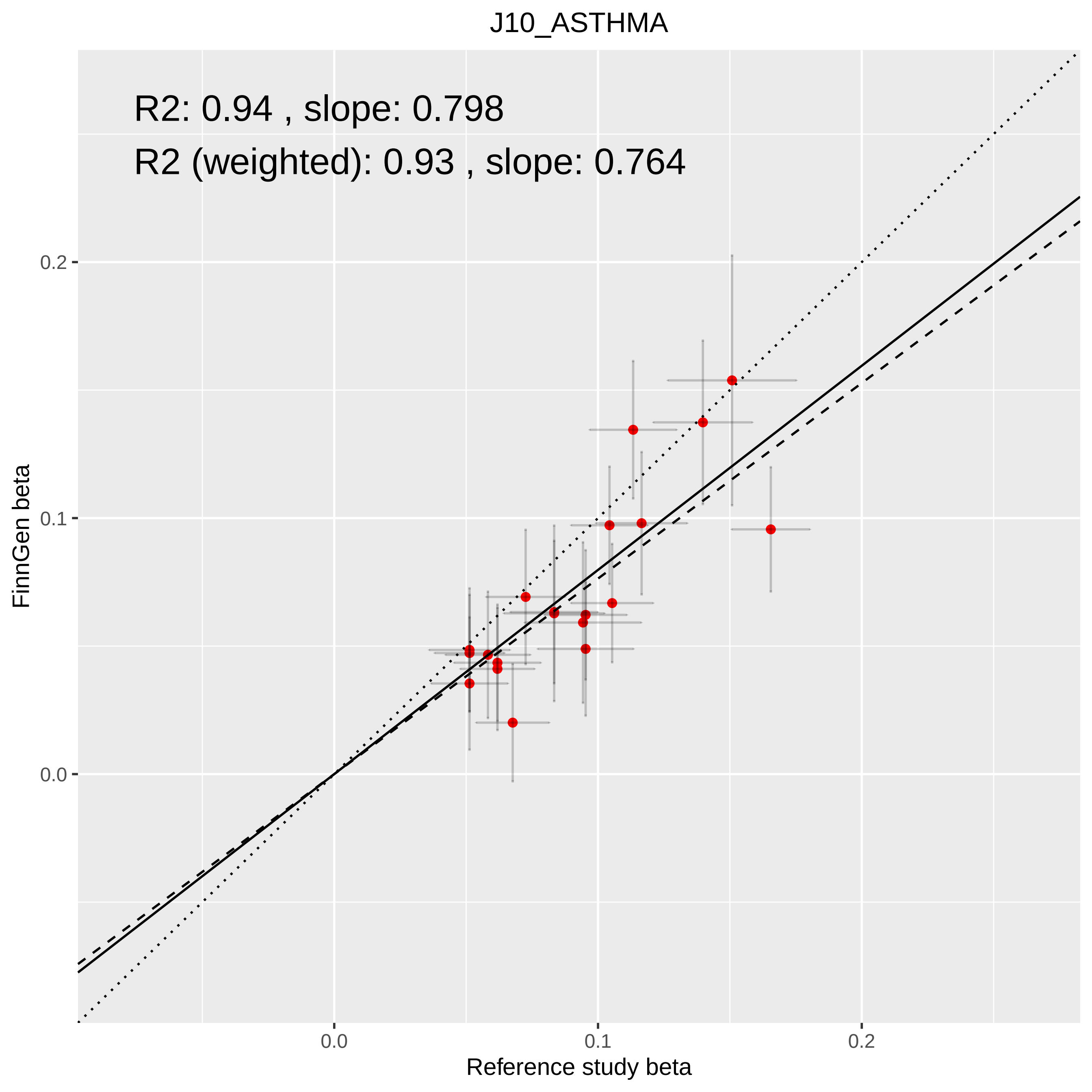

J10_ASTHMA
R2: 0.94 , slope: 0.798
R2 (weighted): 0.93 , slope: 0.764
FinnGen beta
0.2
0.1
0.0
0.0
0.1
0.2
Reference study beta

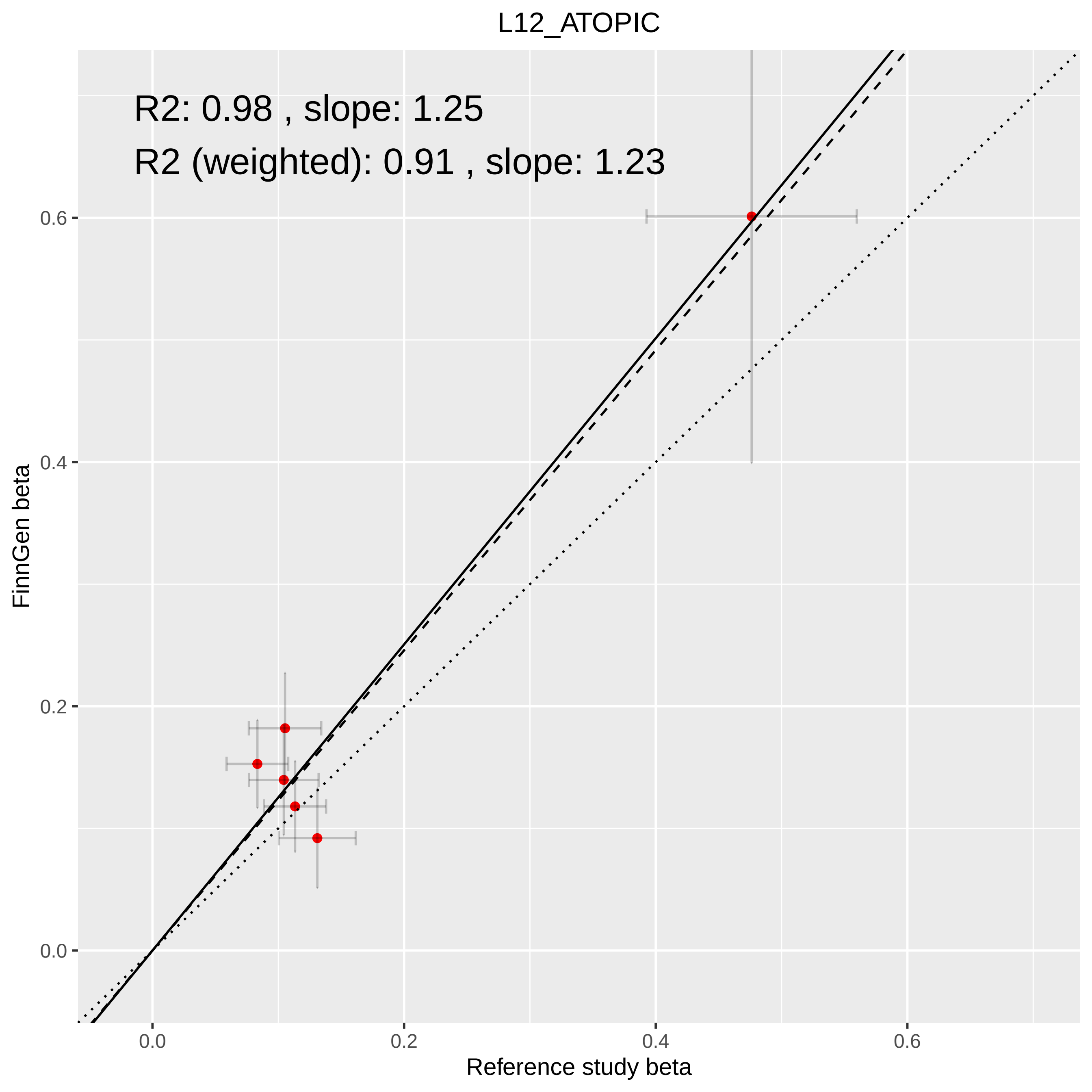

L12_ATOPIC
R2: 0.98 , slope: 1.25
R2 (weighted): 0.91 , slope: 1.23
0.6
0.4
0.2
0.0
FinnGen beta
0.0
0.2
0.4
0.6
Reference study beta

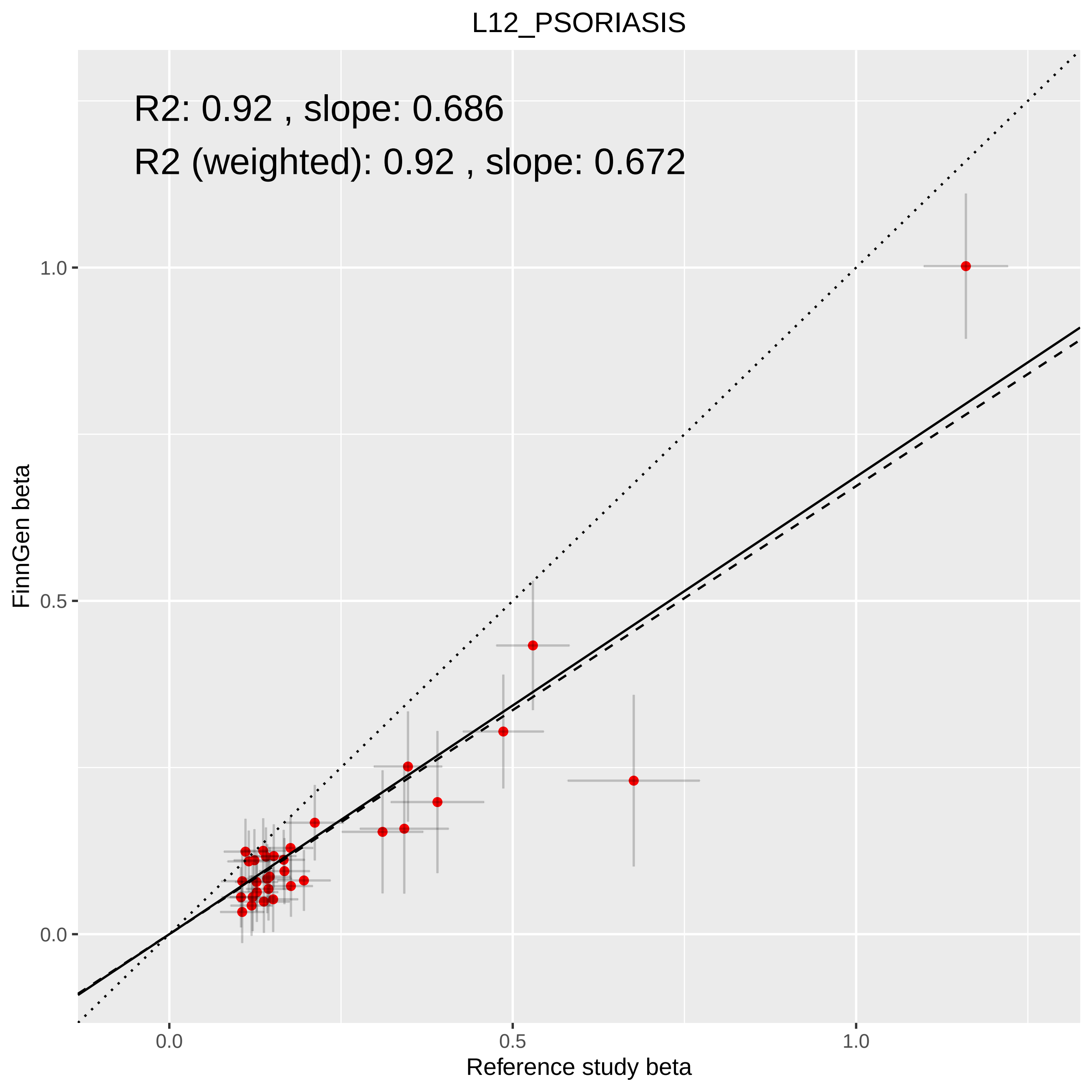

L12_PSORIASIS
R2: 0.92 , slope: 0.686
R2 (weighted): 0.92 , slope: 0.672
FinnGen beta
1.0
0.5
0.0
0.0
0.5
1.0
Reference study beta

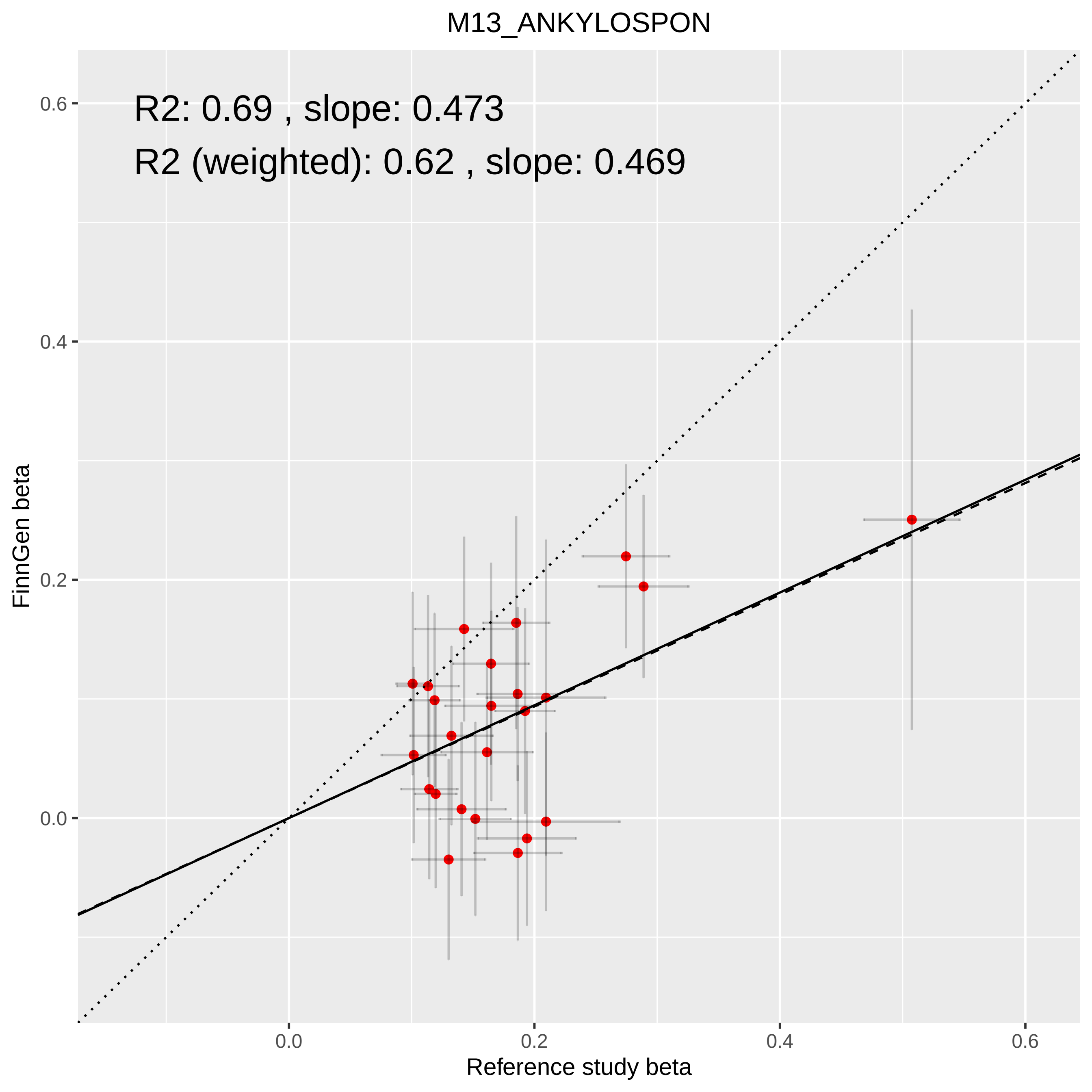

M13_ANKYLOSPON
R2: 0.69 , slope: 0.473
R2 (weighted): 0.62 , slope: 0.469
0.6
0.4
0.2
0.0
0.0
0.2
0.4
0.6
FinnGen beta
Reference study beta

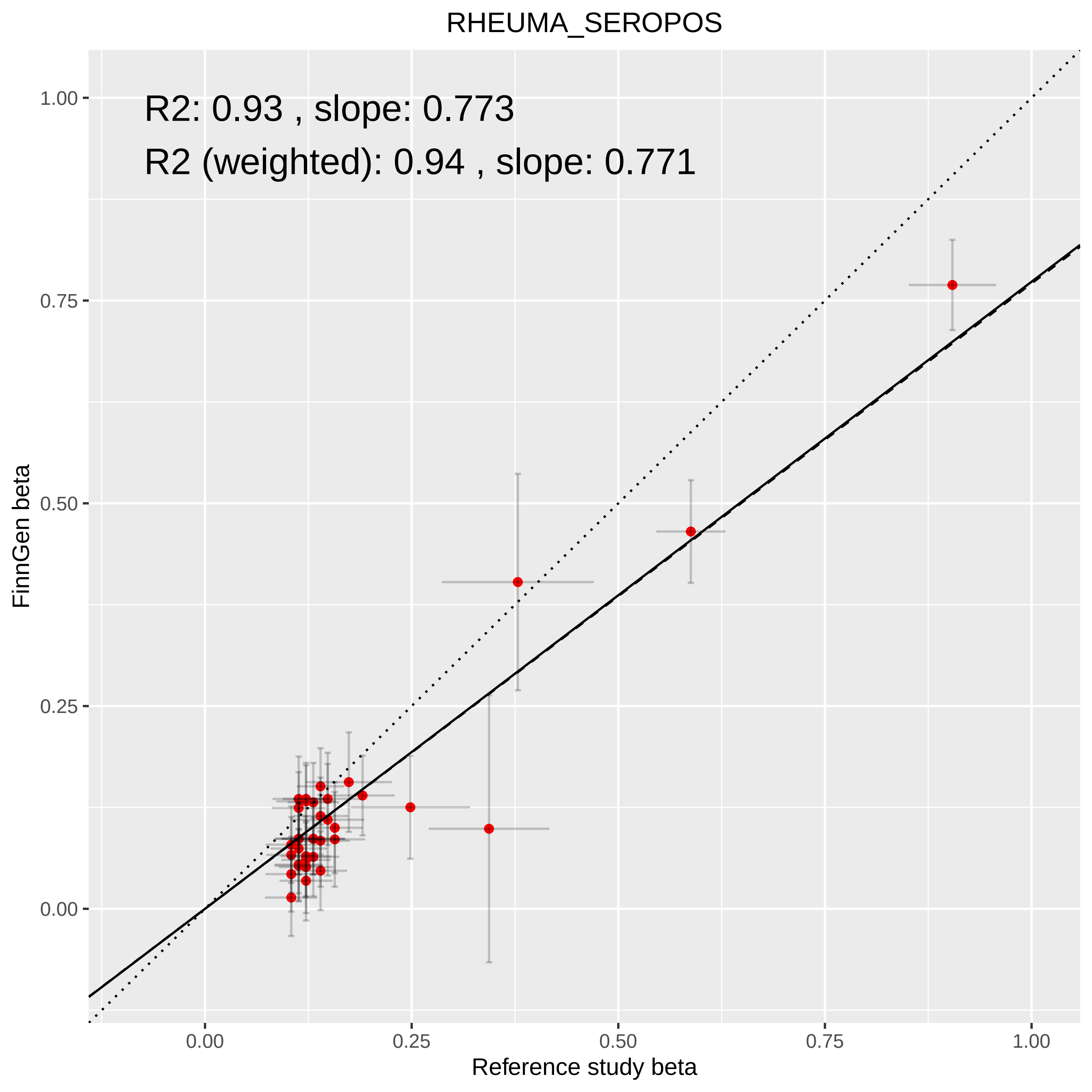
RHEUMA_SEROPOS
R2: 0.93 , slope: 0.773
R2 (weighted): 0.94 , slope: 0.771
FinnGen beta
Reference study beta
0.00
0.25
0.50
0.75
1.00

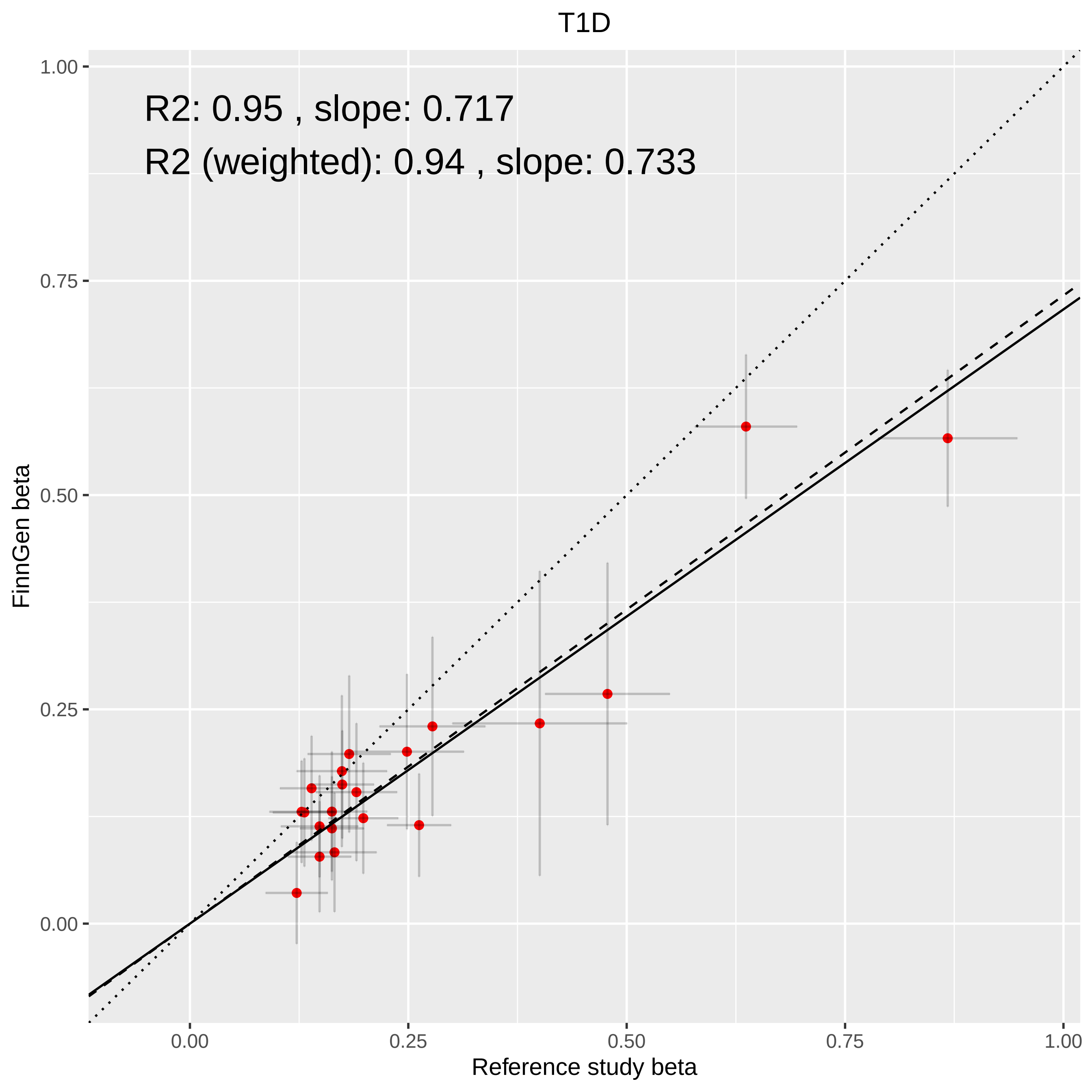

T1D
R2: 0.95 , slope: 0.717
R2 (weighted): 0.94 , slope: 0.733
FinnGen beta
Reference study beta
0.00
0.25
0.50
0.75
1.00

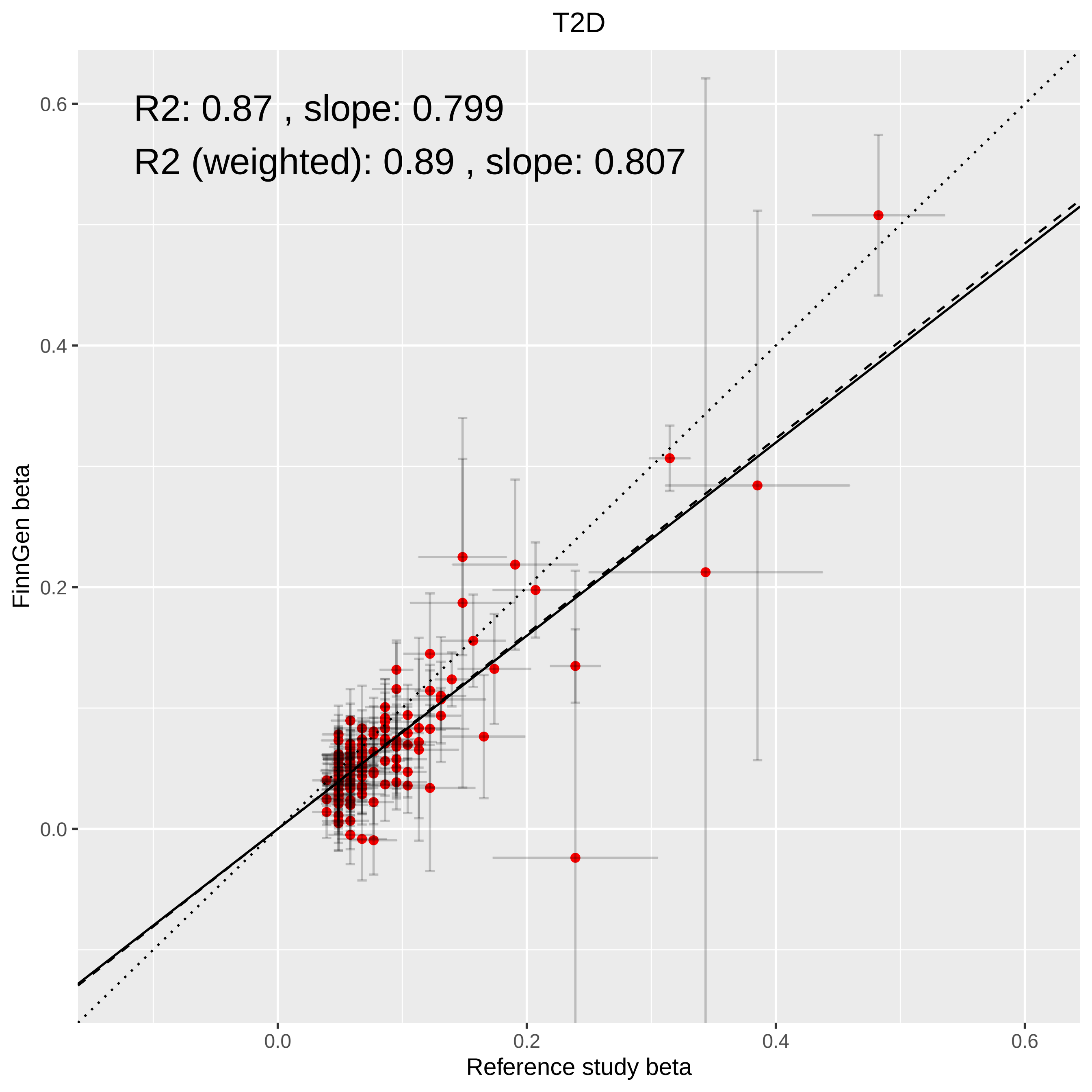
T2D
R2: 0.87 , slope: 0.799
R2 (weighted): 0.89 , slope: 0.807
FinnGen beta
Reference study beta
0.0
0.2
0.4
0.6
0.0
0.2
0.4
0.6
