## Supplementary Methods for "FinnGen: Unique genetic insights from combining isolated population and national health register data"

Supplementary Methods and Figures

### **Section 1**: Phenotyping from nationwide population-based health registries

In Finland, similar to the other Nordic Countries, there are nationwide electronic health registers originally established primarily for administrative purposes to monitor the usage of healthcare nationwide and over the lifespan. These registers have almost complete coverage of major health-related events, such as hospitalizations, prescription drug purchases (not including hospital administered medications), medical procedures or deaths with a history of data collection spanning more than 50 years. All registers can be linked electronically using the unique personal identity code (PIC) which is assigned to every permanent resident of Finland (<https://dvv.fi/en/personal-identity-code>). Only people who have moved permanently abroad are lost to follow-up. The Finnish health registers, including their diagnostic validity and accuracy, have been reviewed previously^1–4^ and more recently using FinnGen data ^5,6^*.*

Health register based phenotypes, called endpoints, were created by combining data (mainly using ICD and ATC codes) from different nationwide heath registers (Supplementary Table 1). For a pheWAS approach we have initially constructed more than 2800 endpoints by combining data from different health registers. To create these endpoints, we have mainly used Finnish version of International Classification of Diseases (ICD 8,9,10), in the care register for health care, and in causes of deaths. To further add specificity and sensitivity of endpoints we enhance them by combining data from other registers, which include the Finnish Cancer Registry (ICD-O-3 codes) and prescription drug purchase (using Anatomical Therapeutic Chemical (ATC) codes) and medication reimbursement data (Supplementary Table 1). Besides the register data, smoking, BMI and gender information was provided by biobanks. The baseline data had been collected in different ways, either from hospital EHRs or in the case of legacy cohorts from study questionnaires. Clinical expert groups and Core FinnGen teams designed which codes are used to create each disease endpoint to optimize specificity and sensitivity. Supplementary Table 3 lists codes and code combinations used for the 15 demonstration endpoints. It illustrates the range of complexity e.g. for primary open angle glaucoma only the ICD-10 code H40.1 from discharge registers was needed (with corresponding ICD-8 and ICD-9 codes), whereas for Alzheimer’s disease and cancers codes from several registers were combined. Additional rules (e.g., age cut offs) for each health-related endpoint are listed as footnotes in Supplementary Table 3. For each endpoint, all recorded events with age at event were first recorded. Then so called first-ever event data was constructed, with age-of-onset for cases, and age at censoring or at the end of follow-up for controls. This data was used for the FinnGen core GWAS analyses.

The Finnish ICD-10 version which is used in hospital inpatient stays, hospital outpatient visits, and causes of deaths was the basis for the endpoint creation. The Finnish version is mostly identical to international ICD-10 classification with minor modifications mostly adding levels of specificity, i.e. there are additions to certain disease classifications in 4^th^ and 5^th^ character level. ICD definitions were then harmonized over different national ICD versions, ICD-10, ICD-9 and ICD-8. Mapping of the diagnosis codes (ICD8-10, ICD-O, SPAT, ICPC), Finnish therapeutic and diagnostic procedure codes (based on Nomesco), and pharmaceutical codes (ATC and VNR) into standardized OMOP-CDM vocabulary is ongoing ([OMOP Common Data Model – OHDSI](https://www.ohdsi.org/data-standardization/the-common-data-model/)).

There is an extensive literature describing the validity of these nationwide registers, thus a comprehensive validation within FinnGen was not prioritized^2–4,7,8^. The portal, Risteys (r5.risteys.finngen.fi for endpoints used in this manuscript, latest FinnGen endpoints: <https://risteys.finngen.fi/>) was designed for exploring FinnGen endpoints. Risteys is linked to MeSH (Medical Subject Headings) and SNOMED health terminology standards.

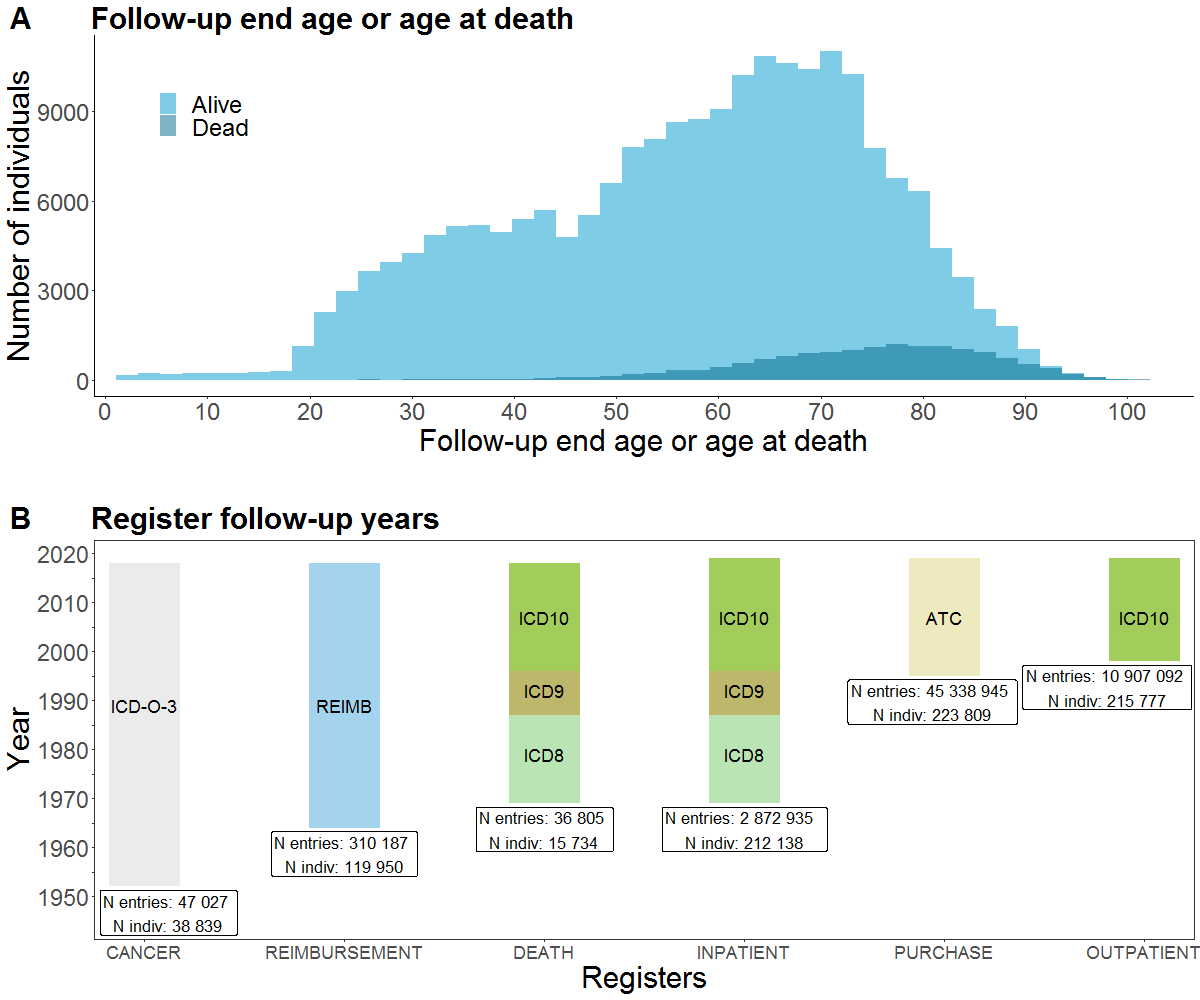

**Supplementary Figure 1. FinnGen Age Distribution and Registers.** A) Distribution of the current age (age at the end of the follow-up) and age of death for FinnGen participants B) Follow-up time and main coding used in each register among FinnGen participants in FinnGen release 5. Abbreviations: CANCER = The Finnish Cancer Registry; DEATH = Cause of death register; INPATIENT = HILMO - Care Register for Health Care: Inpatient hospital visits; OUTPATIENT = HILMO - Care Register for Health Care: Specialty outpatient visits and day surgeries; PURCHASE = Drug Purchases: All Prescription drug purchases; REIMBURSEMENT = Drug Reimbursement: entitlements for prescription drug reimbursement for certain chronic diseases.

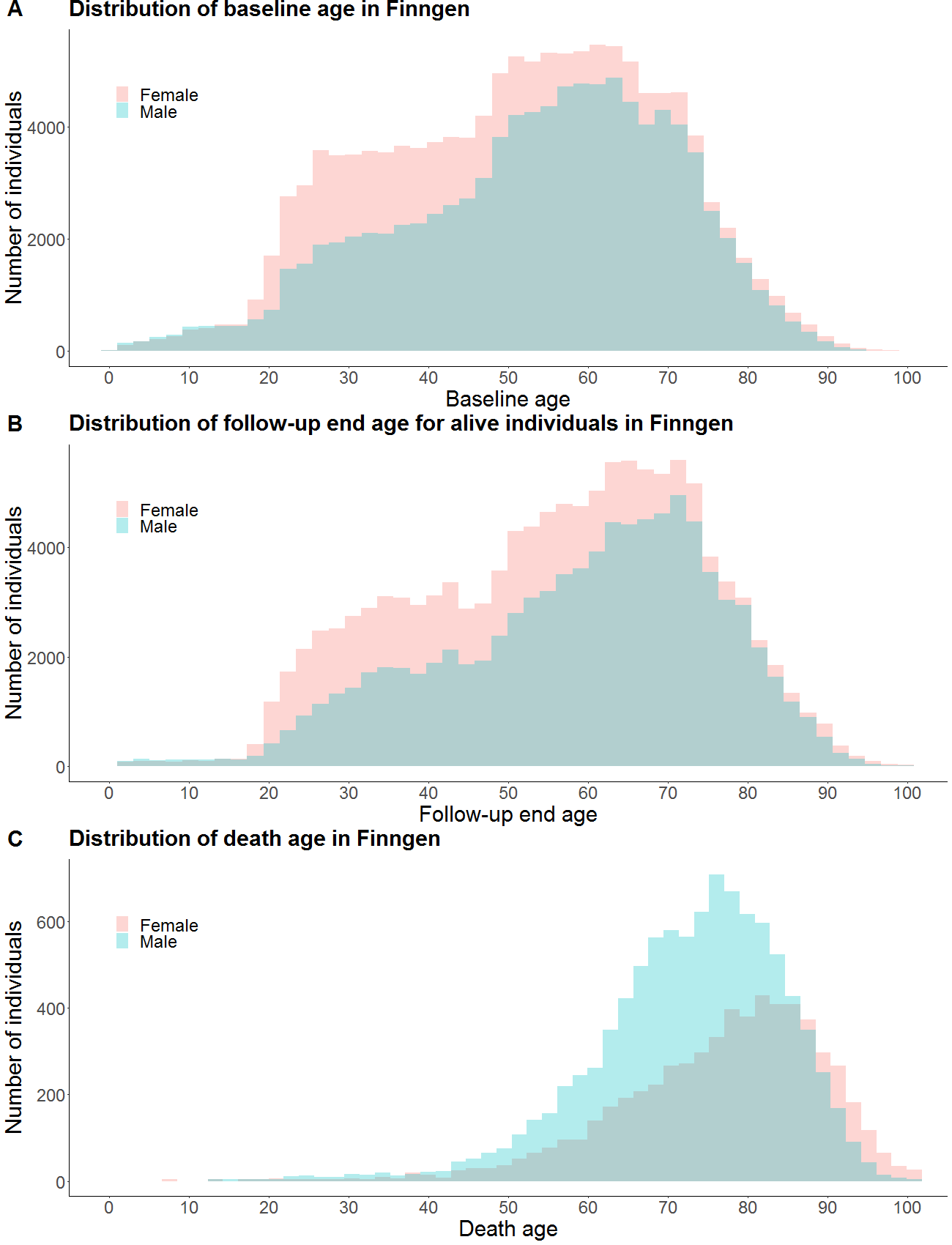

**Supplementary Figure 2.** Distribution of the participants A) age at the baseline (participation age), B) age at the end-of the follow-up or C) age at death.

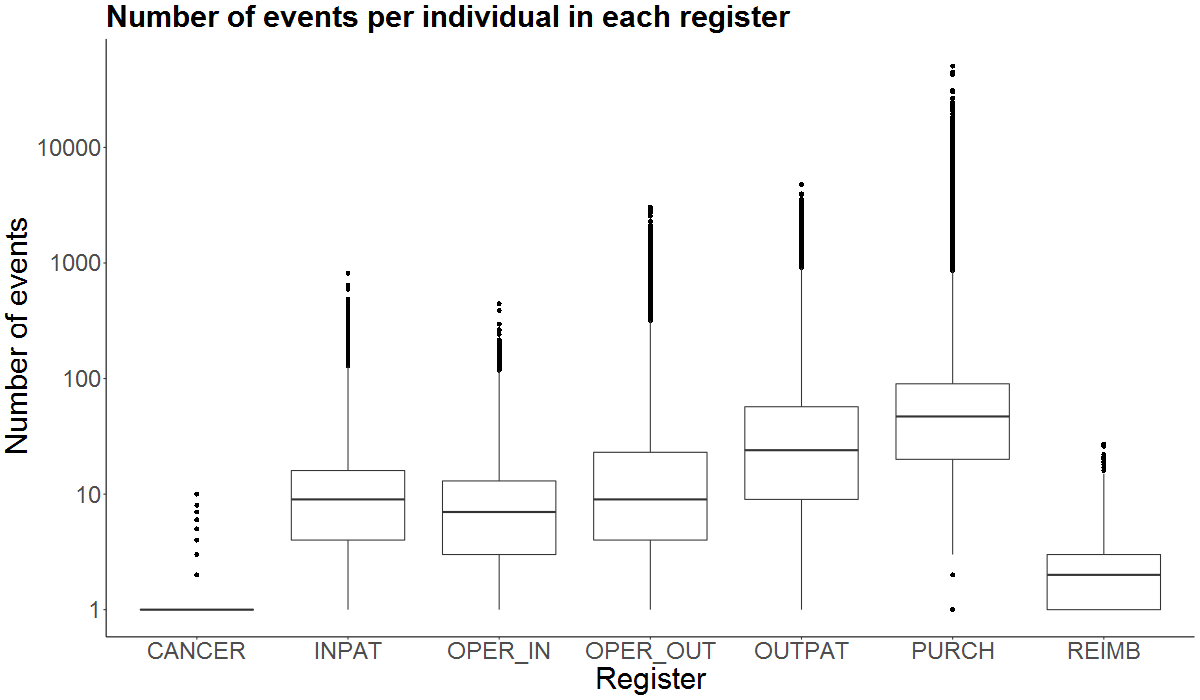

**Supplementary Figure 3.** Number of events (log scale) per individual in each of the registers. Abbreviations: CANCER = The Finnish Cancer Registry; DEATH = Cause of death register; INPAT = HILMO - Care Register for Health Care: Inpatient hospital visits; OUTPAT = HILMO - Care Register for Health Care: Outpatient visits and day surgeries; OPER_IN = Operations in inpatient HILMO register, OPER_OUT: Operations in outpatient HILMO register, PURCH = Drug Purchases: All Prescription drug purchases; REIMB = Drug Reimbursement: entitlements for prescription drug reimbursement.

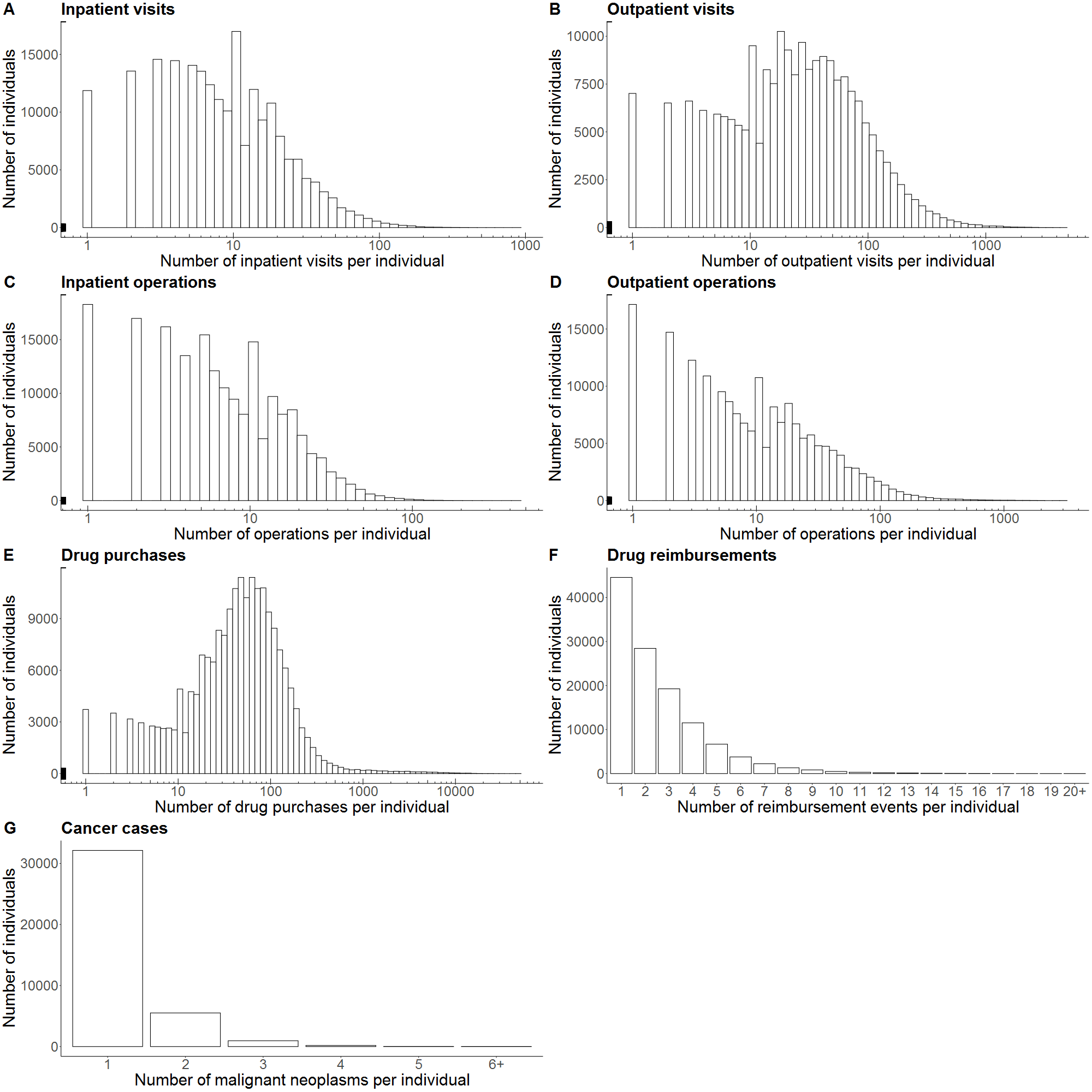

**Supplementary Figure 4**. Number of events (log scale for A-E) per individual in each of the registers.

### **Section 2**: FinnGen participant recruitment and legacy cohorts

The FinnGen study is a nationwide cohort of Finnish individuals, combining genome information with digital health care and registry data.

The FinnGen cohort consists of (a) legacy samples recruited by specific research projects, collected based on study-specific consents (Supplementary Table 2) and later transferred to a biobank according to the Finnish Biobank Act 13 § (mainly from the Biobank of the Finnish Institute for Health and Welfare (THL)), and (b) prospective biobank samples recruited by Finnish biobanks, collected based on a wide biobank consent, according to the Finnish Biobank Act 11 §. Prospective samples in the FinnGen cohort originate from three complementary approaches: (i) samples from six regional hospital biobanks represent a vast variety of patients enrolled in specialized health care, (ii) samples from a private health care biobank allow for targeted enrichment of the FinnGen cohort with patients underrepresented in specialized health care, and (iii) participants recruited through Blood Service Biobank enrich the cohort with healthier individuals. In the present study, we included samples from altogether 224,737 participants (FinnGen release 5, August 2020) from the following biobanks: Auria Biobank, Helsinki Biobank, Biobank of Eastern Finland, Central Finland Biobank, Northern Finland Biobank Borealis, Finnish Clinical Biobank Tampere, THL Biobank, Blood Service Biobank, and Terveystalo Biobank.

Various means were utilized for informing individuals about ongoing biobank recruitment, including active informing by hospital personnel, locally distributed leaflets, national advertising campaigns on TV and in social media, and dedicated biobank research nurses operating in hospital districts. Information about the FinnGen research project was mostly distributed to the public in news articles, interviews, leaflets and on the FinnGen website. Prospective samples collected by the hospital biobanks and the Blood Service Biobank were collected without any specific prioritization. All individuals volunteering to provide a biobank consent and to donate a biobank sample to a Finnish biobank were eligible for the FinnGen study Biobank recruitment and sampling protocols followed the protocols approved by the Finnish Medicines Agency (Fimea), the organization with the legal responsibility for supervising biobank operations in Finland. The Coordinating Ethics Committee of the Hospital District of Helsinki and Uusimaa (HUS) approved the FinnGen study protocol Nr HUS/990/2017.

.

DNA samples were obtained from participating biobanks. DNA was extracted either from whole blood or buffy coat according to the standard procedure of each biobank, and sent to a centralized logistics laboratory in the THL Biobank. THL Biobank laboratory aliquoted and normalized all samples in 96-well plates in 60 µl volume and 15 ng\µl concentration, and delivered the plated samples to Affymetrix Research Services Laboratory, Thermo Fisher Scientific (San Diego, USA).

### **Section 3:** Genotyping and genotype data QC

FinnGen Release 5 presented here contains genotype data for 224,737 individuals after quality control (QC). A total of 154,714 individuals were genotyped with custom Axiom FinnGen1 array and 70,023 individuals from legacy collections genotyped with non-custom genotyping arrays.

**Thermo Fisher array genotyping and genotype calling**

The FinnGen1 (v1) array, technically similar to the UK Biobank (UKBB) genotyping array, is a custom ThermoFisher Axiom array consisting of 736,145 probes interrogating 655,973 variants. In addition to the core imputation ‘backbone’, we added 116,402 (97,665 missense and 20,432 predicted loss of function) rare (MAF < 0.5%) coding variants (97,665 missense and 20,432 predicted loss of function) that had been identified in 15 000 Finnish exomes to be directly genotyped. We also added variants for the KIR and HLA haplotypes (10,800), known pathogenic ClinVar variants (14,900), pharmacogenomic markers (4,600) and 57,008 additional markers that were of special interest for the FinnGen partners investigators in academia and and industry.

DNA samples were plated and shipped to Thermo Fisher Scientific Inc. (San Diego, USA) on pre-barcoded 96-well plates provided by the company. Sample DNA quantity was assessed using the PicoGreen™ assay in the Thermo Fisher Microarray Services Research Laboratory (MRSL). Samples were processed using the Axiom™ 2.0 Assay 96-Array Format Automated Workflow (<https://assets.thermofisher.com/TFS-Assets/LSG/manuals/702963_Final_Ax_96FX_UG.pdf>), including in-line quality control measures. Two positive control samples were included on each 96-well processing plate.

Sample data quality was assessed using two metrics described in the Axiom™ Genotyping Solution Data Analysis User Guide (<https://assets.thermofisher.com/TFS-Assets/LSG/manuals/axiom_genotyping_solution_analysis_guide.pdf>). DQC is a single-sample metric that assesses the separation of foreground and background signal distributions on a set of control probes for non-polymorphic, autosomal loci. A DQC threshold of greater than or equal to 0.82 was applied. Samples falling below this threshold were excluded from further analysis. QC Call Rate (QCCR) is the genotype call rate over a set of 20,000 autosomal SNP probesets which were previously observed to have good performance in the assay and were included on the array with two replicates of each probe sequence, in a cluster of all samples with DQC greater than or equal to 0.82. Samples with QCCR less than 97% were excluded from further analysis.

Genotype calling was performed using the Array Power Tools (APT) software (<https://www.thermofisher.com/us/en/home/life-science/microarray-analysis/microarray-analysis-partners-programs/affymetrix-developers-network/affymetrix-power-tools.html>) with recommended settings. Individual probe sets were assessed using the QC metrics described in the Axiom™ Genotyping Solution Data Analysis User Guide (<https://assets.thermofisher.com/TFS-Assets/LSG/manuals/axiom_genotyping_solution_analysis_guide.pdf>) using the default thresholds, including call-rate ≥95%. Only genotype calls from probesets passing all QC thresholds were included in the data release.

**Legacy chip data genotyping and genotype calling**

Samples originating from various cohort collections were genotyped with different Illumina (Illumina Inc., San Diego, CA, USA) chips. Different sample collections were called with the either GenCall or zCall algorithm and the corresponding genotype calls were used to perform quality control procedures.

**Chip data QC**

In total there were 239173 samples (164794 samples genotyped with FinnGen custom Axiom chip, 74379 legacy samples). The basic quality control included checking of Mendelian errors, exclusion of samples having PI-HAT values >0.9 between sample pairs that were not monozygotic twins or replicate samples, samples with discrepant sex based information between reported sex in register and the sex chromosome and replicate samples with over 50,000 markers with discrepant results. All replicate samples except the one with the highest call rate for each replicate were excluded. Markers with replicate and Mendelian errors were not excluded but were tagged. The following quality control exclusion cut-off criteria were further used: sample-wise missingness rate >0.05%, heterozygosity rate greater than population average ±4 standard deviations (SD), sample contamination detection with PI-HAT >0.1 to more than 14 samples, principal component analysis (PCA) outliers in the first 2 dimensions considering ±4SD. Sex-check F-value ≤0.3 was used for female and ≥0.8 for expected male samples. Variant-wise QC was done by applying Hardy-Weinberg equilibrium test p-value cut-off of <10^-6^, call-rate of >98% and minimum allele count of 1. We ended up removing 10 080 Axiom genotyped samples and 4 356 legacy samples and retained 224 737 samples.

Before genotype imputation, all chip data was checked and mitochondrial, chromosome Y and unmapped or ambiguously mapped variants were excluded. All autosomal chromosomes and chromosome X data were lifted to human reference genome build 38 (GRCh38/hg38) and all reference and alternative alleles were converted and left-aligned according to the build 38 reference sequence as described in Pärn *et al*.^9^. Samples overlapping with the imputation reference panel were excluded. Variants with alternative allele count less than 3 (on batch level) were excluded. Alternative allele frequency (AF) comparison to the SISu v3 reference panel was done and variants not present in the panel or displaying significant AF differences to the panel were excluded (AF difference >10 percent points or AF fold change greater than ±5). Plink 2.0^10^ glm firth-fallback GWAS analysis was performed to detect further variants with significantly different AF (as compared to the SISu v3 reference panel). Variants were removed using a dynamic p-value cut-off of 5 x 10^-8^/genomic-ƛ^3^.

**Population-specific SISu v3 imputation reference panel**

After comprehensive QC procedures chip-genotyped samples were pre-phased and imputed with the population-specific SISu v3 imputation reference panel, developed from high-coverage (25-30x) whole-genome sequencing (WGS) data of 3,775 Finns^5^.

Raw high-coverage (25-30x) whole-genome sequencing (WGS) data for 4,083 Finnish samples were aligned to the GRCh38/hg38 human reference genome assembly using BWA-MEM (Li, 2013). PCR duplicates were marked using Picard^11^, and the Genome Analysis Toolkit (GATK) v3 best-practices^12,13^ were applied for variant calling. Genotype-, sample- and variant-wise quality control and filtering procedures were applied by using the Hail framework (<https://github.com/hail-is/hail>) v0.1 (unless mentioned otherwise). First, low-quality individual genotype calls were set missing. In order to perform the sample-wise filtering, autosomal biallelic variants outside of low-complexity regions (LCRs)^14^ were included if: call-rate (CR) was ≥90%; variant Hardy-Weinberg Equilibrium p-value (pHWE) was ≥1 x 10^-9^; variant quality score recalibration (VQSR) filter was ‘PASS’; quality by depth (QD) was ≥2 for SNV and ≥3 for indel variants and allele count (AC) was ≥3. Sample-wise metrics were calculated with Hail and outlier samples deviating more than ±3SD were identified from sample-wise QC metrics (nSNP, rHetHom, rInsertionDeletion, rTiTv). Next, relatedness was calculated and highly related individuals (KING^15^ kinship coefficient <0.177) were excluded. Then, the top 20 principal component (PC) scores were computed and outlier samples to be excluded were identified based on the first 10 PCs. By using the unfiltered ‘vanilla’ variant call set, variant-wise quality metrics were recalculated for 3,775 high-quality samples. Finally, only variants that met the following criteria were preserved: CR≥90%; VQSR ‘PASS’; QD≥2 for SNVs QD≥6 for indels; pHWE for autosomal chromosomes and for females on chromosome X was ≥ 1e-9; and AC≥3. The resulting samples and variant alleles were pre-phased with Eagle v2.3.5^16^ using the default values (except for the Kpbwt that was set to 20,000). The final SISu v3 imputation reference panel included 3,775 high-quality samples and 16,962,023 MAC>=3 SNV/indel variant alleles ^5^.

**Chip data pre-phasing and genotype imputation**

Chip genotyped samples were pre-phased with Eagle 2.3.5 (<https://data.broadinstitute.org/alkesgroup/Eagle/>) with the default parameters, except the number of conditioning haplotypes was set to 20,000. Genotypes were imputed with the population-specific SISu v3 imputation reference panel, resulting. This resulted in 16 387 711 confidently imputed (INFO>0.6, 16 962 023 imputed variants in total) variants, imputed accurately

**Genotype imputation accuracy estimation**

The ‘best guess’ genotype calls for confidently imputed variants (INFO>0.6) for 6,542 chip-genotyped samples were compared to the whole-exome sequencing (WES) data for the same individuals. Quality control procedures for these WES data were carried out in a similar manner as for the reference panel WGS data. Sample-wise metrics were calculated with Hail and outlier samples deviating more than ±3SD for nSNP, rHetHom, rInsertionDeletion and rTiTv were excluded. Variants inside of low-complexity regions (LCRs)^14^ were excluded, low-quality individual genotype calls were set missing, after which variant-wise call-rate of ≥90% and Hardy-Weinberg Equilibrium p-value (pHWE) cut-off of ≥1 x 10^-9^ were applied. Variant quality score recalibration (VQSR) filter ‘PASS’ was applied together with quality by depth (QD) of ≥2 for SNV and ≥3 for indel variants. Finally, only autosomal biallelic variants with allele count (AC) ≥1 were included.

While treating the WES-based genotype calls as 'gold standard', GATK GenotypeConcordance was used to calculate non-reference sensitivity (the proportion of WES-based non-reference genotype calls that were obtained through the imputation process) and discordance rate (the proportion of non-reference imputed or WES-based genotypes, which had discordant genotype calls).

Upon completion of the steps described in this section, this resulted in 16 387 711 confidently imputed (INFO>0.6, 16,962,023 imputed variants in total) variants across the allele frequency spectrum (Figure 1 A).

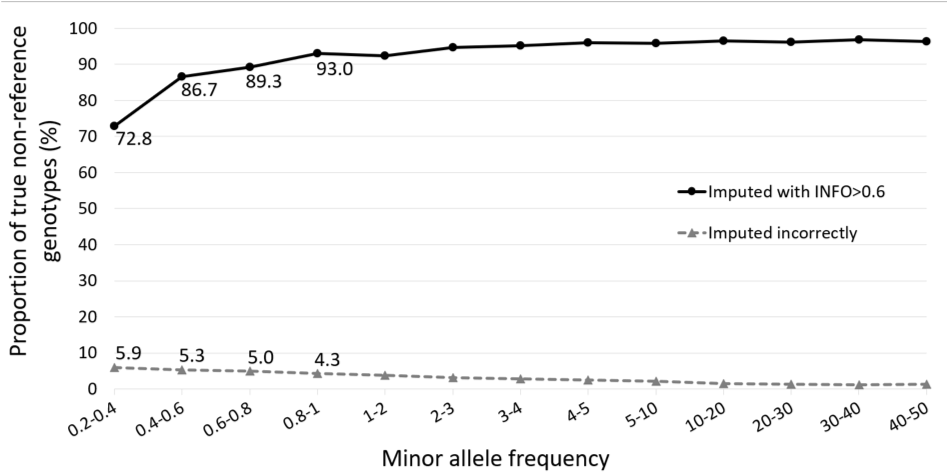

***Supplementary Figure 5.*** *FinnGen coding variant imputation accuracy among confidently imputed (INFO > 0.6) variants. Imputed best guess non-reference genotypes were compared to exome sequenced genotypes from the same individuals. The sensitivity to identify non-reference genotypes (black continuous line) and false positive rate (grey dashed line) are shown in different allele frequency bins*

### **Section 4:** Population structure and cryptic relatedness

We merged 224,737 FinnGen samples with 1000 genomes phase 3 data set into a single data set of 49451 pruned SNPs, on which we calculated PCs. We first removed 2104 duplicated samples. We then utilized Bayesian unsupervised method Aberrant^17^ to identify samples that deviated from the majority of the samples in PCA space. The vast majority of FinnGen samples clustered in their own cluster next to 1000 genomes of Europeans (Supplementary Figure 5). In outlier detection, 3138 FinnGen samples and all 1000 genomes samples except 96 out of 99 Finns were identified as significantly deviating from the majority of the samples (Supplementary Figure 5) and were removed. Since all non-European 1000 genomes samples had been removed, we used the remaining 404 North-Western Europeans and 99 Finn 1000 genomes samples as known samples of other European and Finnish populations to build a classifier. PCA was calculated for the remaining 219,585 FinnGen samples and the 1000 genomes known samples were projected onto the same space. For each FinnGen sample, a probability of being part of the EUR/FIN cluster was calculated, using a chi-squared distribution based on the Mahalanobis distance to the centroid of each cluster. We classified as ethnic Finns the individuals whose relative probability of being part of the FIN cluster was > 95%, resulting in additional 538 outliers being removed (Supplementary Figure 6). The remaining 218,957 samples were retained as Finnish ancestry samples**.** The relationships between remaining samples was estimated with KING software^15^ (Supplementary Figure 7, Supplementary Table 4)

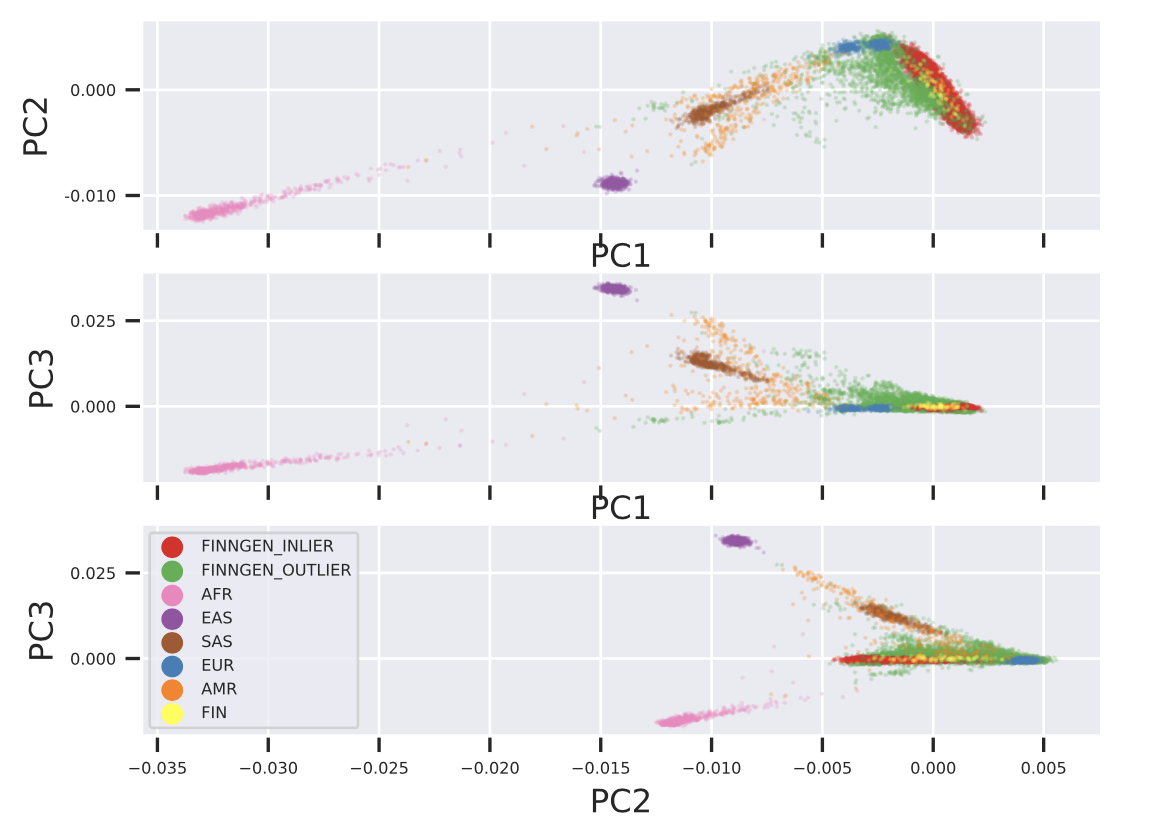

**Supplementary Figure 6**. PCA classification of 224,737 FinnGen participants combined with 1000 genomes samples (AFR,AMR,EAST,EUR,FIN,SAS). FinnGen outlier samples were removed as deviating from the bulk of the FinnGen samples.

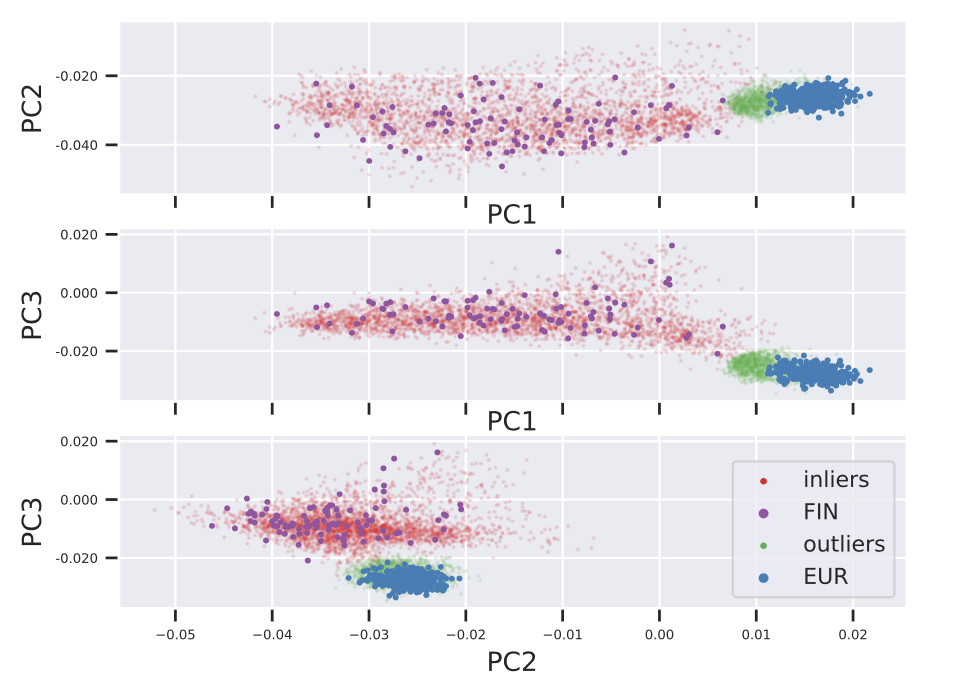

**Supplementary Figure 6**. Classification of Finnish ancestry participants based on 1000 genomes learning samples (EUR, FIN) remaining after 1st round of outlier removal (see Supplementary Figure 5 ). 538 “Outliers” (in green) removed and 218957 “inliers” (in red) kept.

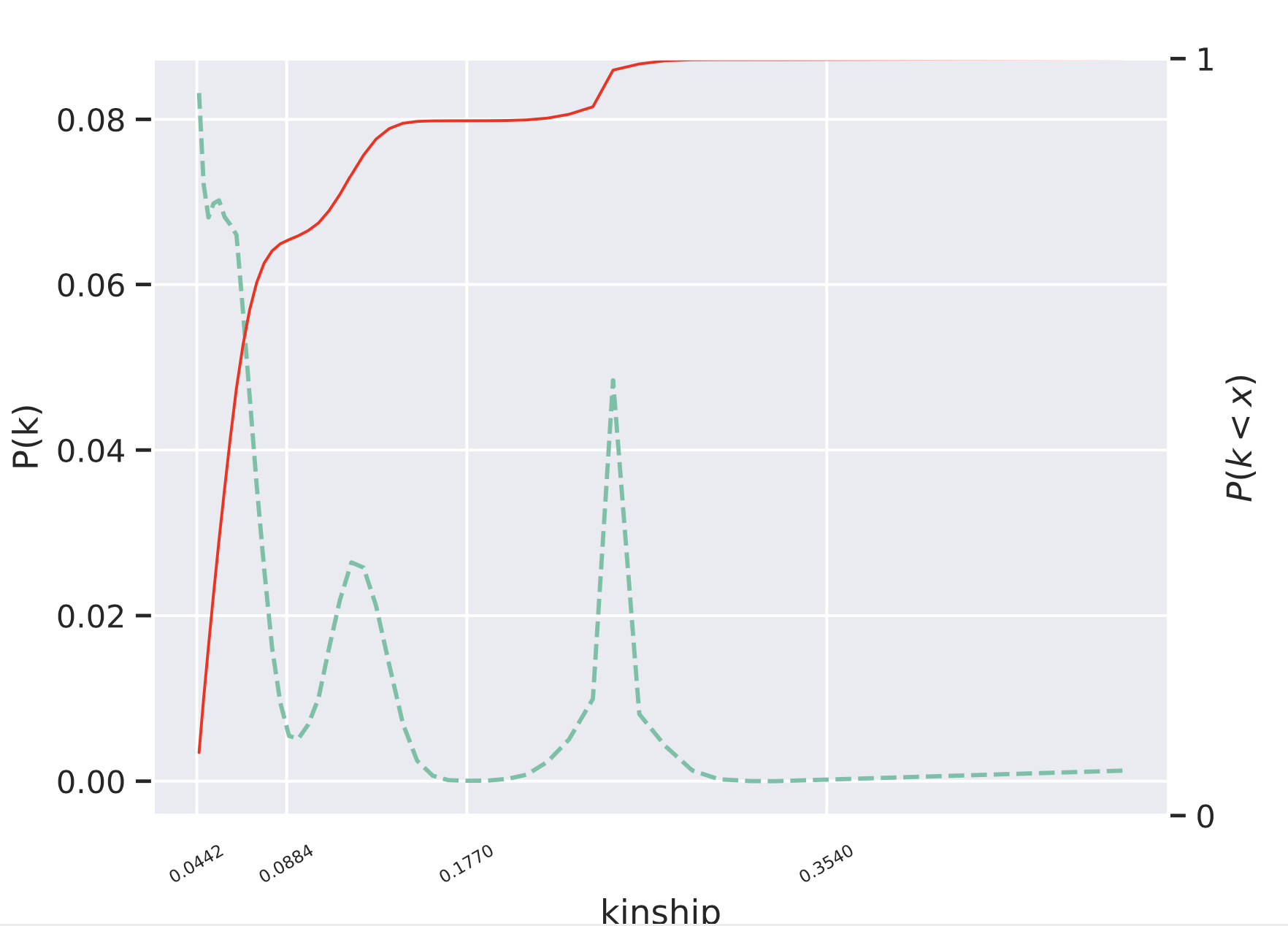

**Supplementary Figure 7**. Density (green dashed lines) and cumulative distribution plots (red solid line) of FinnGen R5 KING kinship estimates. Kinship value intervals >0.354, [0.1770-0.3540], [0.0884-0.1770), and [0.0442-0.0884) were used to classify duplicates/monozygotic twins, 1st, 2nd and 3rd degree relatives, respectively.

### **Section 5**: GWAS/pheWAS Analysis

#### Data Access and Dissemination

All summary statistics for each release are generated by the FinnGen analysis team (pheWAS, finemap, colocalization) and made available via a custom FinnGen PheWeb (<https://results.finngen.fi/>) as well as file downloads (see data access).

FinnGen also provides versatile data access to individual level data for FinnGen Partners while maintaining the appropriate legal framework and level of security by which sensitive Personal Data can be accessed. The European and National data protection and data security regulations and national authority requirements were followed when designing data access solutions.

The custom-built computational environment, called FinnGen Sandbox, is deployed in Google Cloud, using servers located in the EU. Individual level data can be analysed by all FinnGen researchers who have completed appropriate training and have been authorized by the Finnish Social and Health Data Permit Authority Findata (<https://findata.fi/en/>) to access and analyse individual level medical and genetic data. Sandbox is an interactive virtual machine in Google Cloud with graphical user interface and pre-installed suite of data-analysis software and analysis environments. Computationally heavy analyses (e.g. custom GWAS analyses) are supported utilizing parallel Google Cloud worker machines via Cromwell ^18^ pipeline server. The researchers can bring their own analysis scripts and external data but cannot download or export individual-level data from the Sandbox environment. Summary level data can be exported via a request and the data are manually inspected to ensure no personal level data are exported.

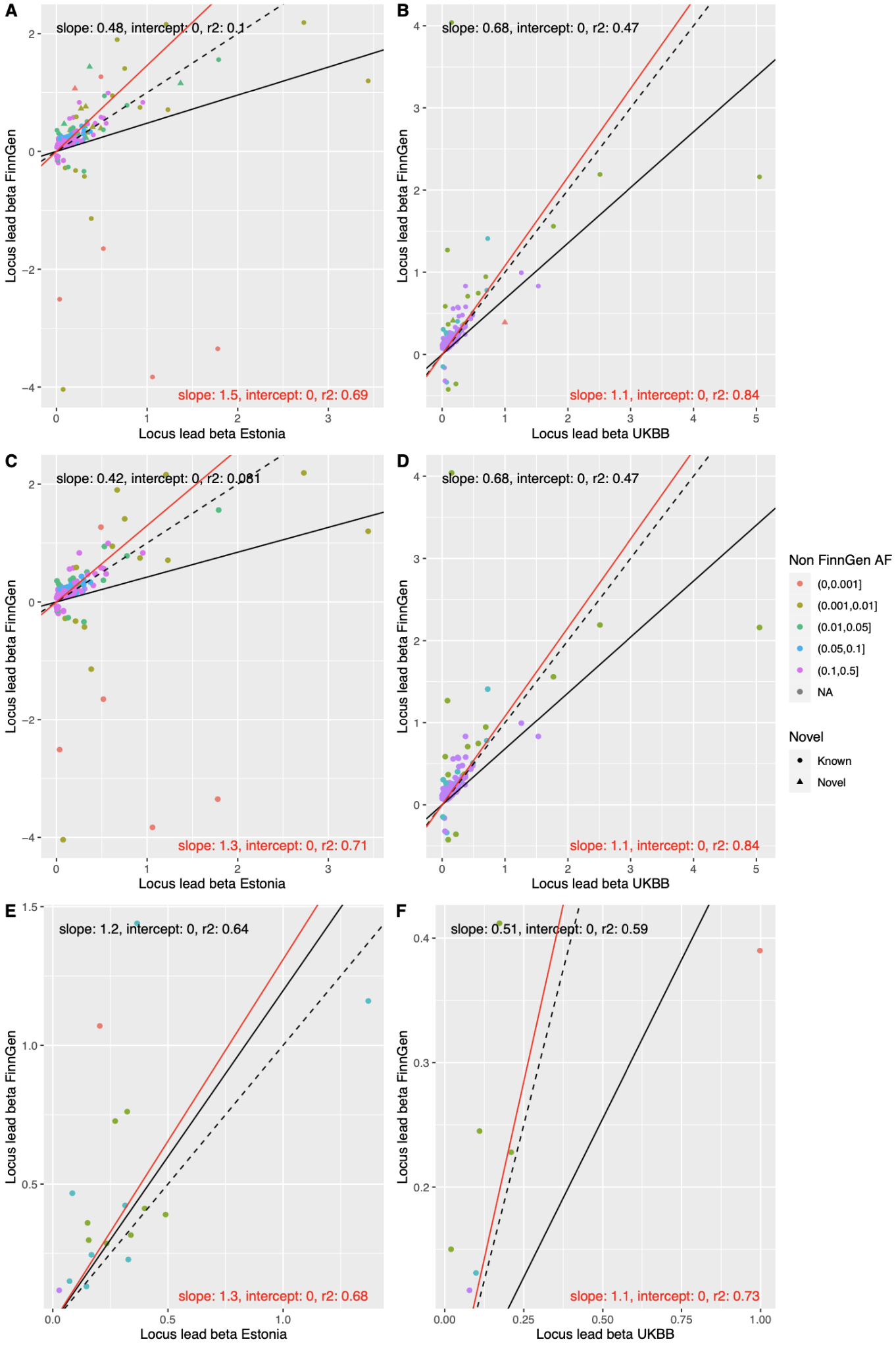

**Supplementary Figure 8**. A,B) Effect size (log(OR), beta) comparison of 275 genome-wide significant lead variants identified in FinnGen among 15 analysed diseases in Estonia and UKBB. The sign of beta is aligned to be positive in Estonia and UKBB. C,D) beta comparison of variants only in known loci. E,F) beta comparison of novel loci. Dashed lines indicates identity line and solid lines are the regression line (red line and text weighted by pooled standard error of betas)

12.	Scaling accurate genetic variant discovery to tens of thousands of samples | bioRxiv. https://www.biorxiv.org/content/10.1101/201178v3.
